# Family-GWAS reveals effects of environment and mating on genetic associations

**DOI:** 10.1101/2024.10.01.24314703

**Authors:** Tammy Tan, Hariharan Jayashankar, Junming Guan, Seyed Moeen Nehzati, Mahdi Mir, Michael Bennett, Esben Agerbo, Rafael Ahlskog, Ville Pinto de Andrade Anapaz, Stefania Benonisdottir, Dorret I. Boomsma, Archie Campbell, Christopher F. Chabris, Zhengming Chen, China Kadoorie Biobank Collaborative Group, Eco de Geus, Erik A. Ehli, Abdelrahman G. Elnahas, Estonian Biobank Research Team, Finngen, Andrea Ganna, Alexandros Giannelis, Liisa Hakaste, Caroline Hayward, Jouke-Jan Hottenga, Mikkel Aagaard Houmark, Jaakko Kaprio, Antti Latvala, James J. Lee, Mikko Lehtovirta, Liming Li, LifeLines Cohort Study, Kuang Lin, Richard Karlsson Linnér, Stefano Lombardi, Nicholas G. Martin, Matt McGue, Sarah E. Medland, Andres Metspalu, Brittany L. Mitchell, Guiyan Ni, Ilja M. Nolte, Matthew T. Oetjens, Sven Oskarsson, Teemu Palviainen, Rashmi B. Prasad, Anu Reigo, Kadri Reis, Julia Sidorenko, Karri Silventoinen, Harold Snieder, Tiinamaija Tuomi, Bjarni J. Vilhjálmsson, Robin G. Walters, Emily A. Willoughby, Jonathan Flint, Loic Yengo, Peter M. Visscher, Augustine Kong, Elliot M. Tucker-Drob, Richard Border, David Cesarini, Patrick Turley, Aysu Okbay, Daniel J. Benjamin, Alexander Strudwick Young

## Abstract

Genome-wide association studies (GWAS) have discovered thousands of replicable genetic associations, guiding drug target discovery and powering genetic prediction of human phenotypes and diseases. However, genetic associations can be affected by gene-environment correlations and non-random mating, which can lead to biased inferences in downstream analyses. Family-based GWAS (FGWAS) uses the natural experiment of random assignment of genotype within families to separate out the contribution of direct genetic effects (DGEs) — causal effects of alleles in an individual on an individual — from other factors contributing to genetic associations. Here, we report results from an FGWAS meta-analysis of 34 phenotypes from 17 cohorts. We found evidence that factors uncorrelated with DGEs make substantial contributions to genetic associations for 27 phenotypes, with population stratification confounding — a form of gene-environment correlation — likely the major cause. By estimating SNP heritability and genetic correlations using DGEs, we found evidence that assortative mating has led to overestimation of SNP heritability for 5 phenotypes and overestimation of the degree of shared genetic effects (pleiotropy) between 22 pairs of phenotypes. Polygenic predictors constructed from DGEs are particularly useful for studying natural selection, assortative mating, and indirect genetic effects (effects of relatives’ genes mediated through the family environment). We validate our meta-analysis results by predicting phenotypes in hold-out samples using polygenic predictors constructed from DGEs, achieving statistically significant out-of-sample prediction for 24 phenotypes with little attenuation of predictive power within-families. We provide FGWAS summary statistics for 34 phenotypes that can be used for downstream analyses. Our study provides both a template for performing FGWAS and an argument for its value for debiasing inferences and understanding the impact of environment and mating patterns.

## Introduction

Genome-wide association studies (GWASs) have generated biological insights, guided drug target discovery^1^, and enabled prediction of phenotypes and disease risks using polygenic predictors (called polygenic indices, PGIs, or polygenic scores), with potential clinical applications^2^. However, recent research^3–10^ has shown that GWASs are susceptible to confounding from indirect genetic effects (IGEs) from relatives — e.g. an effect of parents’ alleles on offspring education mediated through the alleles’ effects on parental education^11^ — assortative mating (when there are correlations across parents’ phenotypes and genotypes), and population stratification. Analytical techniques including principal component analysis (PCA) and linear mixed models have been developed to reduce population stratification confounding^12^, but these techniques often leave residual confounding in GWAS results^5,8,10^. Confounding can cause biases in downstream applications, including: estimation of heritability and genetic correlation^6,7^, Mendelian Randomization analyses^13^, and inferences of natural selection^4,5^.

Family-based GWAS (FGWAS) has been proposed as a solution to the problem of confounding in GWAS^3,9,14–16^ that also enables deeper investigation of the impact of gene-environment correlation — including from IGEs — and non-random mating, including population structure and assortative mating. FGWAS adds the parents’ genotypes to the regressions performed in GWAS (Methods), thereby using the natural experiment of random assignment of genotype within-family — due to Mendelian segregation during meiosis — to estimate ‘direct genetic effects’ (DGEs). Because the segregation of chromosomes during meiosis is independent of environment, estimates of DGEs are free from confounding due to gene-environment correlation, including from IGEs and population-stratification^3,9,15,16^. Because different chromosomes segregate independently during meiosis, DGEs are also free from the confounding that arises in GWAS due to correlations with genetic variants on other chromosomes,^15,16^ which are caused by non-random mating, including population structure and assortative mating (AM). As in GWAS, variants are analyzed one at a time, so DGEs include causal effects of the focal variant and correlated variants on the same chromosome^14–16^.

The coefficients on the parental genotypes are referred to as ‘non-transmitted coefficients’ (NTCs), since they are the expected coefficients on the alleles not transmitted from parents in a regression of phenotype onto transmitted and non-transmitted alleles^3,11,17^. For each parent, whether one or the other allele (e.g. ‘A’ or ‘T’ at a SNP) is transmitted is the random outcome of meiosis. The association between the non-transmitted allele and offspring phenotype (controlling for the transmitted allele) reflects all the factors causing genotype-phenotype association other than the allele being transmitted to the offspring and its direct causal effect on the offspring’s phenotype. These factors include gene-environment correlation due to IGEs and population stratification as well as confounding due to correlations with genetic variants across the genome due to non-random mating^15,16^. The non-transmitted parental alleles are perfect control alleles that differ from the transmitted allele only in the fact that one was randomly transmitted and the other was not. The use of non-transmitted alleles as control variables forms the basis of population-structure robust association tests such as the transmissions disequilibrium test (TDT)^18,19^.

We can relate the parameters estimated in FGWAS to the parameter estimated in GWAS, which we call the ‘population effect’ as it reflects the genotype-phenotype association in the population. Let *β*_*l*_ be the population effect of SNP *l* — as estimated in GWAS — and let *δ*_*l*_, *α*_*pl*_, *α*_*ml*_ be, respectively, the DGE and paternal and maternal NTCs. Under random-mating, *β*_*l*_ = *δ*_*l*_ + *α*_*l*_, where *α*_*l*_ = (*α*_*pl*_ + *α*_*ml*_)/2 is the average NTC^3^ and deviations from this relationship are negligible for samples with low levels of structure^3^, such as those typically used in GWAS. FGWAS thus removes confounding factors, reflected in the average NTC, that can lead to biased inferences in downstream applications of GWAS.

While FGWAS has favorable theoretical properties, it requires parental genotypes, which are often not available in typical GWAS samples. Furthermore, even when parental genotypes are available, estimates of DGEs are less precise than estimates of population effects because they only use within-family genotype variation, which is half of the variation in a random-mating population^3^. The lack of precise FGWAS results has limited their application.

An alternative study design, which we call sib-GWAS, uses genetic differences between siblings to estimate DGEs^3,9^. Because genotype data on sibling pairs is more commonly available than complete genotype data on parents, sib-GWAS has been a popular approach^9,13^, although it is biased by IGEs between siblings^3,15,16^. The sib-GWAS approach was used by Howe et al.^9^, which produced sib-GWAS estimates of DGEs on 24 phenotypes from a meta-analysis of European genetic ancestry cohorts.

Young et al. presented an alternative approach that enables sibling pair data, parent-offspring pair data, and complete parental genotype data to be jointly analyzed in a unified analytical framework through imputation of missing parental genotypes^3,20^. When applied to sibling pair data, this approach increases the effective sample size of DGE estimates by up to one third and of average NTCs by up to one half compared to sib-GWAS^3^. It also enables use of samples with only one or both parents genotyped, without genotyped sibling(s), further increasing power^3,20^. The imputation approach has been proven to give consistent and unbiased estimates of DGEs provided that the imputed parental genotypes are unbiased^3^. Although population structure can introduce bias into imputed parental genotypes, the resulting bias in DGE estimates is negligible for the samples of relatively homogeneous genetic ancestry typically used in GWAS^3,20^.

Here we report results from a meta-analysis of 16 cohorts of European genetic ancestries and one of East Asian genetic ancestry, analyzed using the imputation and FGWAS methodology implemented in the software package *snipar*^3^. We provide FGWAS summary statistics on 34 phenotypes spanning biomedical, psychiatric, and socioeconomic phenotypes, including 18 phenotypes not included in the Howe et al. sib-GWAS. We find evidence that there is substantial population stratification bias in population-effect estimates across a broad range of phenotypes and that assortative mating biases estimates of SNP heritability and genetic correlations. We validate our meta-analysis by performing family-based polygenic prediction analysis in holdout samples. Our results show the value of FGWAS for investigating the impact of gene-environment correlation and non-random mating, as well as debiasing inferences drawn from downstream applications of GWAS.

## Results

### FGWAS requires more stringent quality control than GWAS

We performed FGWAS in each cohort using the subsample with at least one genotyped parent or sibling, imputing the missing parental genotype(s) using *snipar*^3^ (Methods and Supplementary Table 1). We developed a quality-control (QC) protocol for FGWAS summary statistics — including novel QC procedures that take advantage of the unique properties of family data — that is described in Supplementary Note Section 1 and Supplementary Figures 1-2.

While QC for FGWAS shares many steps with QC for GWAS, there are some important differences. Most GWAS use data derived from genotyping arrays, which measure genotypes at pre-specified variants. Missing variants are imputed by finding similar haplotypes in reference samples^21,22^, without use of known pedigree relations and Mendelian Laws. In contrast, the Mendelian Imputation performed by *snipar* imputes genotypes of missing parents from the observed genotypes in a nuclear family according to Mendelian Laws.

We found that, except for the highest quality imputed variants, standard imputation from reference panels did not preserve the relationships between siblings’ genotypes implied by Mendelian Laws, implying that only the highest quality imputed variants are suitable for FGWAS or sib-GWAS (Supplementary Note Section 1 and Supplementary Figure 1). We therefore imposed a stringent imputation quality threshold (INFO score at least 0.99) for our analysis plan (Supplementary File 1). In this regard, our study differs from the sib-GWAS performed by Howe et al., which used variants with INFO score greater than 0.3, implying many low-quality imputed variants were analysed, which may have affected the results and conclusions drawn from their study.

### Multivariate meta-analysis of FGWAS summary statistics facilitates downstream analyses

We used fixed-effect, multivariate meta-analysis to aggregate the FGWAS summary statistics from 16 cohorts of predominantly European genetic ancestries (Methods, Supplementary Tables 1-2, and Supplementary Figure 3). This enabled us to compute meta-analysis estimates of DGEs, NTCs, and population effects, along with their joint sampling variance-covariance matrix, facilitating downstream analyses. Due to cross-ancestry differences in linkage disequilibrium (LD) patterns, we analysed the summary statistics from the China Kadoorie Biobank separately.

Although not necessary to remove confounding from estimates of DGEs, we included genetic principal components as covariates in cohort-level analyses to ensure that population effects derived from our analyses are comparable to those derived from standard GWAS^3,20^. To avoid under-powered analyses, we excluded from further analysis phenotypes where our meta-analysis effective sample size for DGEs was below 5000: ever-cannabis, chronic obstructive pulmonary disease (COPD), alcohol use disorder, and extraversion. This left 30 phenotypes for the subsequent analyses that we report on below (Table 1).

**Table 1.** Meta-analysis results. Results are based on multivariate meta-analysis of summary statistics from 16 cohorts of European genetic ancestries (Methods). Median effective sample sizes were calculated among HapMap3 SNPs for both direct genetic effects (DGEs) and population effects derived from the same data. Effective sample size is the sample size of a standard GWAS in unrelated individuals that would produce estimates of equivalent precision. Due to the stringent QC requirements of FGWAS, effective sample size varies considerably across the genome, with subsets of variants that passed QC in all or nearly all cohorts having greater effective sample size than given here (median in HapMap3). SNP heritability was estimated using LDSC (Methods). For binary outcomes, estimates were transformed to the logistic scale before calculation of effective sample sizes and SNP heritability (Supplementary Note Section 1). Genome-wide correlations between direct genetic effects (DGEs) and population effects are estimated using both LDSC and *snipar* (see Methods and Figure 1 note). See Supplementary Table 3 for an expanded set of meta-analysis statistics. Abbreviations: Pop., population effect; FEV1, forced expiratory volume in 1 second; ADHD, attention deficit and hyperactivity disorder; HDL, high density lipoprotein.

| Phenotype | Median effective<br>N (HapMap3) |  | SNP heritability |  |  |  | DGE-population correlation |  |  |  |
| --- | --- | --- | --- | --- | --- | --- | --- | --- | --- | --- |
|  | DGE | Pop. | DGE | S.E. | Pop. | S.E. | *snipar* | S.E. | *LDSC* | S.E. |
| Height | 105993 | 182202 | 0.352 | 0.020 | 0.413 | 0.021 | 0.913 | 0.005 | 0.970 | 0.005 |
| BMI | 81870 | 178153 | 0.212 | 0.013 | 0.216 | 0.012 | 0.919 | 0.013 | 1.006 | 0.008 |
| Educational attainment (EA) | 47387 | 91221 | 0.072 | 0.008 | 0.143 | 0.007 | 0.689 | 0.023 | 0.859 | 0.028 |
| ADHD | 44748 | 102327 | 0.005 | 0.014 | 0.003 | 0.007 | 0.372 | 0.016 | NA | NA |
| Non-HDL cholesterol | 42160 | 90474 | 0.168 | 0.023 | 0.179 | 0.021 | 0.819 | 0.027 | 0.959 | 0.030 |
| Number of children | 41589 | 102329 | 0.041 | 0.009 | 0.039 | 0.004 | 0.672 | 0.049 | 1.083 | 0.083 |
| HDL cholesterol | 40029 | 79576 | 0.191 | 0.035 | 0.181 | 0.024 | 0.871 | 0.068 | 0.944 | 0.026 |
| Age at first birth (women) | 35982 | 87944 | 0.044 | 0.012 | 0.097 | 0.008 | 0.495 | 0.019 | 0.881 | 0.067 |
| Self-rated health | 35443 | 83433 | 0.043 | 0.013 | 0.062 | 0.008 | 0.614 | 0.048 | 0.812 | 0.078 |
| Blood pressure (systolic) | 32532 | 72193 | 0.097 | 0.016 | 0.109 | 0.010 | 0.804 | 0.043 | 0.952 | 0.033 |
| Blood pressure (diastolic) | 32530 | 71625 | 0.102 | 0.017 | 0.111 | 0.011 | 0.720 | 0.030 | 0.965 | 0.033 |
| Neuroticism | 31649 | 75046 | 0.084 | 0.013 | 0.075 | 0.007 | 0.668 | 0.028 | 0.902 | 0.042 |
| Depressive symptoms | 31132 | 75497 | 0.060 | 0.015 | 0.035 | 0.007 | 0.619 | 0.052 | 0.666 | 0.098 |
| Subjective well-being | 28232 | 65930 | 0.026 | 0.016 | 0.048 | 0.009 | 0.454 | 0.053 | 0.819 | 0.148 |
| Migraine | 25907 | 67816 | 0.055 | 0.021 | 0.069 | 0.009 | 0.920 | 0.043 | 0.928 | 0.077 |
| Drinks per week | 22137 | 50345 | 0.027 | 0.022 | 0.029 | 0.012 | 0.561 | 0.053 | 0.595 | 0.217 |
| Allergic rhinitis | 21247 | 50657 | 0.086 | 0.026 | 0.082 | 0.015 | 0.793 | 0.028 | 0.794 | 0.065 |
| Age-at-menarche | 19678 | 45504 | 0.177 | 0.029 | 0.216 | 0.021 | 0.759 | 0.024 | 0.954 | 0.034 |
| FEV1 | 18645 | 45121 | 0.167 | 0.022 | 0.138 | 0.012 | 0.880 | 0.042 | 0.940 | 0.032 |
| Cigarettes per day | 16121 | 37207 | 0.014 | 0.022 | 0.063 | 0.015 | NA | NA | 1.027 | 0.513 |
| Ever-smoker | 14935 | 34550 | 0.356 | 0.029 | 0.463 | 0.022 | 0.801 | 0.014 | 0.974 | 0.019 |
| Morning person | 13347 | 36632 | 0.081 | 0.042 | 0.109 | 0.018 | 0.843 | 0.077 | 0.904 | 0.146 |
| Household income | 12884 | 31956 | 0.045 | 0.038 | 0.107 | 0.016 | 0.446 | 0.040 | 0.911 | 0.297 |
| Cognitive performance | 12361 | 26345 | 0.188 | 0.027 | 0.186 | 0.016 | 0.543 | 0.036 | 0.975 | 0.045 |
| Depression | 12216 | 31531 | 0.025 | 0.015 | 0.082 | 0.010 | 0.679 | 0.032 | NA | NA |
| Hypertension | 7506 | 18771 | 0.397 | 0.091 | 0.372 | 0.046 | NA | NA | 0.837 | 0.067 |
| Asthma | 6549 | 16229 | 0.360 | 0.077 | 0.378 | 0.048 | 0.865 | 0.035 | 0.963 | 0.035 |
| Eczema | 6326 | 16139 | 0.134 | 0.072 | 0.169 | 0.036 | 0.781 | 0.040 | 1.019 | 0.123 |
| Myopia | 5498 | 13859 | 0.526 | 0.110 | 0.517 | 0.050 | 0.870 | 0.028 | 0.828 | 0.043 |
| Individual income | 5489 | 14742 | 0.024 | 0.032 | 0.041 | 0.013 | NA | NA | NA | NA |

### Correlations between direct genetic effects and population effects

An important question is the degree to which DGEs, as estimated by FGWAS, differ from population effects, as estimated by GWAS. Howe et al.^9^ argued that DGEs are systematically smaller in magnitude than population effects for several phenotypes. While inflation of population effects leads to inflated estimates of SNP heritability, for many purposes low genome-wide correlation between DGEs and population effects is more problematic than inflation/deflation of effects; for example, under this scenario, polygenic predictors derived from population effects will never achieve perfect correlation with the DGE component of the phenotype, implying they will never capture the full heritability^8,15,23^.

The causes of low correlation between DGEs and population effects are likely distinct from the causes of inflation/deflation of population effects. For example, for confounding due to population stratification, the variant-level bias is likely unrelated to the variant’s DGE^10,24^. In that case, population effects differ from DGEs due to a random bias term with mean zero, which would produce a correlation below 1 but no systematic inflation/deflation. In contrast, classical AM would be expected to inflate population effects by a constant scale factor relative to DGEs —with the inflation reflecting the strength of correlation between parents’ DGE components^23,25^ —which would not affect the genome-wide correlation between DGEs and population effects.

Young et al.^3^ showed that the correlation between DGEs and population effects is below one for educational attainment (EA) and cognitive performance in UK Biobank data. Using an improved and expanded version of the method developed in Young et al. (implemented in *snipar*), we estimated the genome-wide correlation between meta-analysis estimates of DGEs and populations effects (Methods and Supplementary Note Section 2). The genome-wide correlation between DGEs and population effects is below 1 (FDR<0.05, one-sided test after Benjamini-Hochberg correction, which we use hereafter for multiple-testing correction) for 27/30 phenotypes (Figure 1, Table 1, Supplementary Table 3). The correlation could not be estimated for the remaining 3 phenotypes (cigarettes-per-day, hypertension, and individual income) due to negative estimates of the variance in DGEs and/or population effects.

**Figure 1.**
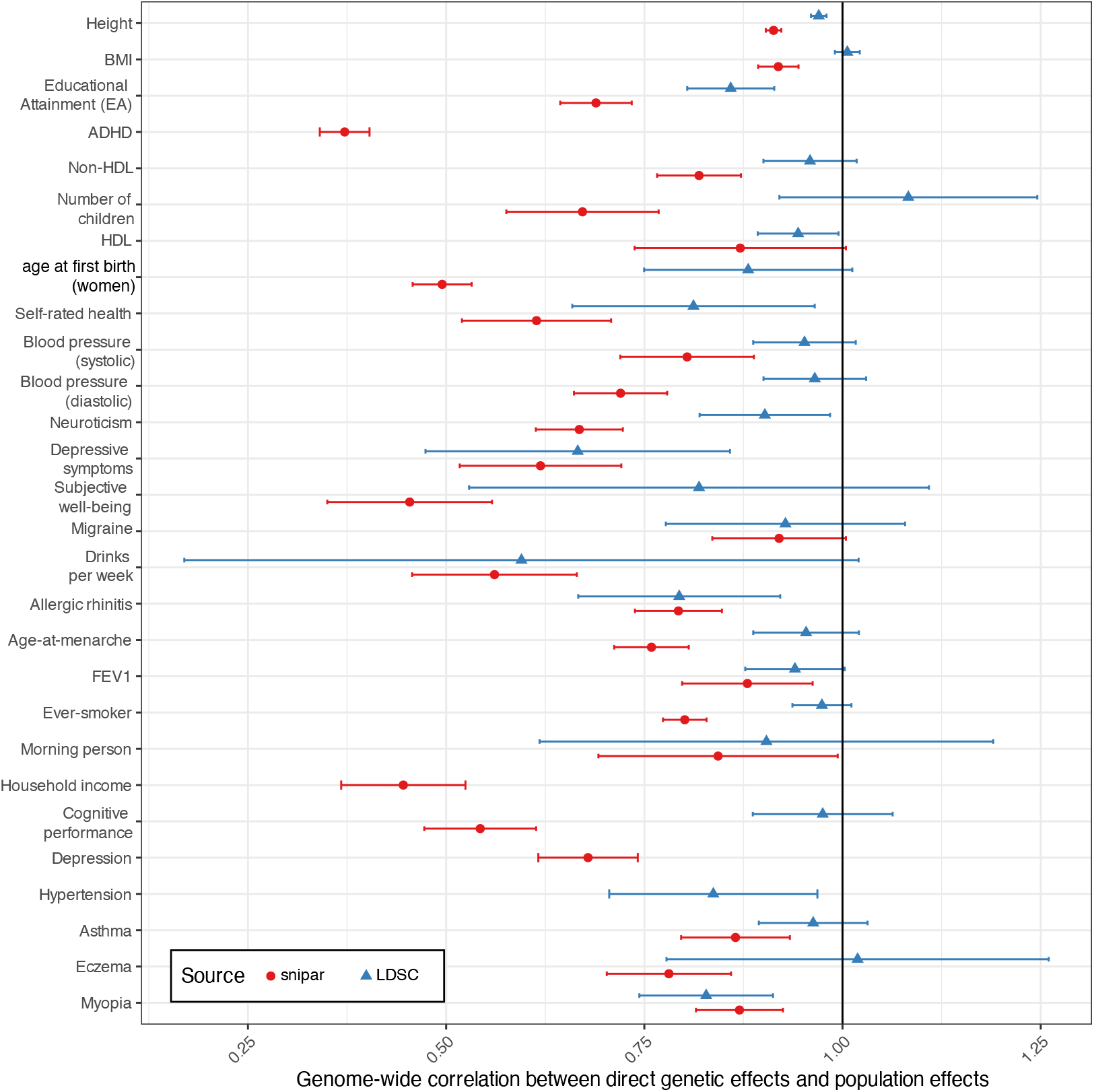
Genome-wide correlations between direct genetic effects (DGEs) and population effects. Horizontal bars give 95% confidence intervals. The correlations as estimated by snipar measure the genome-wide correlation between marginal effects, whereas, by adjusting for local LD, the correlations estimated by LDSC aim to measure the genome-wide correlation between the joint-fit effects while adjusting for population stratification. The correlations estimated by snipar thus give a better measure of how different genome-wide summary statistics on DGEs and population effects would be in the absence of sampling error, whereas LDSC gives a better measure of how correlated DGEs and population effects would be after adjusting for sampling error, local LD, and some component of population stratification. Differences between the two estimates can therefore be informative about the contribution of population stratification to confounding in GWAS, with higher estimates from LDSC suggesting a contribution from population stratification. Abbreviations: HDL, high density lipoprotein cholesterol; FEV1, forced expiratory volume in 1 second adjusted for height; BMI, body mass index. See Table 1 for numerical values.

We investigated whether the correlation between DGEs and population effects differed between the cohort level and the meta-analysis level by performing a random-effects meta-analysis of cohort-level estimates of the correlation between DGEs and population effects (Supplementary Figure 4). For EA, the meta-analysis estimate of the cohort-level correlations was 0.547 (S.E.=0.052), lower than from the meta-analysis summary statistics, 0.689 (S.E.=0.023). Similar patterns were observed for other phenotypes (Supplementary Table 3). A plausible explanation for this is that confounding at the variant level varies somewhat randomly across cohorts and is thus partially cancelled out in meta-analysis estimates of population effects, leading to increased correlation with DGEs.

The method we used — implemented in *snipar —* estimates the correlation between the DGEs and population effects (which are marginal effects) while accounting for sampling errors. This is different from estimating genetic correlation using LD score regression (LDSC) or related techniques that attempt to measure the correlation between underlying joint-fit effects, adjusting for population stratification and local LD^5,26^. Thus, LDSC-estimated genetic correlations between DGEs and population effects will underestimate the degree of confounding in population effects to the degree that LDSC successfully adjusts out population stratification confounding. LDSC-estimated correlations between DGEs and population effects below 1 could therefore indicate the influence of confounding factors other than population stratification in standard GWAS — such as IGEs — or imperfect control for population stratification by LDSC^5^.

To gauge how much of the confounding in population effects can be attributed to population stratification as opposed to IGEs or non-random mating, we estimated the correlation between DGEs and population effects using LDSC (Figure 1 and Table 1) and compared these to the *snipar* estimates. We found that height, myopia, hypertension, allergic rhinitis, depressive symptoms, neuroticism, self-rated health, HDL cholesterol, and EA had correlations statistically significantly below 1 (FDR<0.05, one-sided test). However, most of the correlations estimated by LDSC were close to 1, indicating that much of the confounding in GWAS is likely due to uncorrected population stratification.

### Substantial contribution of confounding to GWAS population effects

To further investigate confounding, we estimated the proportion of genome-wide variance in population effects (the non-sampling variance in genome-wide population effect estimates) that is uncorrelated with DGEs (Methods, Figure 2, and Supplementary Table 3), a likely characteristic of population-stratification confounding^10,24^, although other factors could contribute, including IGEs that are weakly correlated with DGEs and/or cross-trait AM^7^. Estimates of the contribution to population effects from factors uncorrelated with DGEs were 10.7% (S.E.=0.6%) for height and 10.2% (S.E.=1.0%) for BMI but reached 48.3% (S.E.=3.2%) for EA and 58.7% (S.E.=7.0%) for depressive symptoms. These results indicate that confounding factors uncorrelated with DGEs make a relatively small but non-negligible contribution to GWAS of traits such as height and BMI but comprise the majority of population effects for some phenotypes. These results apply genome-wide, where most variants likely have very weak or zero DGEs. Thus, the relative contribution from confounding factors at strongly associated variants — such as genome-wide-significant variants — is likely much smaller than genome-wide.

**Figure 2.**
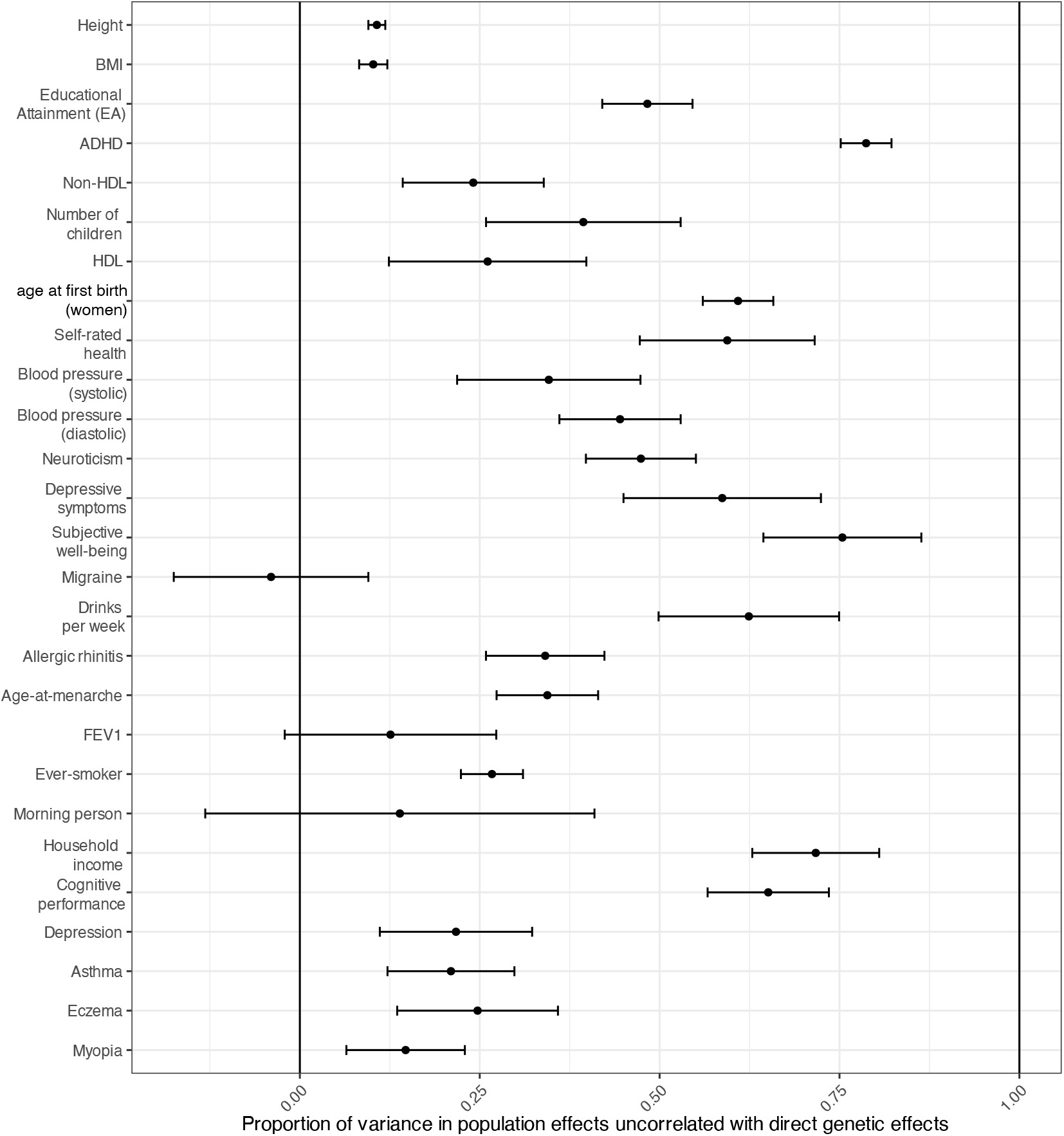
Proportion of non-sampling variance in population effects that is uncorrelated with direct genetic effects (DGEs). Horizontal bars give 95% confidence intervals. Only includes phenotypes with median effective sample size for DGEs > 5000 and SE < 0.25. This statistic is computed by performing a type of genome-wide regression of population effects on DGEs accounting for sampling errors (Methods). This gives a measure of the contribution of factors that are uncorrelated with DGEs, a likely characteristic of population stratification confounding^**10**,**24**^. Abbreviations: HDL, high density lipoprotein cholesterol; FEV1, forced expiratory volume in 1 second adjusted for height; BMI, body mass index; EA, educational attainment; Ever-smoker, whether an individual has ever smoked. See Supplementary Table 3 for numerical values.

We also analyzed the LDSC intercept, which has been proposed as a measure of confounding in GWAS test statistics, for both DGE and population effect summary statistics, finding results consistent with the removal of confounding in FGWAS estimates of DGEs (Methods and Supplementary Table 3).

### Inflation and deflation of direct genetic effects relative to population effects

We estimated the degree to which population effects are systematically inflated/deflated relative to DGEs by performing a genome-wide regression of population effects onto DGEs that accounts for sampling errors (Methods and Supplementary Figure 5). Height and systolic blood pressure have inflated population effects, whereas 10 phenotypes (ADHD, eczema, depression, cognitive performance, household income, drinks-per-week, subjective wellbeing, neuroticism, age at first birth in women, and number of children) have deflated population effects (FDR<0.05, two-sided test).

These results run counter to the intuition that FGWAS and sib-GWAS DGE estimates should be deflated relative to population effects because the influence of AM and IGEs, which are often assumed to be highly correlated with DGEs, is removed^9,23,27^. These results also differ from those derived from the sib-GWAS by Howe et al., which reported that DGEs were smaller than population effects for number of children, depressive symptoms, EA, cognitive ability, ever-smoker, and height. However, their analysis only examined SNPs that were strongly associated (P<5×10^-8^ or P<1×10^-5^) based on standard GWAS in a UK Biobank subsample and weighted the analysis towards SNPs with stronger population effects, which may have contributed toward their analysis finding more deflation than there is genome-wide.

### Negative correlations between DGEs and NTCs due to ascertainment and selection

A phenomenon related to deflation of population effects is negative genome-wide correlation between DGEs and average NTCs, first noted by Young et al. for cognitive performance and neuroticism in the UK Biobank^3^. This is because the population effect of a SNP, *β*_*l*_, is approximately the sum of the DGE and average NTC: *β*_*l*_ ≈ *δ*_*l*_ + *α*_*l*_. So if DGEs and average NTCs are negatively correlated, they will tend to cancel each other out, resulting in deflated population effects.

We found that the correlation between DGEs and average NTCs was different from zero (FDR<0.05, two-sided test) for 8 out of 24 phenotypes (Supplementary Table 3): below zero for ADHD, allergic rhinitis, eczema, cognitive performance, and household income; and above zero for systolic blood pressure, EA, and height (Supplementary Figure 6). This is consistent with deflation of population effects for eczema, cognitive performance and household income and inflation for height and systolic blood pressure. We also estimated that population effects on EA are inflated by a factor of 1.131 (S.E.=0.072), but this estimate is not statistically distinguishable from 1.

Young et al. showed that negative correlations between DGEs and average NTCs — and concomitant deflation of population effects — could be due to collider bias induced by biased sampling with respect to phenotype values^3^. It is therefore plausible that the deflation of population effects we observed for 9 phenotypes is due in part to ascertainment bias, although natural selection (specifically, directional or stabilizing selection) may also contribute through negative LD induced between sign-concordant causal alleles, consistent with the Bulmer effect^28^. The influence of natural selection is likely strongest for age at first birth in women and number of children since these traits are directly related to evolutionary fitness^29^. Another phenomenon that could contribute is within-family contrast effects, where family members (e.g. siblings) differentiate from each other, inducing IGEs in the opposite direction to DGEs^30^.

We also estimated the correlations between DGEs and average NTCs using LDSC, which should adjust for some of the contribution of population stratification confounding to NTCs. We found statistically significant positive estimates for height and EA (Supplementary Table 3 and 6). The primary explanation is likely AM, which is strong for EA and height^25,31^, although IGEs that are positively correlated with DGEs could also contribute^3^. AM leads to inflated population effects relative to DGEs^23,25^ and average NTCs that are positively correlated with DGEs^3,23^. To see this, consider that under AM at equilibrium, 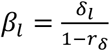, where *r* is the correlation between parents’ DGE components^23^. Thus, 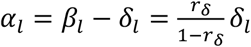.

### Smaller SNP heritability from direct genetic effects than population effects

Genotype-phenotype data on unrelated individuals can be used to estimate ‘SNP heritability’: the proportion of phenotypic variance explained by a linear model of the genotyped SNPs^32^. This is usually achieved by application of Genomic-Relatedness-Matrix Restricted Maximum Likelihood (GREML) to individual level data^32^ or by application of LDSC to GWAS summary statistics^33^. Variants of these methods have been used to investigate the genetic architecture of phenotypes^34,35^.

The definition of SNP heritability in terms of the variance explained by a linear model of genotyped SNPs differs from traditional definitions of heritability, which defined heritability as the proportion of phenotype variance explained by causal genetic effects. Thus, SNP heritability can include contributions from IGEs, population stratification (more relevant for GREML than LDSC, which attempts to adjust for population stratification), and can be inflated by AM^6^. Genome-wide summary statistics on DGEs from FGWAS can be used instead as inputs to LDSC^9^, thereby giving SNP heritability estimates that remove contributions from IGEs, population stratification, and inflation due to AM, bringing them closer to traditional definitions of heritability. However, DGE-based SNP heritability estimates do not account for the increase in genetic variance due to AM-induced correlations between causal alleles, leading to a downward bias similar to other family-based heritability estimates such as classical twin designs and relatedness disequilibrium regression^23,36^. In contrast, LDSC will overestimate SNP heritability when applied to population effects that are inflated due to AM^6^.

Table 1 and Figure 3 show SNP heritability estimated from meta-analysis estimates of DGEs and population effects. SNP heritability estimates from DGEs are smaller than from population effects (FDR<0.05, two-sided test, Methods, Supplementary Table 5) for age at first birth in women, EA, depression, whether an individual has ever smoked, and height. Since AM is strong for all of these phenotypes^31^ (AM is likely for age at first birth in women indirectly due to the correlation of age at first birth with EA^29,37^), these results suggest that larger LDSC SNP heritability estimates from population effects are primarily due to AM^6^, with the influence of population stratification diminished to the degree that LDSC successfully adjusts for it.

**Figure 3.**
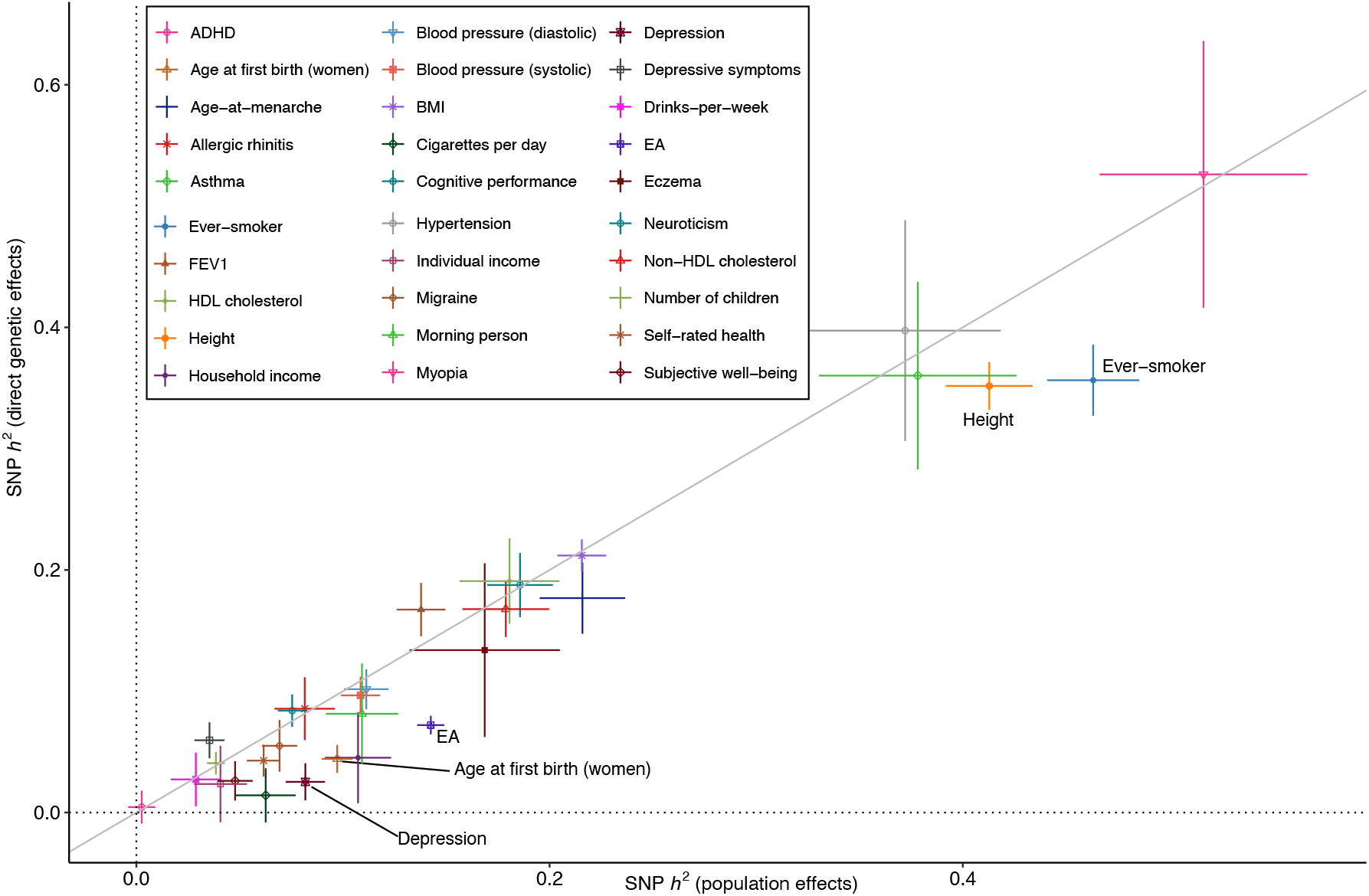
Comparison of SNP heritability estimates from direct genetic effects and population effects. The x-axis is the SNP heritability estimate from applying LDSC^**33**^ to genome-wide summary statistics on population effects. The y-axis is the SNP heritability estimate from applying LDSC to direct genetic effects (DGEs) (Methods). Vertical and horizontal error bars give the 95% confidence intervals. The diagonal line is the identity. We label the phenotypes with statistically detectable differences (FDR< 0.05, two-sided test): Age at first birth (women); EA, educational attainment; Ever-smoker, whether an individual has ever smoked; Depression; and Height.

Our SNP heritability estimates are generally similar to those from Howe et al.^9^ although with greatly increased precision for some phenotypes. However, we do not replicate the Howe et al. result for cognitive performance: we find similar SNP heritability from DGEs (0.188, S.E. 0.027) and population effects (0.186, S.E. 0.016), whereas Howe et al. found substantially smaller heritability from DGEs (0.14, S.E. 0.043) than from population effects (0.24, S.E. 0.031).

Between-cohort heterogeneity has been proposed as an explanation for lower SNP heritability estimates from meta-analysis than from individual cohorts^38^. For EA, we investigated this hypothesis. However, we found nearly identical results when using meta-analysis summary statistics as when meta-analyzing cohort-level SNP heritability estimates (Supplementary Figure 7): both indicated that the SNP heritability estimated from DGEs is around 7% compared to around 14% estimated from population effects. Our estimate of the SNP heritability from DGEs for EA, 7.2% (S.E. 0.8%), was larger than estimated by Howe et al., 4% (S.E. 0.8%).

### Differing results from China Kadoorie Biobank and European meta-analysis

We performed a similar set of analyses in the China Kadoorie Biobank (Methods). We observed results consistent with strong ascertainment bias and/or confounding due to negative gene-environment correlations (Supplementary Table 4), including higher SNP heritability from DGEs than from population effects for phenotypes including BMI, EA, and height — the opposite of the results from the European ancestry meta-analysis. Analyses of additional Chinese and East Asian genetic ancestry data will be needed to confirm whether these results are cohort specific or apply more widely to Chinese and other East Asian genetic ancestry cohorts.

### Functional enrichment analyses give similar results whether using direct genetic effects or population effects

A possible consequence of confounding in standard GWAS population effect estimates is biased estimates of functional enrichment — the degree to which genes/variants having certain functional annotations contribute more to SNP heritability than others. To investigate this, we performed a functional enrichment analysis using the same stratified LDSC^34^ analysis as in Lee et al.^27^. We analyzed both DGE and population effect estimates on EA and height from our European genetic ancestry meta-analysis, finding no discernable differences between enrichment estimates from DGE and population effects for either phenotypes (Supplementary Figures 8-9), although power for this analysis was limited for EA DGEs. This suggests that functional enrichment estimates from stratified LDSC may not be particularly susceptible to bias from confounding in standard GWAS, potentially due to adjustments for stratification made by LDSC and/or because stratification may affect all loci similarly irrespective of functional annotation.

### Robust estimation of pleiotropy using direct genetic effects

Genome-wide population effect summary statistics on two phenotypes can be input to LDSC to estimate the ‘genetic correlation’ between the phenotypes^26^, defined as the genome-wide correlation in population effects, adjusting for local LD and population stratification/sample overlap. This approach has been instrumental in the development of multi-phenotype methods such as GenomicSEM^39^, which use GWAS summary statistics to learn about the shared genetic architecture of phenotypes. However, population-effect estimates include contributions from IGEs, population stratification, and AM. This has led some to question whether genetic correlations estimated from population effects truly reflect underlying shared biology (pleiotropy) or shared IGEs/confounding in the population-effect estimates^7,8^.

To test how analyses of pleiotropy based on genetic correlations have been influenced by confounding, we applied GenomicSEM to both DGEs and population effects (Methods, Figure 4, Supplementary Tables 6-7). (We used GenomicSEM to perform a statistical test for differences in genetic correlations estimated from DGEs and population effects, which would not be possible in LDSC.) We found a general inflation of test statistics comparing genetic correlations estimated using DGEs and population effects (Supplementary Figure 10). The genetic correlation estimates were statistically different for 22 trait pairs (FDR < 0.05, two-sided test).

**Figure 4.**
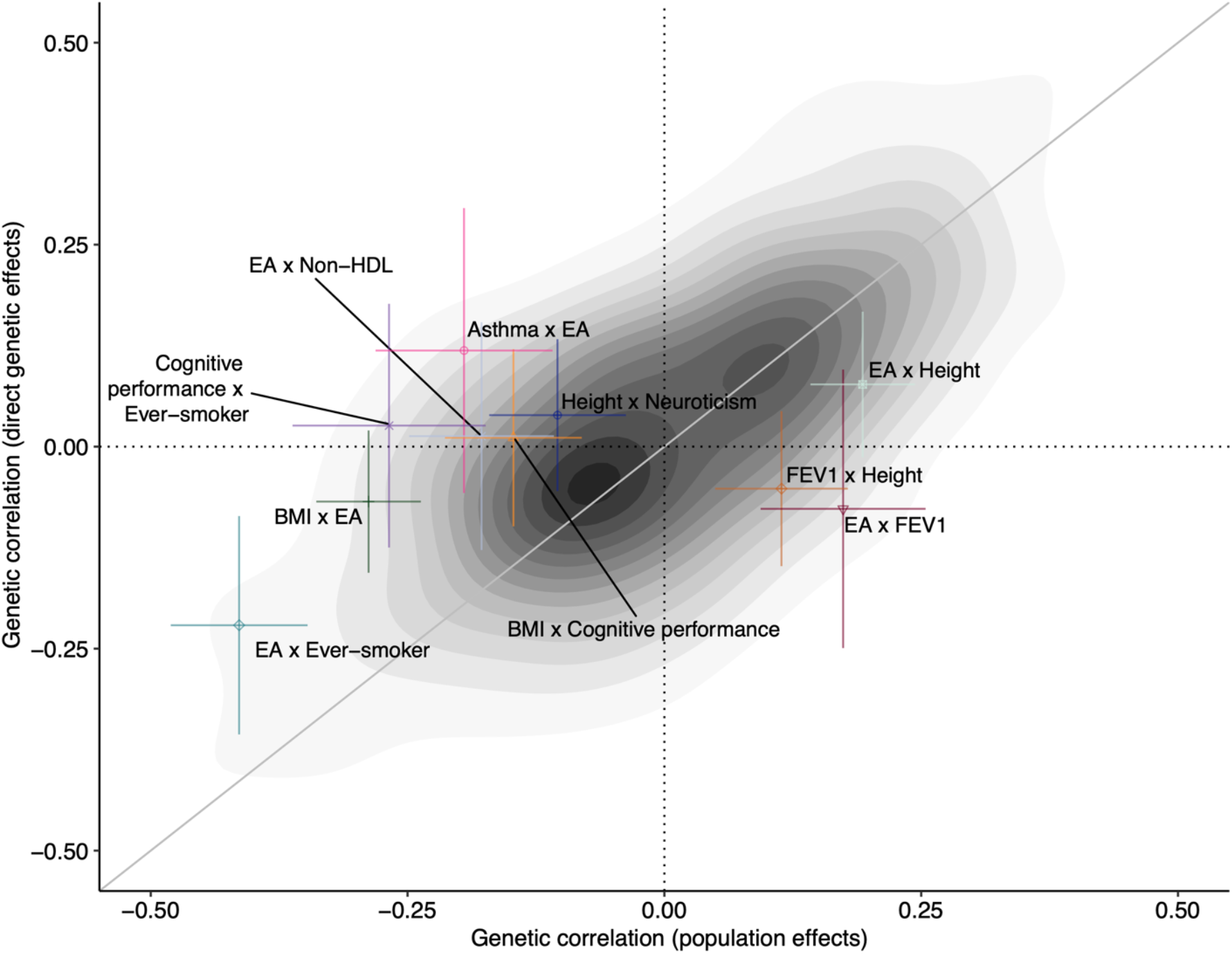
Comparison of genetic correlations estimated from population effects (x-axis) and direct genetic effects (y-axis). The shading gives the density of points from 435 pairs of phenotypes. We have marked and labeled the trait pairs where the genetic correlations are statistically distinguishable (FDR<0.01, two-sided test). The diagonal line is the identity. Errors bars indicate 95% confidence intervals. Trait abbreviations: BMI, body mass index; EA, educational attainment (years); FEV1, forced expiratory volume in 1 second; Non-HDL, total cholesterol minus high density lipoprotein cholesterol; Ever-smoker, whether an individual has ever smoked.

Some pairs of phenotypes appear to have strong genetic correlations when estimated using population effects but have genetic correlations close to zero when estimated using DGEs: for example, the genetic correlation between EA and BMI reduces in magnitude from -0.288 (S.E. 0.026) to -0.068 (S.E. 0.045). A general attenuation of genetic correlations between EA and other traits is observed when DGEs are used in place of population effects (Supplementary Figure 11), indicating that factors other than DGEs (including IGEs and AM) inflate population-effect-based estimates of genetic correlations. However, for many pairs of phenotypes, genetic correlations are similar whether estimated using DGEs or population effects. For example, there is little attenuation in genetic correlations with BMI, except for BMI-EA (Supplementary Figure 12).

We investigated the hypothesis put forward by Border et al.^7^ that cross-trait AM (xAM) has inflated the magnitude of genetic correlation estimates from (standard GWAS) population effects (Methods). xAM is expected to have a negligible influence on genetic correlations estimated using DGEs because the vast majority of correlations between causal alleles induced by xAM are cross-chromosome and therefore do not contribute to DGE estimates^14–16^. Using a Bayesian method that adjusts for sampling errors, we find that cross-mate phenotypic correlations (a measure of xAM) explain substantial variation in both population-effect genetic correlations estimates (R^2^ = 40.77%, 95% CI: 34.44% - 46.52%) and DGE correlation estimates (R^2^ = 15.37%, 95% CI: 10.41% - 20.71%). There is expected to be a relationship between cross-mate phenotype correlations and DGE genetic correlations under univariate AM when there is true pleiotropy (Methods); the fact that the relationship is stronger for population-effect genetic correlations is evidence that xAM contributes to population-effect genetic correlation estimates. Moreover, cross-mate phenotypic correlation estimates predict differences between population and DGE genetic correlations (R^2^ = 6.53%, 95% CI: 3.50% - 10.07%; Supplementary Figure 13). These results support Border et al.’s hypothesis and indicate that pleiotropy should be investigated using DGE-based genetic correlation estimates.

### PGIs based on direct genetic effects exhibit less confounding

Polygenic predictors (called polygenic indices, PGIs, or polygenic scores) based on DGE estimates from FGWAS – hereafter, DGE PGIs – have favorable properties due to the removal of confounding from PGI weights, making them suited to applications that are sensitive to confounding^4,5,9,13,23^. We examined out-of-sample prediction using PGIs derived from our meta-analysis estimates of DGEs and population effects. We used the Millennium Cohort Study^40^ (MCS) as our primary validation cohort and the UK Biobank^41^ (UKB) as a secondary validation cohort for phenotypes that are not available in MCS (Supplementary Table 8).

We performed standard and family-based PGI (FPGI) analyses using *snipar* (Methods, Figure 5, and Supplementary Table 9). In the standard analysis, we perform a regression controlling for standard covariates (age, sex, principal components), and we report the standardized coefficient on the PGI, called the ‘population effect’. Even DGE PGIs may be correlated with genetic factors not directly captured by the PGI or environmental factors, leading to confounding in standard PGI analysis^23^. In the FPGI analysis, we add parental PGIs as covariates, enabling estimation of the ‘direct effect’ of the PGI, which reflects only DGEs^15,16^.

**Figure 5.**
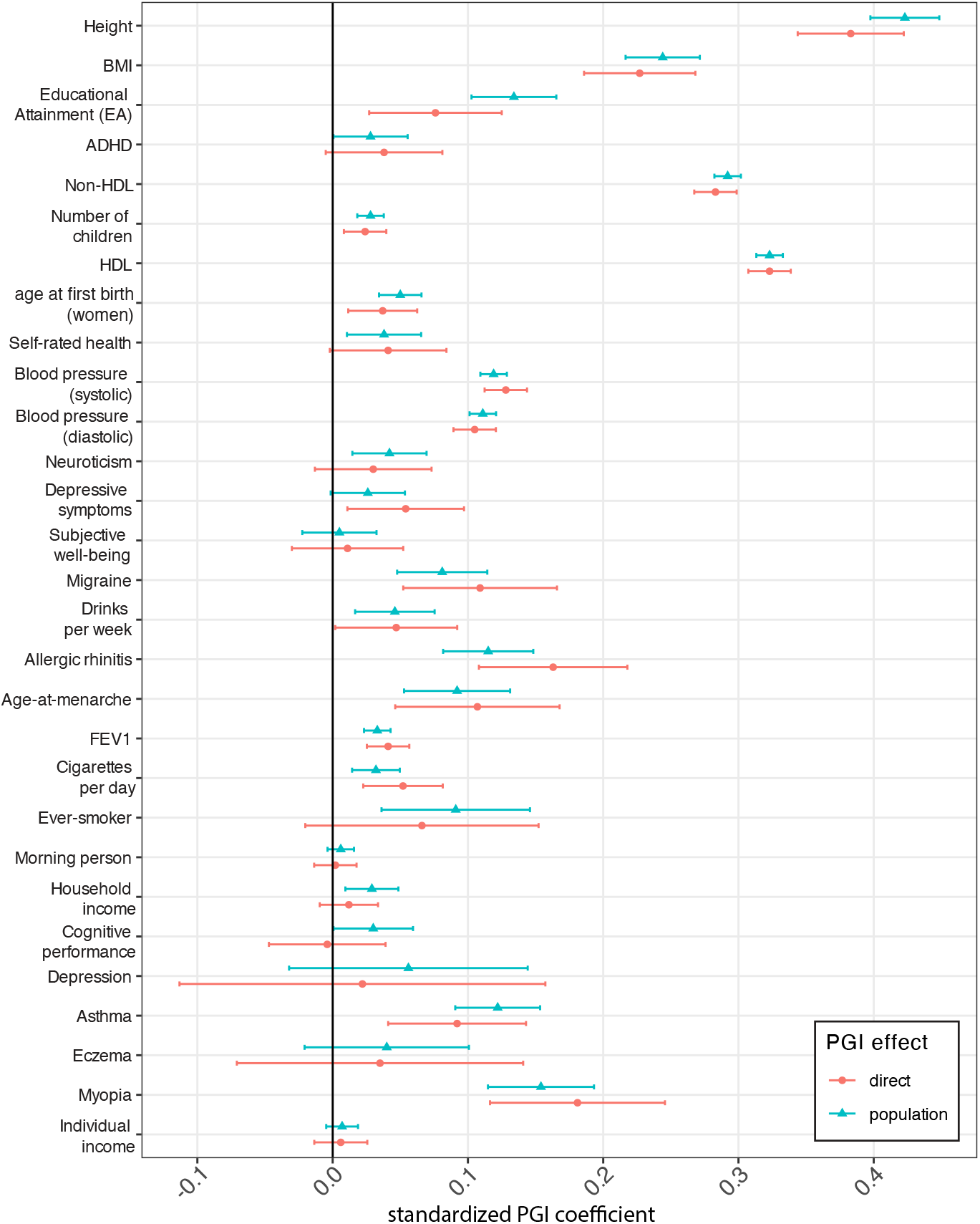
Out-of-sample polygenic prediction analyses using direct genetic effect (DGE) summary statistics. We give standardized effect estimates (for phenotype and DGE PGI normalized to 1), corresponding to partial correlation coefficients. The ‘direct effect’ of the DGE PGI is the partial correlation with the phenotype conditional on parental PGIs (using the same PGI weights) and standard covariates, thus reflecting only direct genetic effects^15,16^. The population effect of the PGI is the partial correlation conditional on standard covariates (without parental PGIs) and thus reflects DGEs, IGEs, and confounding factors. Error bars give 95% confidence intervals. For EA and cognitive performance PGIs, here we show the results on average English and Math GCSE grades and the age 17 cognitive assessment from MCS, respectively — see Figure 6 for an expanded set of outcomes for these PGIs. An expanded set of numerical results is available in Supplementary Table 9. Abbreviations: EA, educational attainment (years); BMI, body mass index; HDL, high density lipoprotein; FEV1, forced expiratory volume in 1 second; Ever-smoker, whether an individual has ever smoked; Non-HDL, total cholesterol minus HDL cholesterol.

DGE PGIs had out-of-sample predictive power statistically distinguishable from zero for 24 phenotypes (FDR < 0.05, one-sided test). The height DGE PGI achieved the highest partial *R*^2^ (17.9%, 95% C.I. 15.8%-20.1%), with DGE PGIs for diastolic and systolic blood pressure, HDL and non-HDL cholesterol, and EA all achieving partial *R*^2^ above 1% (Supplementary Table 9). The direct and population effects of the DGE PGIs are generally similar (Figure 5), indicating little attenuation of PGI predictive power within-family. However, for population-effect-based PGIs, PGI population effects were substantially larger than PGI direct effects for EA, age at first birth in women, and household income (Supplementary Figure 14).

If, in FPGI analysis, the average coefficient on the parental PGIs (average NTC of the PGI) differs from zero, this shows that the PGI’s predictive power derives from factors other than DGEs of the variants used in the PGI and those in local LD with them^15^. These include environmental factors and DGEs of other variants with which the PGI is correlated due to non-random mating^15,23^. Cognitive performance, educational attainment, allergic rhinitis, height, and household income DGE PGIs had average NTCs that show evidence of being non-zero (two-sided *P*-value < 0.05), although they were not statistically distinguishable from zero after Benjamini-Hochberg correction. We therefore found only limited evidence of confounding when using DGE PGIs, although such confounding is expected under AM and thus likely affects height and education-related DGE PGIs (when analyzing relevant outcomes)^16,23^.

We analysed an expanded set of educational and cognitive phenotypes using EA PGIs constructed from both DGEs and population effects (Figure 6, Supplementary Figure 14, and Supplementary Table 9). The population-effect-based EA PGI exhibited much greater attenuation of the PGI’s prediction power within-family (i.e. much smaller direct effect than population effect) than the DGE EA PGI. This finding is consistent with a greater contribution from confounding factors to the predictive power of (standard-GWAS derived) population-effect-based EA PGIs^17,23^. It is not consistent with the within-family attenuation of EA PGI prediction being due solely to AM: under that explanation, we would expect to see greater shrinkage from the DGE PGI than from the population-effect-based PGI (*R*^2^ = 5.3%) because the DGE PGI (*R*^2^ = 1.8%) has a smaller *R*^2^ (ref^23^).

**Figure 6.**
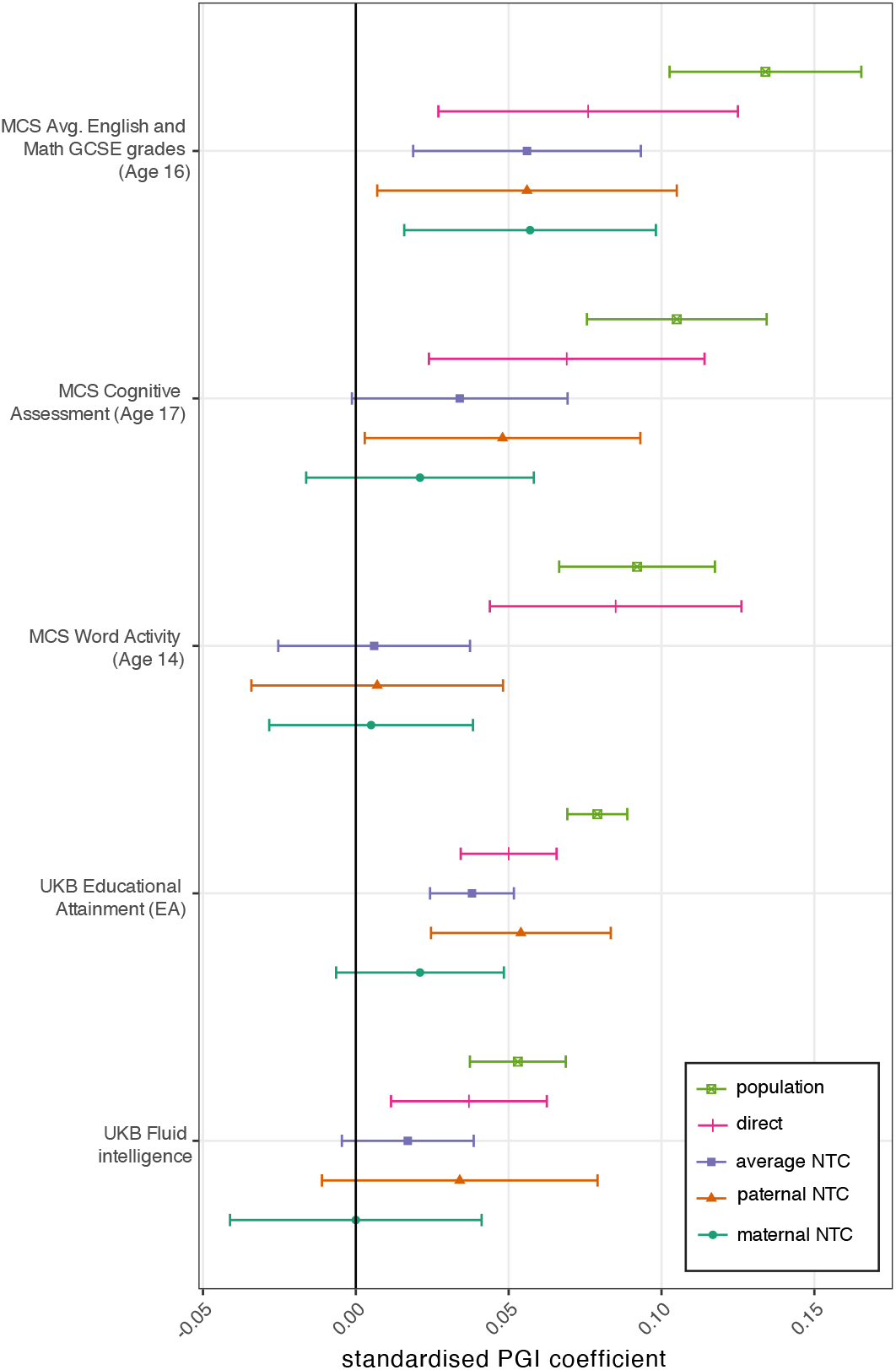
Out-of-sample polygenic prediction analysis using the educational attainment (EA) direct genetic effect (DGE) PGI. Family-based PGI analysis was performed on education and cognitive-performance-related outcomes. Error bars give 95% confidence intervals. Outcome phenotypes: Avg. Eng. & Math GCSE Score (Supplementary Note Section 4); educational attainment outcome as defined in Okbay et al.^18^; word Activity score from MCS Sweep 6 (age 14); cognitive assessment outcome from MCS Sweep 7 (age 17); fluid intelligence score from UK Biobank. Full descriptions of outcome phenotypes can be found in Supplementary Table 8. An expanded set of numerical results is available in Supplementary Table 9.

## Discussion

We presented results from a meta-analysis of FGWAS results on 34 phenotypes from 16 cohorts of European genetic ancestries and one cohort of East Asian genetic ancestry. Our study provides a template for performing FGWAS meta-analysis. By imputing missing parental genotypes^3^, we were able to maximize power while using the same analytical framework for different data types (e.g. sibling pairs, parent-offspring pairs, etc.). Meta-analysis can be performed using multivariate fixed-effects meta-analysis (Methods), resulting in meta-analysis estimates of all the parameters of interest (DGEs, NTCs, and population effects) along with their joint sampling variance-covariance matrix, facilitating downstream analyses. Our analyses showed the value of FGWAS in debiasing inferences drawn from GWAS and in investigating the impact of gene-environment correlation and non-random mating.

By estimating the genome-wide correlation between direct and population effects^3^ (Table 1 and Figure 1), we found that DGEs and population effects have correlations below one for 26 phenotypes, with quite low correlations (<0.75) estimated for 13 phenotypes including diastolic blood pressure, neuroticism, and depression — phenotypes not closely related to education or cognitive ability, the only previous phenotypes shown to have correlations below one.

Using novel methodologies for comparing DGEs, NTCs, and population effects, we show that the low correlation between DGEs and population effects is due to confounding factors in population effects (as estimated from standard GWAS) that are uncorrelated with DGEs, likely uncorrected population stratification (Methods and Figure 2). Our population effect estimates were adjusted for genetic principal components, implying that substantial residual confounding can persist after principal-component adjustment. This may be because the principal components derived from common variants do not effectively capture recent structure in the population^10^.

While the contribution from population stratification can be quite large relative to DGEs genome-wide — where most variants do not have true DGEs — the relative contribution is likely smaller for strongly associated variants, such as those that reach genome-wide significance in standard GWAS. Standard GWAS remains the most powerful study design for discovering variants robustly associated with human traits and diseases, and our results do not imply that strongly associated loci discovered and replicated by GWAS are false positives. Moreover, our analyses were restricted to common variants (minor allele frequency >1%), so our conclusions may not apply to GWAS of rare variants.

Confounding may be more likely to affect methods that use genome-wide summary statistics than only strongly associated loci. Some, but not all, of these methods have been shown to be particularly vulnerable to biased inferences due to uncorrected population stratification confounding: for example, measuring the strength of AM^23^, assessing evidence for IGEs^11,42,43^, assessing evidence for polygenic selection using ancient and modern DNA^4,5,9^, and Mendelian Randomisation^13^. For these applications, DGE estimates from sib-GWAS and FGWAS should be preferred over population effect estimates.

Inferences drawn from the application of LD score regression (LDSC) to GWAS results appear to be fairly robust to confounding due to population stratification, at least some of which is adjusted out by LDSC^5,33^. This includes functional enrichment analyses using stratified LDSC, which did not display obvious differences when applied to DGEs and population effects (Supplementary Figures 8-9). However, we found that LDSC SNP heritability estimates from DGEs were lower than from population effects for five phenotypes, including depression, for which SNP heritability was estimated at 2.5% (S.E. 1.5%) when using DGEs and at 8.2% (S.E. 1.0%) when using population effects. Since all the phenotypes displaying statistically detectable differences in SNP heritability are known to be affected by AM^31^, AM (rather than population stratification) is likely to be the primary explanation^6^, although IGEs could also contribute^44^.

Genetic correlation estimates from LDSC have been used to investigate pleiotropy and are used as inputs to multi-phenotype methods such as GenomicSEM. We demonstrated that genetic correlation estimates from population effects are different from those from DGEs for 22 pairs of phenotypes, with some pairs displaying qualitatively different estimates. We found evidence supporting the hypothesis put forward by Border et al.^7^ that genetic correlation estimates have been inflated by cross-trait AM, but other factors may also contribute, such as IGEs. Our results argue for the use of DGE estimates when investigating pleiotropy.

When we performed family-based analysis of PGIs constructed from meta-analysis estimates of both DGEs and population effects, we found less attenuation of the predictive power of DGE-based PGIs within-family than for population-effect-based PGIs. Our summary statistics enabled construction of DGE PGIs whose out-of-sample predictive power is statistically distinguishable from zero for 24 phenotypes. These summary statistics will enable downstream analyses that are sensitive to confounding.

Although our meta-analysis provides DGE summary statistics that are powerful enough for many analyses, the effective sample size for DGEs (on the order of 10^4^ to 10^5^) remains an order of magnitude lower than for the most powerful GWAS meta-analyses, which have sample sizes in the millions^17,45^. Therefore, GWAS-derived population-effect PGIs will likely provide greater out-of-sample prediction ability than DGE PGIs in the near-term. However, the confounding present in GWAS population effects means that population-effect estimates will not converge to DGEs, implying that FGWAS will produce estimates of DGEs with a smaller total error (bias plus sampling error) than standard-GWAS-derived population-effects once FGWAS effective sample sizes pass some threshold. While some of the confounding in GWAS may contribute to out-of-sample prediction ability in contexts similar to the original GWAS, such confounding may reduce prediction ability in other contexts, such as predicting across ancestries and within families^8,20^. Thus, DGE PGIs, or hybrid PGIs combining GWAS and FGWAS results, may provide improved out-of-sample prediction over population-effect PGIs in certain contexts long before FGWAS effective sample sizes approach current large-scale GWAS sample sizes.

The predominance of GWAS in human genetics has led to study designs that prioritize sampling the maximum number of unrelated individuals. This sampling strategy maximizes power to discover genotype-phenotype associations but often results in datasets without many first-degree relative pairs. While more powerful analytical approaches are being developed^20^, building family-based sampling into the design of future biobanks is crucial for realizing the potential of FGWAS and related methods. Furthermore, FGWAS and related methods present an opportunity to analyze genetically diverse samples in a way that is not susceptible to population stratification confounding^20^, but, to realize this potential, family-based sampling should be built into future efforts to diversify human genetics data.

## Supporting information

Supplementary File 2 Finngen Banner Authors

Supplementary File 1 Analysis Plan

## Data Availability

Meta-analysis summary statistics will be made available from the SSGAC data portal by the time of publication.

https://thessgac.com/

## Code availability

Cohort-level imputation of missing parental genotypes and family-based GWAS analyses were performed using *snipar*, which is freely available here (https://github.com/AlexTISYoung/snipar) with documentation here (https://snipar.readthedocs.io/en/latest/guide.html). Relationships between effects were estimated using the *correlate*.*py* script in *snipar*, and family-based PGI computation and analyses were performed using the *pgs*.*py* script in *snipar*. The code for performing the quality control and meta-analysis and other ancillary analyses is available as a git repository here: https://github.com/JonJala/within_family_project. SNP heritability and genetic correlation analyses were performed using LDSC v1.0.0 and GenomicSEM v0.0.5. Code for performing cross-trait assortative mating analyses is available here: https://github.com/rborder/FGWAS_meta_xAM.

## Acknowledgements

The study was supported by Open Philanthropy and the National Institute on Aging/National Institutes of Health through grants R24-AG065184, R01-AG042568, R01-AG083379 (to the University of California, Los Angeles) and R00-AG062787 (to the University of Southern California). This research has been conducted using the UK Biobank Resource under Application Number 11425. See Supplementary Note Section 5 for additional acknowledgements.

## Author Contributions

A.S.Y., D.J.B., A.O., P.T., and D.C. conceived and designed the study.

E.A., R.A., B.O.Å., D.I.B., B.B., A.C., C.F.C., Z.C., E.D.G., A.H., J.K., J.J., L.L., R.K.L., N.G.M., M.McGue, S.E.M., S.O., K.R., H.S., R.G.W., E.Y., and P.M.V. contributed to the Cohort Study Design & Management.

R.A., D.I.B., B.B., A.C., E.D.G., L.H., J.H., K.H., J.K., A.L., A.L., M.L., N.G.M., M.McGue, S.E.M., S.O., T.P., R.B.P., A.R., K.R., K.S., T.T., and E.Y. contributed to Cohort Data Collection.

B.B., E.A.E., A.H., C.H., J.K., K.L., R.K.L., N.G.M., M.McGue., S.E.M., M.T.O., and R.G.W. contributed to Cohort Genotyping.

R.A., L.B., D.I.B., B.B., A.C., R.C., E.D.G., L.H., A.F.H., J.H., M.A.H., A.L., M.L., K.L., R.K.L., M.McGue., B.L.M., T.P., R.B.P., A.R., K.R., K.S., E.A.W., B.S.W., R.B., A.S.Y., J.G, and M.B. contributed to Cohort Phenotype Preparation A.S.Y., J.G., T.P., B.L.M., L.B., J.H., R.A., R.K.L., A.G., K.R., A.G.E., R.C., S.B., G.N. led cohort-level FGWAS analyses.

A.S.Y, J.G, S.M.N, M.McGue., R.A., S.B., L.B., B.B., R.C., A.G.E., A.G., A.F.H., C.H., J.H., M.A.H., K.L., R.K.L., B.L.M., G.N., I.M.N., T.P., R.B.P., J.S., B.J.V., E.A.W., L.Y., P.M.V., E.M.T., and R.B. contributed to Cohort Level Data Analysis.

T.Tan. and H.J. performed quality control and meta-analysis.

A.S.Y. developed the multivariate meta-analysis method and downstream analyses methods implemented in *snipar*.

T.Tan. analysed the meta-analysis summary statistics.

E.M.T. supervised GenomicSEM analyses.

T.Tan. performed out-of-sample polygenic prediction analyses with assistance from J.G. and M.B.

M.Mir and T.Tan. performed the analysis of the effect of imputation quality on sibling genotype relationships.

R.B. performed cross-trait assortative mating analyses.

A.S.Y., T.Tan., H.J., D.J.B., A.O., P.T., R.B., E.M.T., A.K., P.M.V., L.Y., J.F., J.J.L., and M.Mir wrote the manuscript.

A.S.Y., D.J.B., A.O., and P.T. jointly supervised research.

## Competing interests

A.S.Y. is an advisor to and holds equity in Herasight, Inc. The remaining authors declare no competing interests. Following the disclosure of A.S.Y.’s financial interest in Herasight Inc. on July 30, 2025, previous co-authors from the University of Oslo (E.Y., A.H., R.C.) and the Norwegian University of Science and Technology (B.Å., K.H., A.L., L.B., A.H., B.W., B.B.) withdrew from this study, and E.Y. and B.B. withdrew the summary statistics from the MoBa and HUNT cohorts, respectively

## Methods

### Cohort level analyses

Genome-wide associations studies (GWASs) have discovered thousands of associations between genetic variants and human traits^46^. GWAS proceeds by performing a regression of the form:

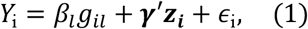

where *Y*_*i*_ and *g*_*il*_ are, respectively, the phenotype and genotype of individual *i* at SNP *l*, and ***z***_***i***_ is a vector of covariates. The GWAS parameter *β*_*l*_ is called the ‘population effect’ — as it reflects the genotype-phenotype association in the population, conditional on covariates — estimates of which are used as input to downstream analyses. Family-based GWAS (FGWAS) is defined by the regression:

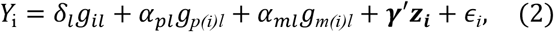

where *g*_*p(i)l*_ and *g*_*m(i)l*_ are the genotypes of individual *i*’s father and mother.

We distributed an analysis plan to each cohort (Supplementary File 1). In addition to the 30 phenotypes specified in the analysis plan, we analyzed chronic obstructive pulmonary disease (COPD), hypertension, and alcohol use disorder in Finngen. The analysis plan gave the cohort-level genotype and phenotype quality control steps. Missing parental genotypes were imputed in each cohort using *snipar*, with some cohorts using phased data to perform the imputation (Supplementary Table 1). Samples were restricted to homogeneous European ancestry subsamples for the 16 cohorts of predominantly European ancestries with the exact procedure varying depending upon cohort. Phenotypes were adjusted for standard covariates: age, sex, and genetic principal components. Following imputation of missing parental genotypes, FGWAS was performed using *snipar*. For samples without genotyped parents (Finnish Twin Cohort, Swedish Twin Register, Minnesota Twins, iPSYCH), FGWAS was performed using the imputed sum of parental genotypes^3^; the remaining cohorts used model (2) with parental genotypes replaced with their imputed values when missing. Summary statistics provided by each cohort were passed through a quality control pipeline, described in Supplementary Note Section 1, before meta-analysis was performed.

### Meta-analysis

For each variant *l*, we produced meta-analysis estimates of the parameter vector *θ*_*l*_ ≔ [*δ*_*l*_, *α*_*pl*_, *α*_*ml*_]^*T*^, where *δ*_*l*_ is the direct effect of SNP *l*, and *α*_*pl*_ and *α*_*ml*_ are the paternal and maternal non-transmitted coefficients (NTCs) (see model (2)). However, the parameter vector is not identifiable in cohorts without genotyped parents, where the imputed sum of parental genotypes is used rather than separate genotypes for each parent, as in model (2). For these cohorts, we obtained an estimate of the collapsed parameter vector, [*δ*_*l*_, (*α*_*pl*_ + *α*_*ml*_)/2]*T*, which is a linear transformation of *θ*_*l*_.

To combine the estimates from different samples, we used a generalization of multivariate (fixed-effects) meta-analysis that allows the observation from each cohort to be a linear transformation of the underlying parameter vector. Consider *z*_*jl*_ ∼ *N*(*A*_*j*_*θ*_*l*_, ∑_*jl*_), where *z*_*jl*_ is the estimated parameter vector for variant *l* from cohort *j* = 1, 2, …, *J*, and *A*_*j*_ is the matrix that gives the linear transformation that relates the underlying parameter vector to the parameter vector estimated in cohort *j*. Provided that the combination of estimates enables identifiability of *θ*_*l*_, the maximum likelihood estimate (MLE) of *θ*_*l*_ is given by:

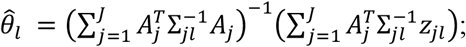

and the variance of the MLE is given by 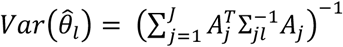. From the meta-analysis estimate of *θ*_*l*_, we can derive meta-analysis estimates of the population effect and average NTC through linear transformation. Let *α*_*l*_ = (*α*_*pl*_ + *α*_*ml*_)/2 be the average NTC. Under random-mating, the population effect, *β*_*l*_, is *β*_*l*_ = *δ*_*l*_ + *α*_*l*_. For samples with minimal structure — such as used in our meta-analysis — deviations from this relationship will be negligible^3,20^. We therefore obtained meta-analysis estimates of the expanded parameter vector through linear transformation:

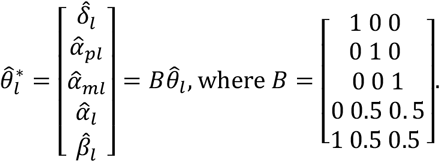

Thus, the meta-analysis estimate of the expanded parameter vector 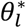 is 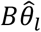 and its sampling variance-covariance matrix is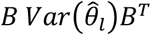.

Whenever a phenotype was available in Finngen (height, number of children, age at first birth in women, BMI, depression, ever-smoker, ADHD), we removed the Finnish cohorts that are part of FinnGen (Finnish Twin Cohort and the Botnia Family Study) from the meta-analysis and used FinnGen alone instead. We excluded educational attainment (EA) summary statistics from the Botnia Family Study due to low genetic correlation with the reference GWAS^17^.

In addition to the multivariate meta-analysis, we performed univariate meta-analysis of DGEs. We did this because multivariate meta-analysis may introduce some bias into DGE estimates due to heterogeneity in the parameter vector across cohorts. However, we found that univariate and multivariate meta-analysis estimates of DGEs were highly correlated (r=0.999) for EA (Supplementary Figure 3), suggesting that results obtained from both univariate and multivariate meta-analysis will be highly concordant. For all the results in this manuscript, we use the estimates from the multivariate meta-analysis because the joint sampling variance-covariance matrix obtained from multivariate meta-analysis facilitates downstream analyses. However, DGE summary statistics from both univariate and multivariate meta-analysis are available publicly (Data Availability).

### Estimating genome-wide relationships between effects

To estimate relationships between different types of effect, such as DGEs and population effects, we derived a moment-based estimator that accounts for the sampling errors in the estimates. For example, let 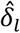 be the estimated DGE for variant *l*, and let 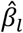 be the estimated population effect. Then we have that

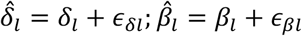

where *δ*_*l*_ is the DGE, and *ε*_*δl*_ is the sampling error; and *β*_*l*_ is the population effect, and *ε*_*βl*_ is the sampling error. The variance-covariance matrix of the sampling errors at each SNP is known from the multivariate meta-analysis (above):

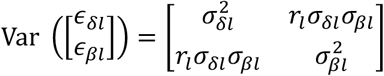

where 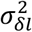 and 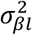are the sampling variances of the DGE and population effect estimates, and *r*_*l*_ is their sampling correlation.

The genome-wide correlations between effects and other quantities can estimated by computing the variances and covariances of the true effects. For example, we may wish to estimate the genome-wide correlation between DGEs and population effects:

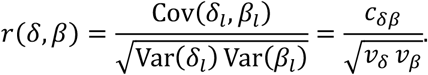

Following an approach similar to Okbay et al.^47^ (2016) (Supplement section 3.2.1.2), we assume the effects have expectation zero across the SNPs and apply the Law of Total Variance, obtaining

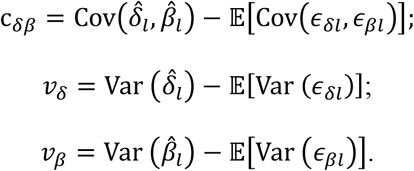

We now derive estimators for regressions of population effects on DGEs, which enable us to make inferences about inflation/deflation of population effects relative to DGEs as well as to infer the proportion of population effect variance that is due to factors uncorrelated with DGEs. Let *β*_*l*_ = *s*_*l*_ + *aδ*_*l*_, where *s*_*l*_ represents variation in population effects uncorrelated with DGEs, which could be from factors including population stratification and/or IGEs uncorrelated with DGEs. We assume that 𝔼_*l*_[*s*_*l*_] = 0 and 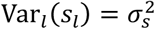. In effect, we are modeling population effects as scaled DGEs plus a random bias term with mean zero. Inflation of effects due to assortative mating would be expected to increase *a* above 1 (ref^23^). Now we consider estimating *a* and 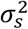 from genome-wide estimates of *δ*_*l*_ and *β*_*l*_. We estimate *a* by a noise-adjusted regression. By applying the Law of Total Variance, one can show that

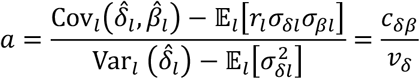

To estimate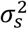, we attempt to subtract out the DGE component from *β*_*l*_. Let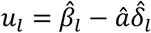, then

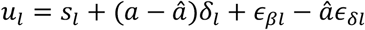

Consider the variance of *u*_*l*_:

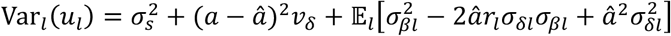

The issue here is that, since we do not know the true value of *a*, we cannot apply this formula exactly to estimate 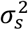. However, we can approximate (*a* − â)^2^ with its expectation: 𝔼[(*a* − â) ^2^] = Var (â). Therefore,

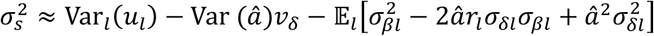

We derive weighted sample estimators for these moments in Supplementary Note Section 2.

### LDSC Intercept Analyses

The LDSC intercept has been proposed as a measure of confounding, with deviations above one argued to represent the extent of spurious inflation in GWAS test statistics^33^. We found that 29 phenotypes had intercepts detectably above one (FDR<0.05, one-sided test) when using population-effect summary statistics (Supplementary Table 3). Using DGE summary statistics, we found 15 phenotypes with intercepts detectably above one (FDR<0.05, one-sided test). Beyond uncorrected-for population stratification, other phenomena may lead to LDSC intercepts above one, including: violation of LDSC assumptions about genetic architecture^48^, differences in LD between the reference and meta-analysis samples, and cryptic relatedness^9^. In cases of a large product of sample size and SNP heritability, a normalization of the LDSC intercept called the ‘ratio’ has been proposed as a more appropriate measure of confounding^49^. Of the 15 phenotypes with DGE-based intercepts significantly greater than one, height, BMI, and ever-smoker showed small DGE-based ratios (<0.15). Nine of the other phenotypes either failed our sample-size filter for the DGE meta-analysis or showed *post hoc* a product of DGE sample size and heritability in below those excluded *a priori* for sample size only. The remaining phenotypes (age at menarche, myopia, allergic rhinitis) showed large ratios (>0.25), potentially indicating unusual genetic architectures. Overall, the 18 phenotypes with an acceptable product of DGE sample size and SNP heritability showed an average DGE-based LDSC ratio of 0.104, smaller than their average population-based ratio of 0.306—consistent with a successful removal of confounding bias.

### SNP heritability estimation

Using LDSC, we estimated ‘SNP heritability’ using meta-analysis estimates of DGEs, average NTCs, and population effects for each phenotype. To reduce the impact of variants with low precision estimates, we filtered out variants with effective sample size less than 0.8 of the median effective sample size for DGEs. Since LDSC was designed for GWAS summary statistics on population effects derived from samples of unrelated individuals, we use the effective sample size for the required sample size input^3,9^.

For an element *γ*_*l*_ of the parameter vector, the effective sample size is the sample size that would give a regression coefficient with sampling variance equal to 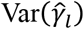 from a regression of phenotype onto genotype in unrelated individuals. We calculate as 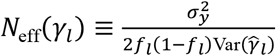 the effective sample size for *γ*_*l*_, where *f*_*l*_ is the meta-analysis allele frequency for variant *l*, and 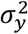 is the phenotypic variance.

For the European genetic ancestry meta-analysis, we used GenomicSEM^39^ to calculate P-values for the difference between SNP heritability estimated using DGEs and population effects. GenomicSEM allows users to fit structural equation models based on GWAS summary statistics. We used GenomicSEM software to run multivariable LD-score regression on the DGE and population effect summary statistics for each phenotype. For the LD reference panel, we used the EUR LD scores provided with LDSC^33^.

The GenomicSEM output includes the sampling variance-covariance matrix of the SNP heritability estimates from DGEs and population effects. This allowed us to calculate the P-value for the difference between SNP heritability estimated from DGEs and population effects (Supplementary Table 5).

### China Kadoorie Biobank analysis

In addition to the meta-analysis of 16 European ancestry cohorts, we conducted analyses on summary statistics from the China Kadoorie Biobank (CKB) for 15 phenotypes (Supplementary Table 4). To perform quality control on these summary statistics, we used EAS allele frequencies from 1000 Genomes^50^. For the analyses involving LDSC and *snipar*, we used an LD reference panel generated using *snipar* from the CKB data. To compare our results to a reference GWAS, we used summary statistics from Sakaue et al.^51^ (Supplementary Table 10). However, these were only available for four of the phenotypes in our analysis: BMI, height, blood pressure (systolic), and blood pressure (diastolic).

### Estimating cross-trait genetic correlations

For the European genetic ancestry meta-analysis, we used the multivariable LDSC function within GenomicSEM software to estimate the genetic covariance matrix for each pair of phenotypes and the associated sampling covariance matrix — which indexes the estimation errors in the genetic variances and covariances, along with their interdependencies. We note that while the multivariable LDSC function within GenomicSEM is capable of producing a standardized genetic covariance matrix (S_Stand) that is equivalent to a genetic correlation matrix, the associated standard errors (contained within the diagonal of V_stand) do not correspond to the standard errors of the genetic correlations, but instead correspond to standard errors of the genetic covariances that have been rescaled to the standardized metric — i.e. ignoring uncertainty in genetic variance estimates. We specified a model within GenomicSEM software to directly estimate the genetic correlation matrices with their appropriate standard errors from the unstandardized genetic covariance matrix (S) and its associated sampling covariance matrix (V). We calculated the genetic correlations between phenotypes using both DGE and population effect estimates (Supplementary Table 6), where we used the EUR LD scores provided by LDSC and the effective sample sizes (referred to in the GenomicSEM documentation as N_hat) as inputs.

We passed the output from the multivariable LD-score regression through a user-specified model within Genomic SEM in which the DGE and population effect summary statistics for each of the two phenotypes is affected exclusively by separate latent variables with fixed variances of 1.0. The freely estimated loading on the latent variable is equal to the square root of its SNP heritability. (As square roots have both positive and negative solutions, we restrict the model to positive solutions for interpretability.) By allowing the latent variables for each phenotype to covary, we obtain estimates of the genetic correlation based on both DGE and population effect estimates, and directly obtain their standard errors.

To compute the statistical significance of differences between genetic correlations estimated using DGEs and population effects, we fit a follow-up model within the GenomicSEM software in which we constrain the values of the genetic correlations based on DGEs and population effects to be equal. The estimation of a single joint parameter, rather than two separate parameters, to represent the genetic correlation for both DGE and population effects, reduces the degrees of freedom by 1, and the chi-square statistic for this model and associated P-value index the extent of violation of this equality assumption. In some cases, this procedure yielded correlations outside of [-1, 1], leading to convergence errors. When this occurred, we reran the model, constraining the relevant estimates to fall within [-0.9999, 0.9999].

### Investigation of the relationship between cross-trait assortative mating and genetic correlations

Following the procedure of Border et al.^7^, we performed mate identification in the “white British” subsample [field 22006] of the UK Biobank^41^. Among those in the analysis sample, we first selected sex-discordant pairs of unrelated individuals who reported living with their spouse [field 709], had the same values for distance to coast [field 24508], inverse distance to nearest road [field 24010], nearest distance to nearest major road [field 24012], and household size (field 790), and were concordant on whether their property was rented versus owned [field 780]. Ambiguous cases (i.e., when three or more participants matched on all criteria) were discarded. This resulted in 39,710 putative mate pairs that were used to measure cross-mate cross-phenotype correlations.

To compute cross-mate cross-phenotype correlations, we first Winsorized all continuous phenotypes at the.005 and.995 empirical quantiles separately within sex. As mates are typically similar in age, we sought to mitigate any potential inflation of cross-mate correlations due to cohort effects. To achieve this, we estimated residual Pearson, polychoric, or polyserial correlations across mates after regressing out age from the phenotype (or the latent continuous phenotype in the case of binary phenotypes). For simplicity of presentation, we constrained cross-mate cross-trait correlations to be equal across sexes—e.g., for height and BMI, we constrained the cross-mate correlations for female height x male BMI and female BMI x male height to be equal. No such constraints were needed for age at first birth and age at menarche, which were only measured in females. Cross-mate correlations were estimated using *lavaan* v0.6-15 (ref^52^).

To perform a Bayesian analysis of the relationship between cross-mate cross-phenotype correlations and genetic correlations, we estimated the correlation between latent variables *x*_*ij*_, which denotes the cross-mate cross-phenotype correlations for phenotypes *i* and *j*, and *y*_*ij*_, which denotes the population genetic correlation, the DGE genetic correlation, or the difference between them. Each of these correlations are estimated with error, so we model the estimated quantities as

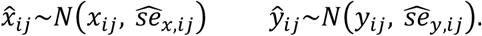

Here, 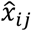 and 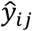 denote the corresponding estimated correlations associated with traits *i* and *j* with known standard errors, 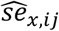and 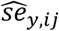. All models were fit via Hamiltonian Monte Carlo using stan v2.21.0 via the brms v2.17.0 R library^53^. For code used to fit these models, see Code Availability.

Here, we outline the rough expected relationships between cross-mate phenotype correlations and DGE and population effect based genetic correlations. If mates assort on one phenotype X that’s pleiotropic with Y (e.g. BMI and adiposity), then X and Y will be phenotypically and genetically correlated across mates, but the genetic correlation would be fully mediated through the cross-mate correlation on X. Letting Y*, X* denote the phenotypes of one’s mate, Y is correlated with X due to pleiotropy, and X is correlated with X* due to AM, which is in turn correlated with Y* due to pleiotropy. This implies that a relationship between cross-mate phenotype correlations and genetic correlations due to pleiotropy is expected even under univariate AM. Since DGE based genetic correlations should be almost entirely free from the influence of xAM, a relationship between cross-mate phenotype correlations and DGE genetic correlations can be explained by pleitropy and univariate AM, which would not be expected to generate spurious genetic correlations between phenotypes without any pleiotropy.

However, under xAM, a relationship between cross-mate phenotype correlation and population effect genetic correlations will occur even in the absence of pleiotropy, and when there is some pleiotropy, it will be overestimated^7^. This is because of bias in the population effect estimates due to xAM. To summarise, a stronger relationship between cross-mate cross-phenotype correlations and population effect genetic correlations than with DGE genetic correlations is consistent with some (but not all) of the pleiotropy signal being artifactual as we should expect in a world where lots of phenotypes are correlated across mates.

### Validation Phenotypes

We chose MCS as the primary validation cohort as it is a nationally representative sample of people born around the year 2000 in the UK. For around half of the sample, it has both parents genotyped; the other half has one parent genotyped. Validation phenotypes were chosen by finding the phenotype in MCS most similar to the phenotype on which summary statistics were collected. If such a phenotype was not available in MCS, we found the closest phenotype in UKB, and we used summary statistics from a meta-analysis excluding UKB to compute the PGIs. See Supplementary Note Section 4 and Supplementary Table 8 for further details on the validation phenotypes.

### PGI Analyses

We compute PGIs separately from DGE and population effect estimates for each phenotype. The PGI weights were computed using PRS-CS^54^. We use the EUR LD reference panel provided in PRS-CS, which was constructed using UK Biobank data and comprises 1,117,425 SNPs from HapMap3.

For the UK Biobank prediction sample, we used the subsample identified as white British by UK Biobank^41^, and for the MCS prediction sample, we used the subsample identified as closest to the EUR superpopulation cluster from 1000 Genomes, as described in Guan et al.^20^. Using *snipar*, we imputed missing parental genotypes for the samples with at least one sibling and/or parent genotyped (but without both parents genotyped), as described in Guan et al.^20^ and Supplementary Table 1.

We performed standard and family-based PGI (FPGI) analyses using *snipar*^3^ (Supplementary Table 9). For PGIs derived from both DGE estimates (DGE PGIs) and population effect estimates, we performed regressions of the form:

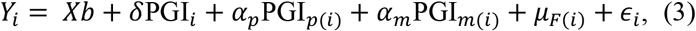

where *X* is the design matrix of the covariates (sex, a third degree polynomial in age, their interactions, and the first 20 genetic PCs); *b* is the vector of regression coefficients for the covariates; PGI_)_ is the PGI of individual *i*; PGI_*p*(*i*)_ is the paternal PGI; PGI_*m*(*i*)_ is the maternal PGI; *δ* is called the “direct effect” of the PGI^15^; *α*_*p*_ and *α*_*m*_ are the paternal and maternal non-transmitted PGI coefficients; *μ*_*F*(*i*)_ is the phenotypic mean in the family which individual *i* is in, which we modelled with a random effect^3^; and *ε*_*i*_ is the residual error. Both offspring and parental PGIs were computed using the same set of SNPs and the same weights: i.e. both used weights derived from DGE summary statistics or population effect summary statistics. When a parent was not genotyped, we used their genotypes as imputed by *snipar* to compute the PGI for that parent^3^. We estimated the ratio between the average NTC and direct effect of the PGI 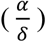 and the difference between the maternal and paternal NTCs (*α*_*m*_ − *α*_*p*_). We used the Delta Method to estimate the standard error for 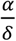.

To estimate the ‘population effect’ of the PGI, we performed a regression without controlling for parental PGIs, but using the same sample:

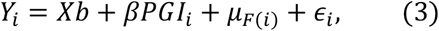

where *β* is the population effect of the PGI. Unlike in the single variant case, the population effect estimated from *β* can differ substantially from *δ* + *α* — where *α* is the average NTC for the PGI — when there is AM^3,7^. At equilibrium, *β* = *δ* + (1 + *r*_par_)*α*, where *r*_par_ is the correlation between parents’ PGIs^23^.

For binary phenotypes, we ran a similar set of regressions, but instead fit a generalized linear mixed model using the *glmer()* function in the R package *lme4*^55^.We control for sex, age, the interaction of age and sex, and the first 10 PCs, setting *nAGQ = 1* and using the *bobyqa* optimizer. For the migraine and eczema phenotypes, we use only the first 5 PCs and set nAGQ = 0 to achieve model convergence. For depression, we fit a generalized linear model controlling for age, sex, the interaction of age and sex, and the first 20 PCs, due to convergence issues with the generalized linear mixed model.

## Notes

### Competing Interest Statement

A.S.Y. is an advisor to and holds equity in Herasight, LLC.
The remaining authors declare no competing interest.

### Author Declarations

We used only previously collected data from 17 cohorts and have included links to relevant publications and ethics approvals for the contributing cohorts in Supplementary Note Section 5.

### Summary of Updates

Withdrawal of HUNT and MoBa authors.

