## Supplementary Figures for "Family-GWAS reveals effects of environment and mating on genetic associations"

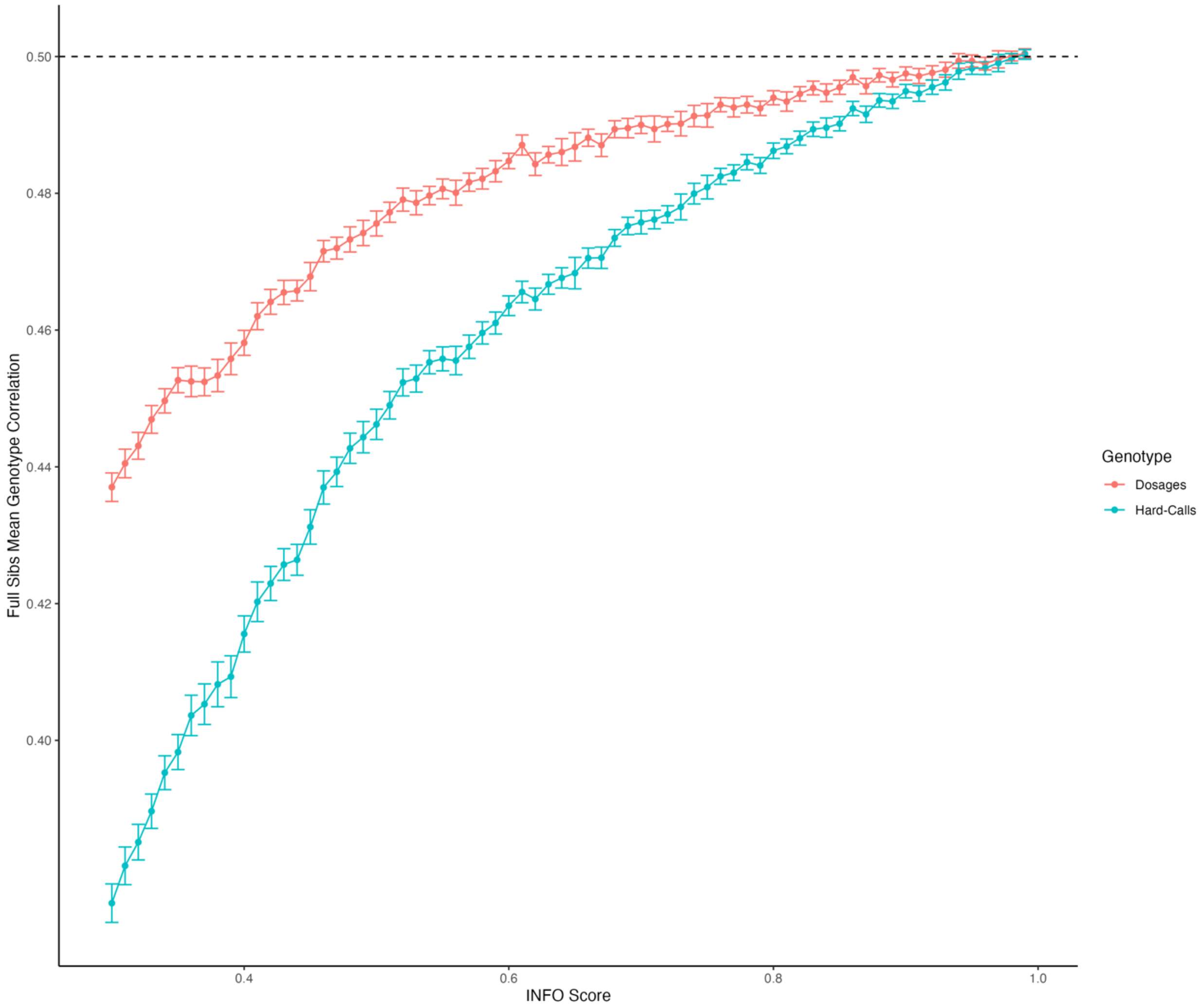


**Supplementary Figure 1. Correlations between siblings' imputed genotypes as a function of INFO score.** Using 19,290 sibling pairs from the UK Biobank white British subsample, we computed the mean correlation between sibling pairs' imputed genotypes using 1000 randomly selected SNPs with minor allele frequency (MAF)>1% from across the genome falling within the INFO score bin. We used INFO score bins from [0.30,0.31) to [0.99,1.00] in increments of 0.01. We show the mean correlation in each bin along with its 95% confidence interval for both imputed dosages (expected genotype given genotype probabilities) and imputed hard-call genotypes (most likely genotype). The sib-GWAS produced by Howe et al. used SNPs with MAF>1% and INFO score > 0.3 for their meta-analysis. See Supplementary Note Section 1 for further discussion of these results.

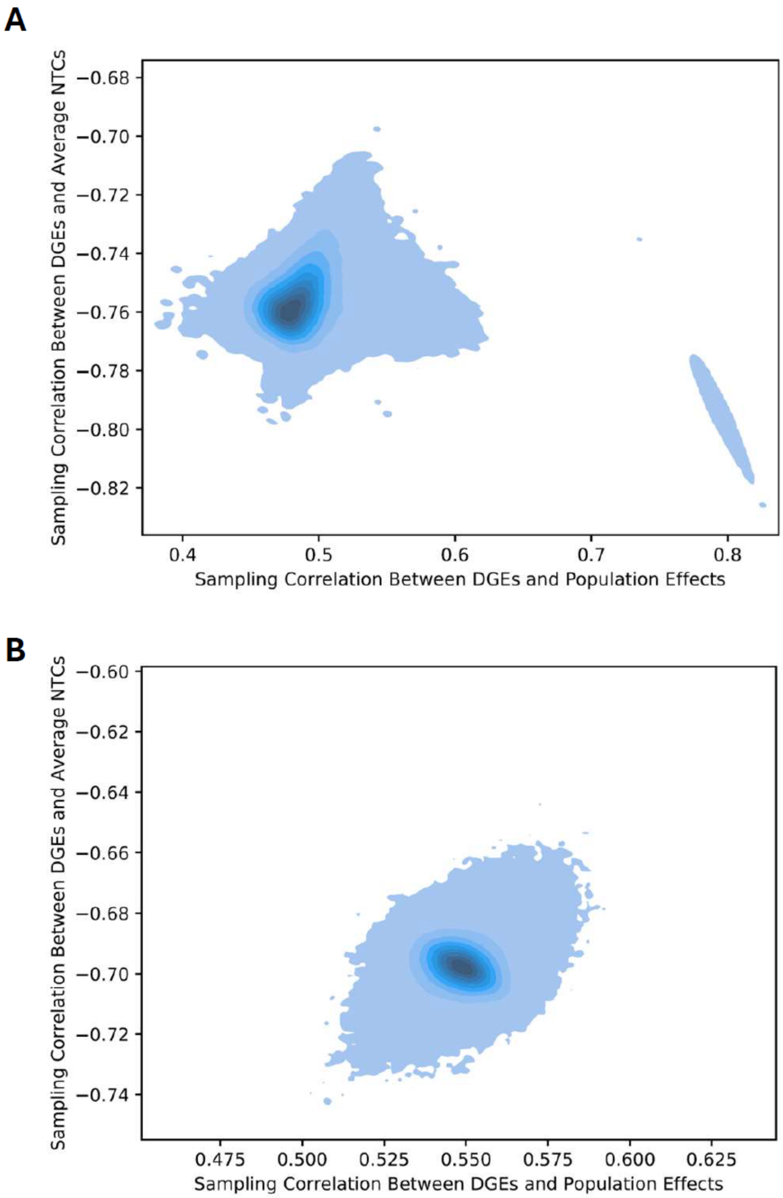


**Supplementary Figure 2. Sampling correlations between elements of parameter vector**. The x-axis plots the sampling correlation between direct genetic effects (DGEs) and population effects. The y-axis plots the sampling correlation between DGEs and average non-transmitted coefficients (NTCs). Shading indicates the density of SNPs. Panel A shows sampling correlations for BMI in Finnish Twin Cohort summary statistics; Panel B shows sampling correlations for BMI in Generation Scotland summary statistics. Variants with outlying sampling correlations are filtered out (Supplementary Note Section 1).

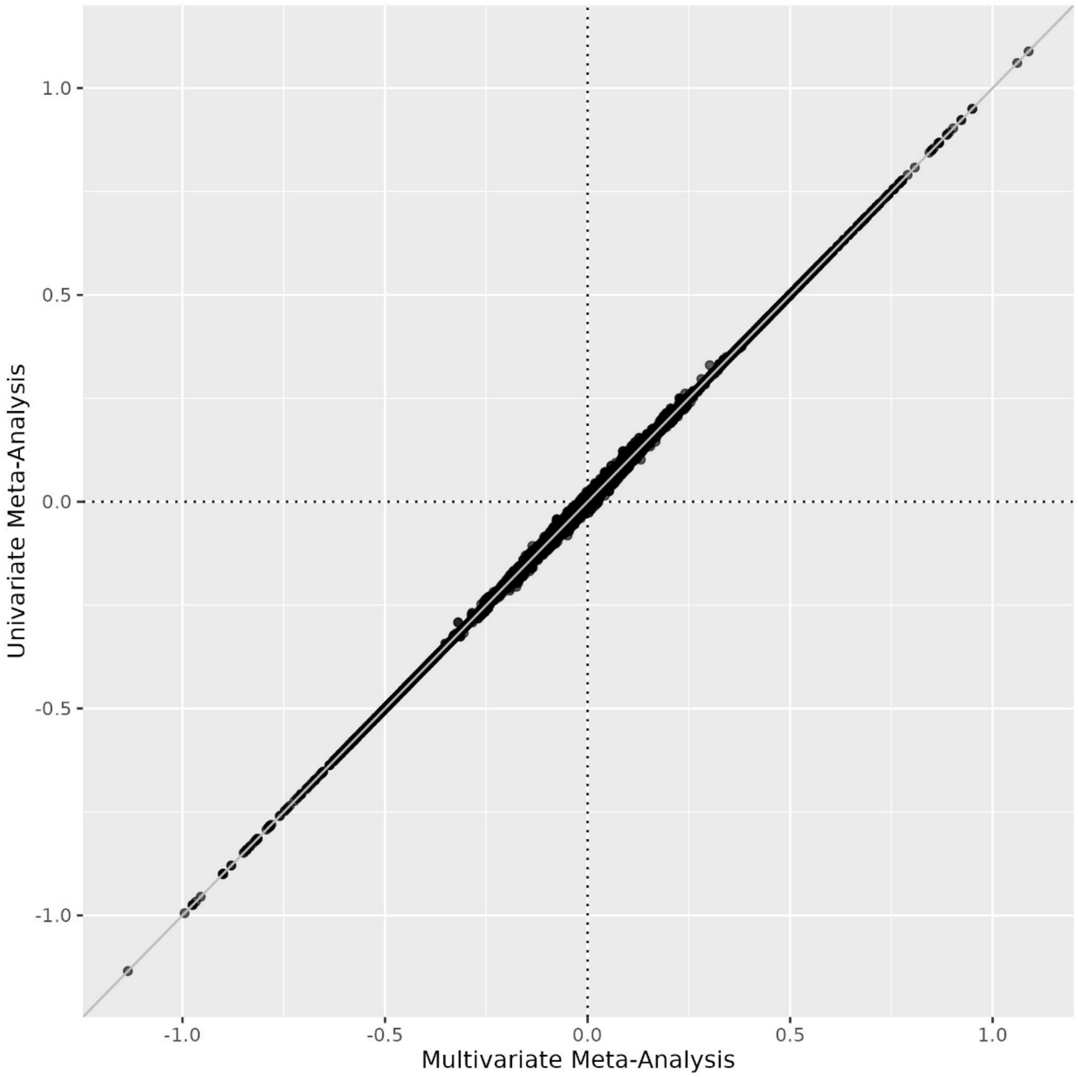


**Supplementary Figure 3. Comparison of univariate meta-analysis with multivariate meta-analysis estimates of direct genetic effects on educational attainment (EA).** The correlation is 0.999.

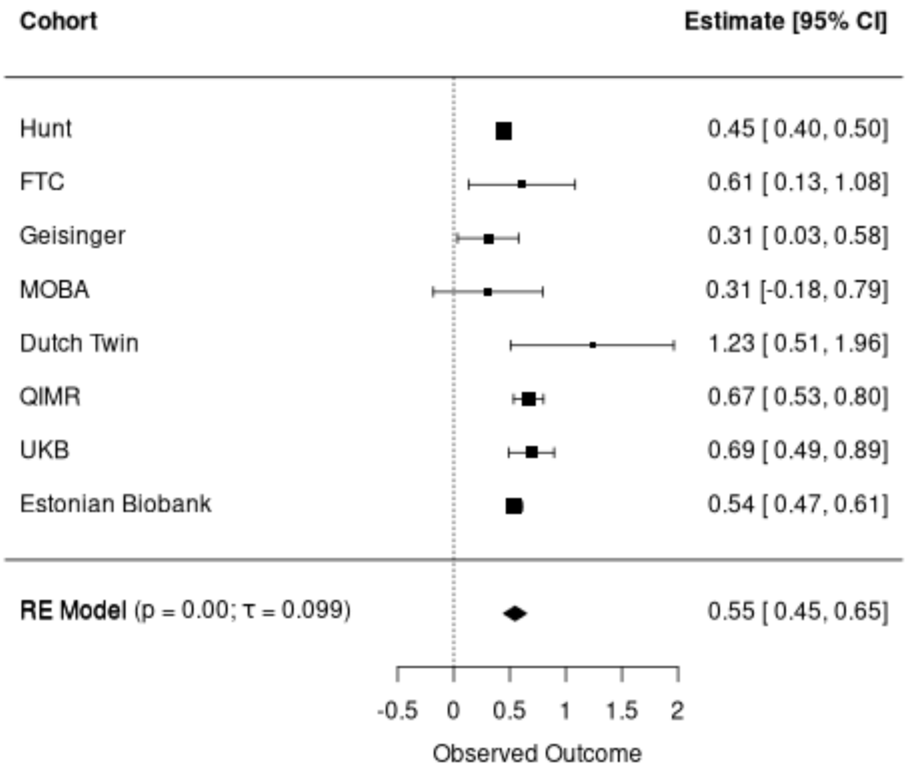


**Supplementary Figure 4. Random effects meta-analysis of cohort-level DGE-population correlation estimates for educational attainment (EA)**. Estimates are from applying the *snipar correlate.py* script to cohort level summary statistics. Only includes estimates with standard error < 0.5. Error bars show 95% confidence intervals. Abbreviations: FTC, Finnish Twin Cohort; MOBA, Norwegian Mother, Father and Child Cohort; QIMR, QIMR Berghofer Medical Research Institute; Dutch Twin, Netherlands Twin Register; Hunt, Trøndelag Health Study; UKB, UK Biobank.

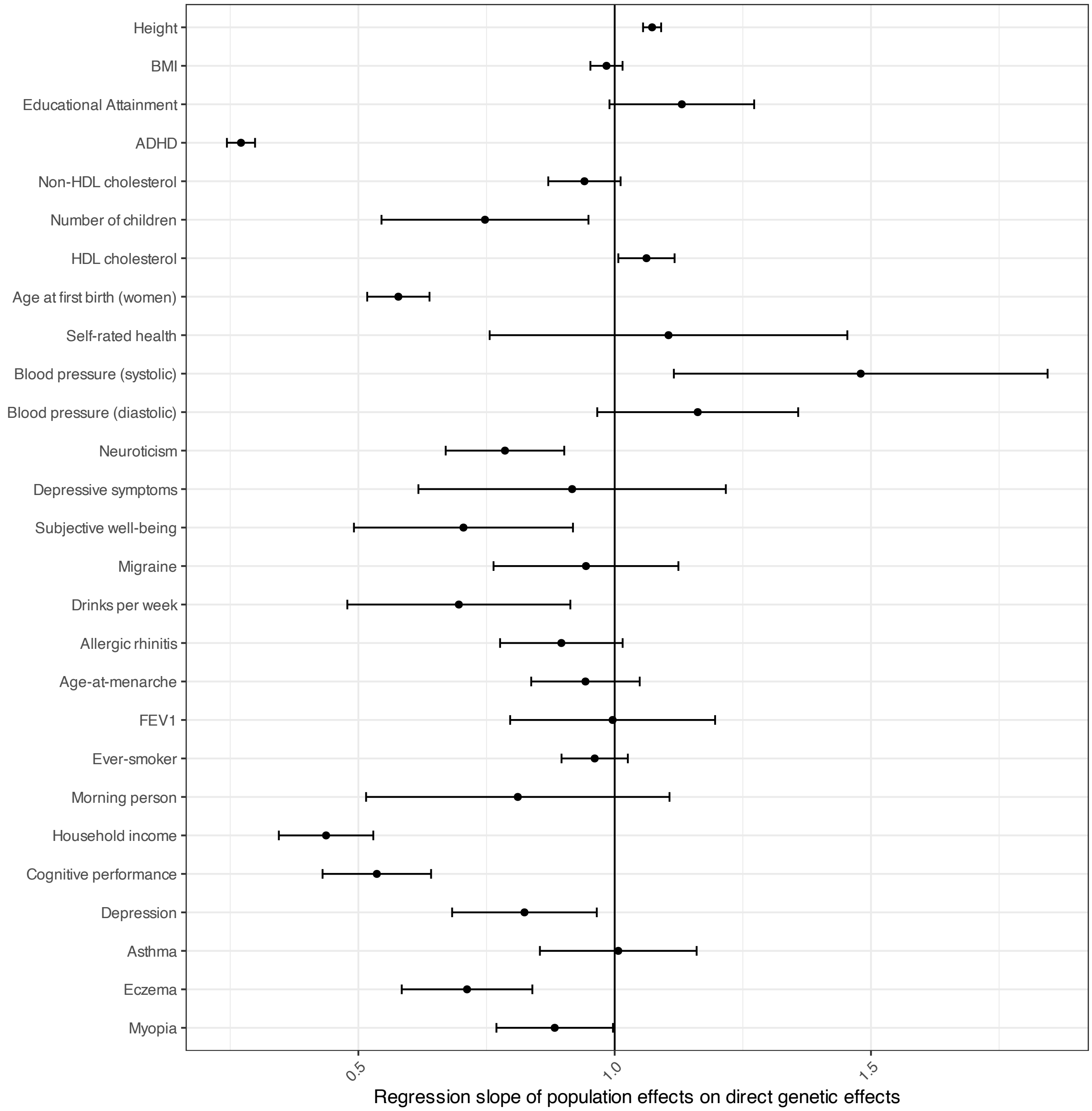


**Supplementary Figure 5. Slope from genome-wide regression of population effects on direct genetic effects**. Using a method implemented in *snipar*, we performed a regression of population effects onto direct genetic effects (DGEs) that accounts for the sampling errors in the estimates (Methods). Here we give the slope which is a measure of the degree to which population effects are systematically inflated/deflated relative to DGEs, with values above 1 indicating population effects are systematically larger than DGEs and values below 1 indicating that population effects are systematically smaller than DGEs. Horizontal bars give 95% confidence intervals. Only includes phenotypes with median direct effective N > 5000 and population-direct regression SE < 0.25. Abbreviations: HDL, high density lipoprotein cholesterol; FEV1, forced expiratory volume in 1 second adjusted for height; BMI, body mass index; EA, educational attainment; ever-smoker, whether an individual has ever smoked.

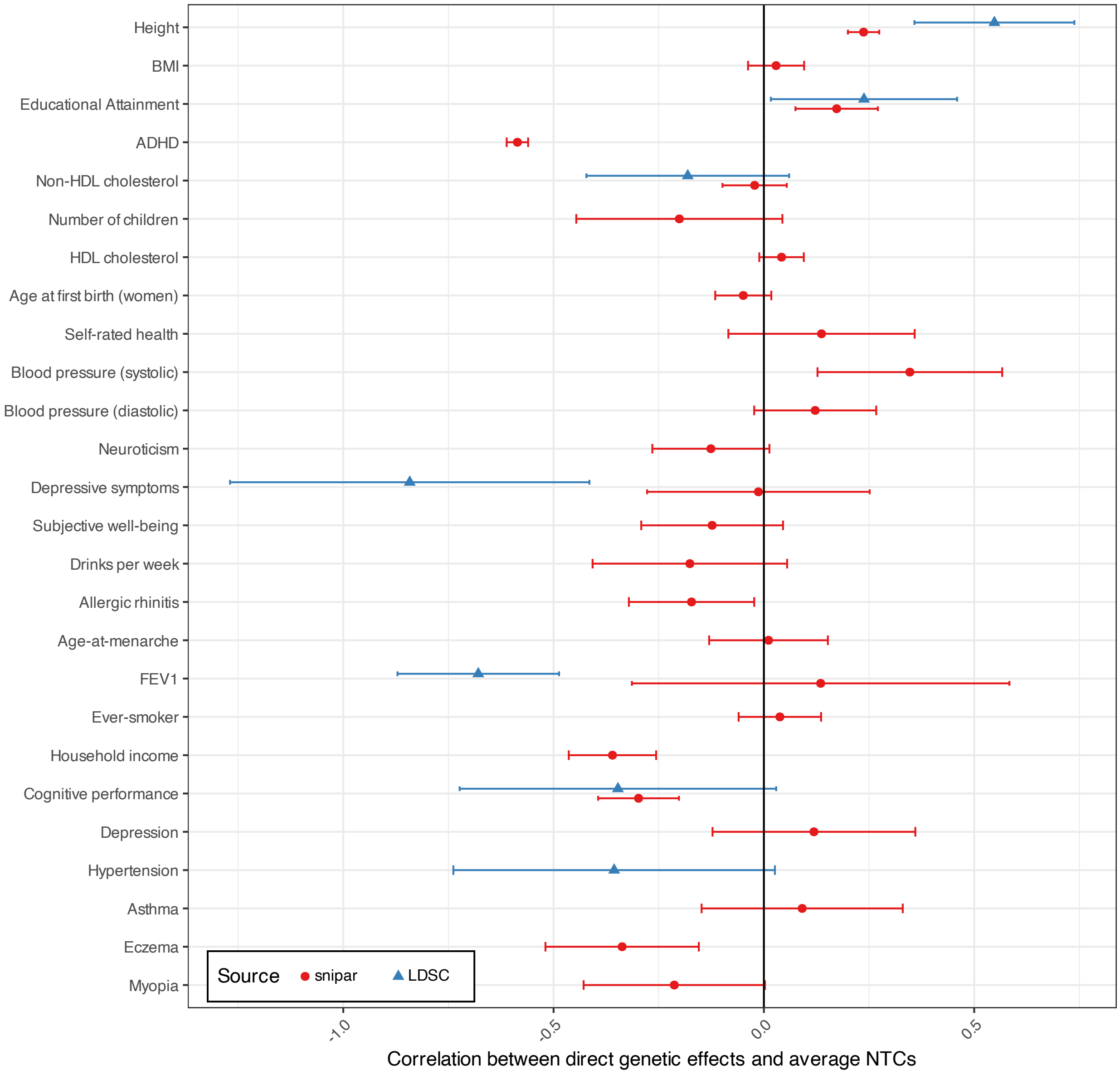


**Supplementary Figure 6. Genome-wide correlations between direct genetic effects (DGEs) and average non-transmitted coefficients (NTCs).** Correlations were estimated using *snipar* and linkage disequilibrium score regression (LDSC). Horizontal bars give 95% confidence intervals. Only includes phenotypes with median DGE effective N > 5000 and SE < 0.25. Abbreviations: HDL, high density lipoprotein cholesterol; FEV1, forced expiratory volume in 1 second adjusted for height; BMI, body mass index; EA, educational attainment; ever-smoker, whether an individual has ever smoked.

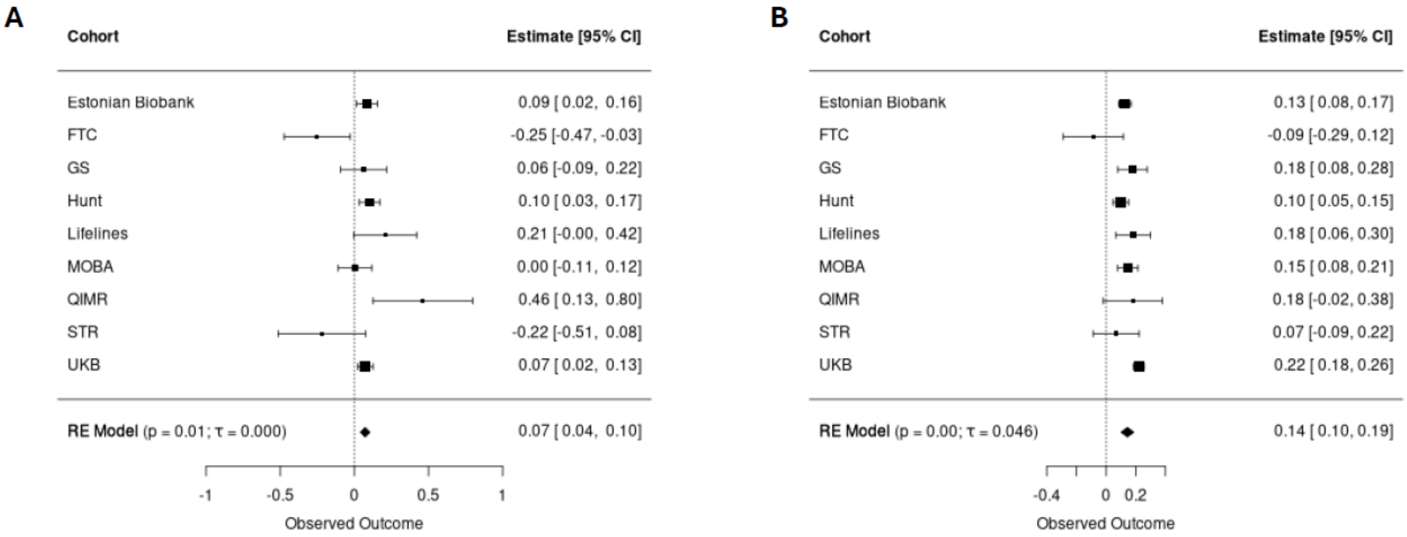


**Supplementary Figure 7. Random effects meta-analysis of cohort-level direct genetic effect (Panel A) and population effect (Panel B) SNP heritability estimates for educational attainment (EA)**. Only includes estimates with direct genetic effect heritability standard error < 0.25. Error bars show 95% confidence intervals. Abbreviations: FTC, Finnish Twin Cohort; GS, Generation Scotland; MOBA, Norwegian Mother, Father and Child Cohort; QIMR, QIMR Berghofer Medical Research Institute; STR, Swedish Twin Register; Hunt, Trøndelag Health Study; UKB, UK Biobank.

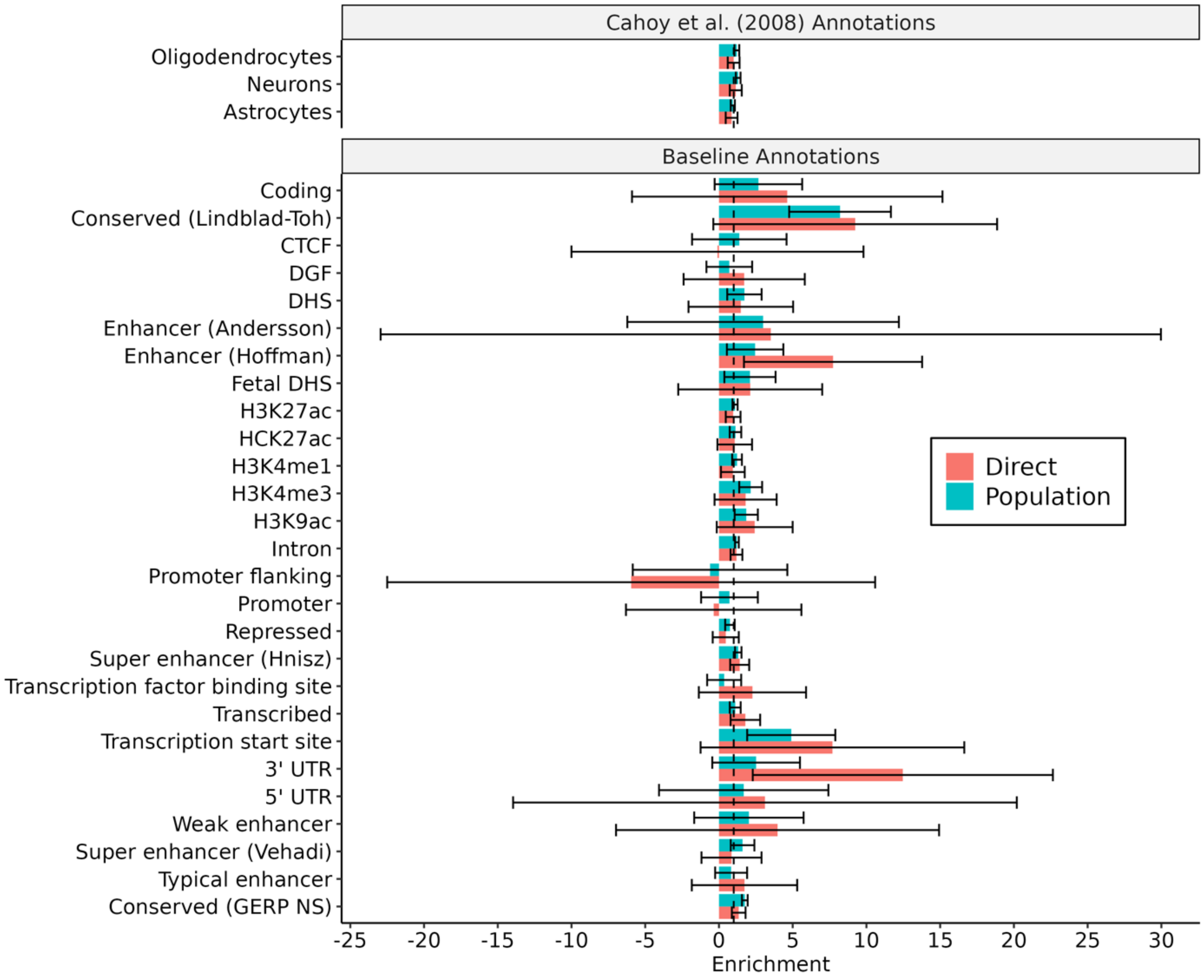


**Supplementary Figure 8. Stratified LD score regression (SLDSC) enrichment for educational attainment**. The figures display the results of running SLDSC on baseline annotations from Gazel et al. (2017) and gene set annotations from Cahoy et al. (2008). Estimates of baseline annotations are from running SLDSC on all the baseline annotations jointly. Estimates of Cahoy et al. (2008) annotations are from running SLDSC on the baseline annotations jointly with one Cahoy et al. annotation added at a time. Analyses were performed using both direct genetic effect summary statistics (red) and population effect summary statistics (teal) from our European ancestry meta-analysis. Error bars are 95% confidence intervals calculated from block jackknife standard errors.

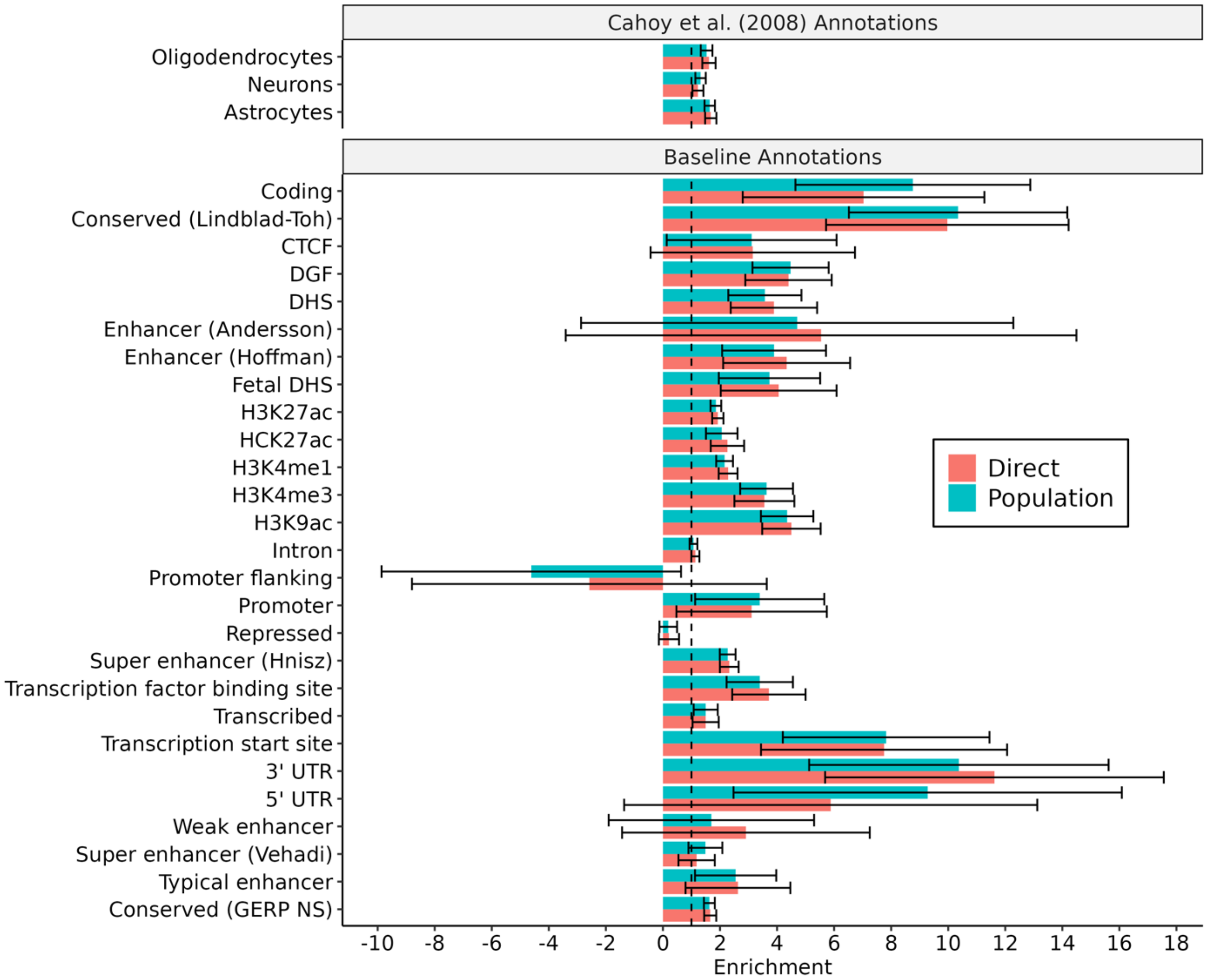


**Supplementary Figure 9. Stratified LD score regression (SLDSC) enrichment for height**. The figures display the results of running SLDSC on baseline annotations from Gazel et al. (2017) and gene set annotations from Cahoy et al. (2008). Estimates of baseline annotations are from running SLDSC on all the baseline annotations jointly. Estimates of Cahoy et al. (2008) annotations are from running SLDSC on the baseline annotations jointly with one Cahoy et al. annotation added at a time. Analyses were performed using both direct genetic effect summary statistics (red) and population effect summary statistics (teal) from our European ancestry meta-analysis. Error bars are 95% confidence intervals calculated from block jackknife standard errors.

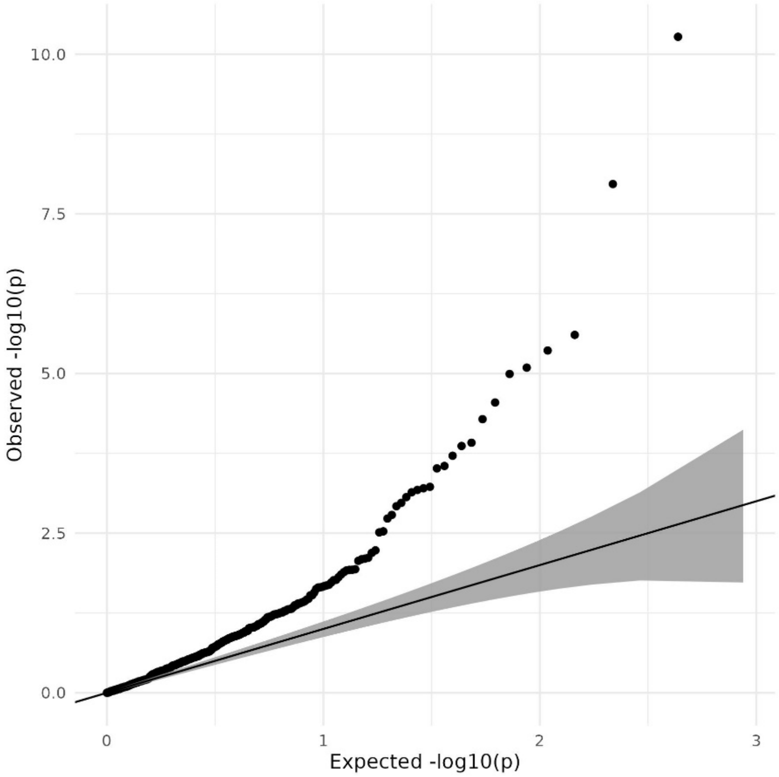


**Supplementary Figure 10. QQplot of -$log_{10}$(p)-values for a difference between genetic correlations calculated using direct genetic effects and population effects.** Shaded area gives 95% confidence interval under the null hypothesis. Numerical values are given in Supplementary Table 7. See Methods for details.

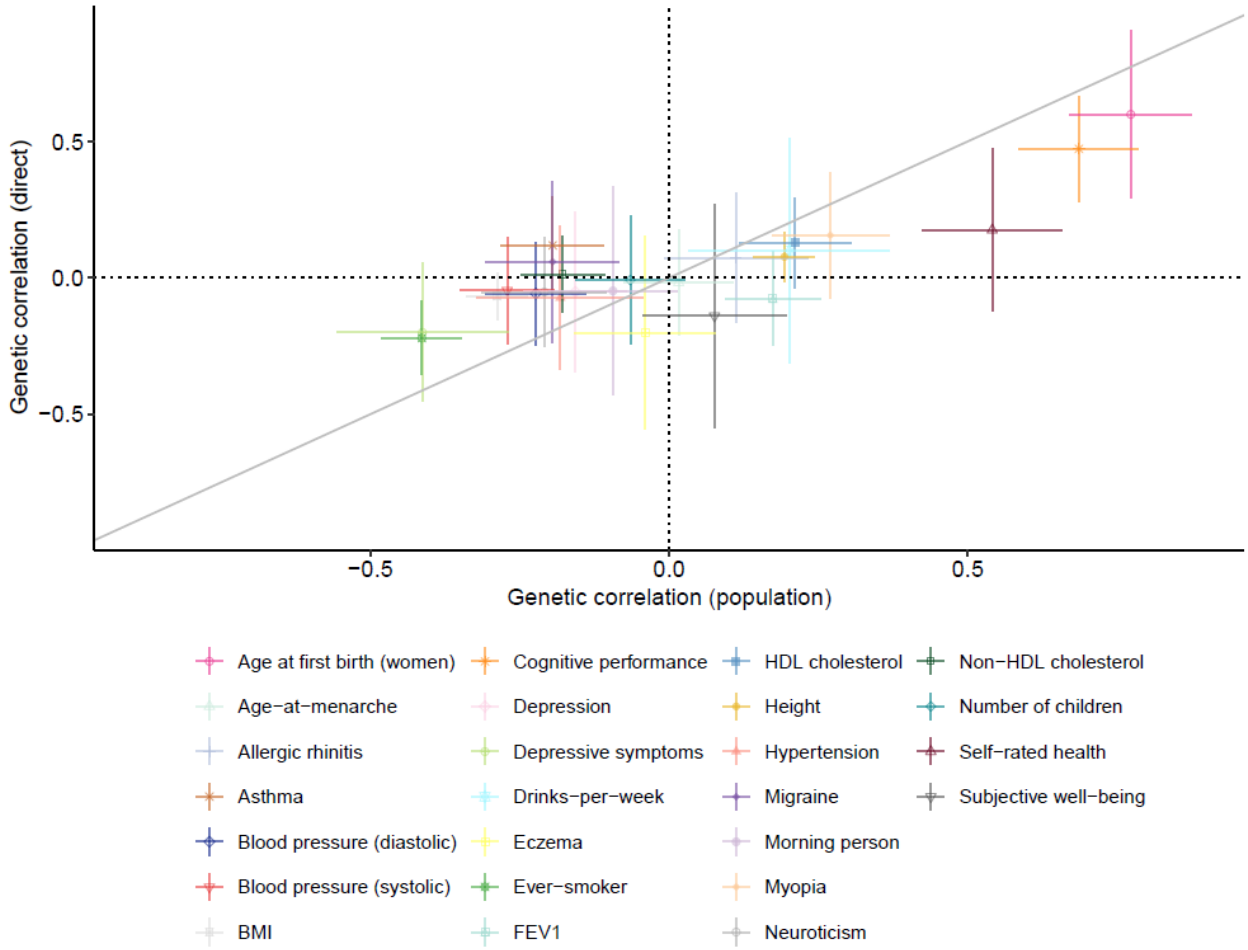


**Supplementary Figure 11. Genetic correlations between educational attainment (EA) and other traits calculated using direct genetic effects (y-axis) and population effects (x-axis).** Only includes phenotypes with median effective N for direct genetic effects greater than 5000 and standard error for genetic correlation from direct genetic effects below 0.25. Error bars give 95% confidence intervals. Abbreviations: ADHD, attention-deficit/hyperactivity disorder; BMI, body mass index; ever-smoker, whether an individual has ever smoked, FEV1, forced expiratory volume in 1 second adjusted for height; HDL, high density lipoprotein.

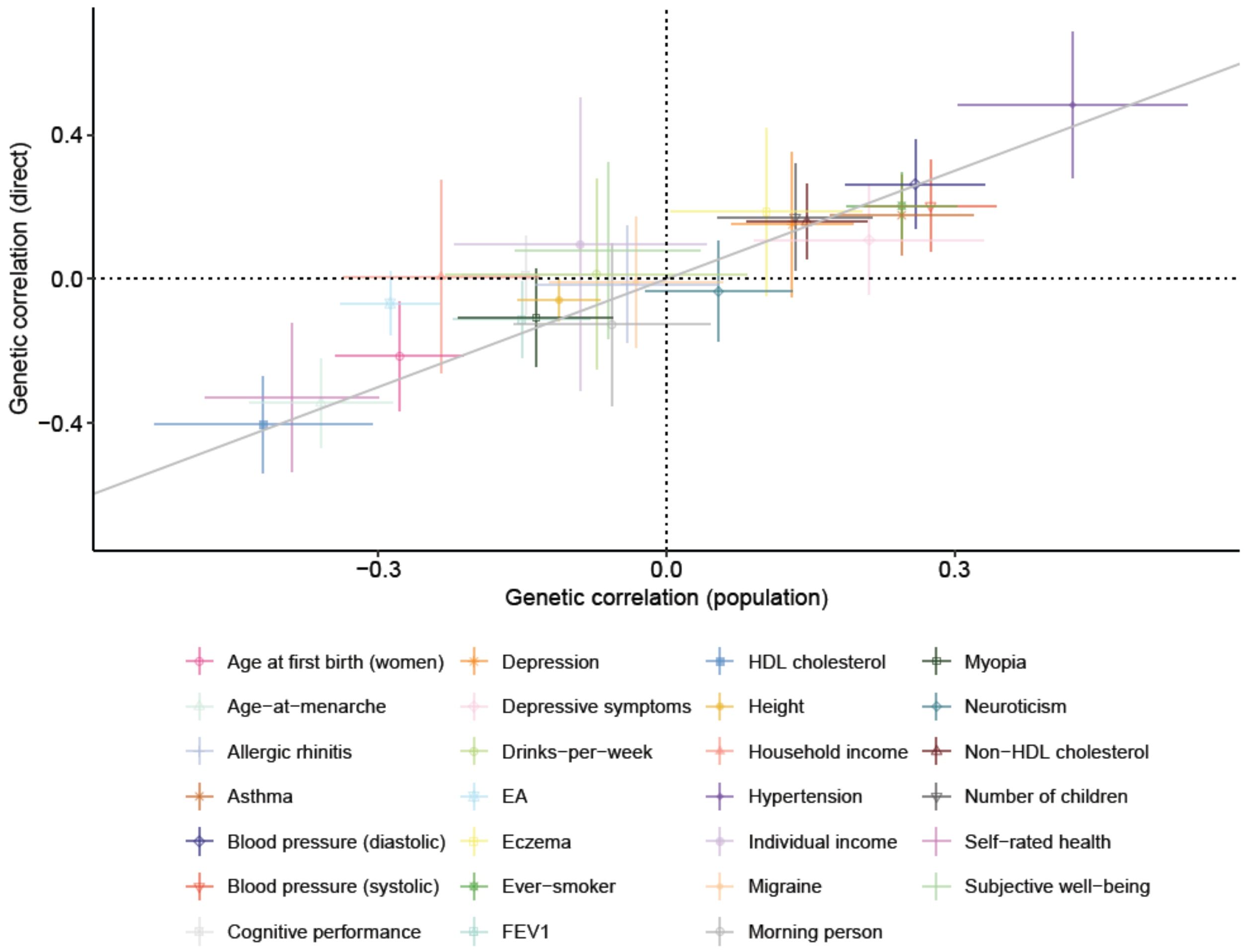


**Supplementary Figure 12. Genetic correlations between BMI and other traits calculated using direct genetic effects (y-axis) and population effects (x-axis).** Only includes phenotypes with median effective N for direct genetic effects greater than 5000 and standard error for genetic correlation from direct genetic effects below 0.25. Error bars give 95% confidence intervals. Abbreviations: ADHD, attention-deficit/hyperactivity disorder; EA, educational attainment; ever-smoker, whether an individual has ever smoked, FEV1, forced expiratory volume in 1 second adjusted for height; HDL, high density lipoprotein.

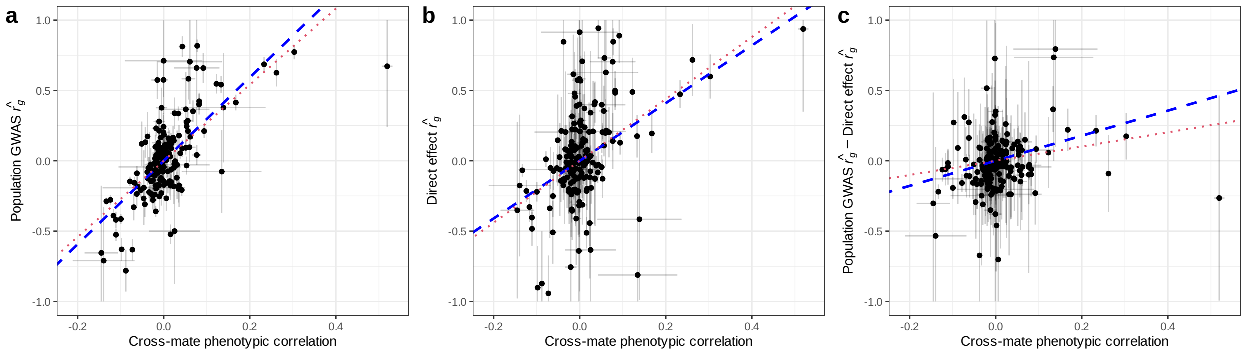


**Supplementary Figure 13. Associations between cross-mate phenotypic correlations and genetic correlation estimates**. Genetic correlation estimates derived from (a) population effects and (b) direct genetic effects (DGEs), as well as (c) their difference. Crosshairs depict standard errors (truncated to [-1,+1] for readability) and the fitted lines reflect ordinary least squares (red, doted) and measurement-error-aware Bayesian (blue, dashed) regression models (Methods).

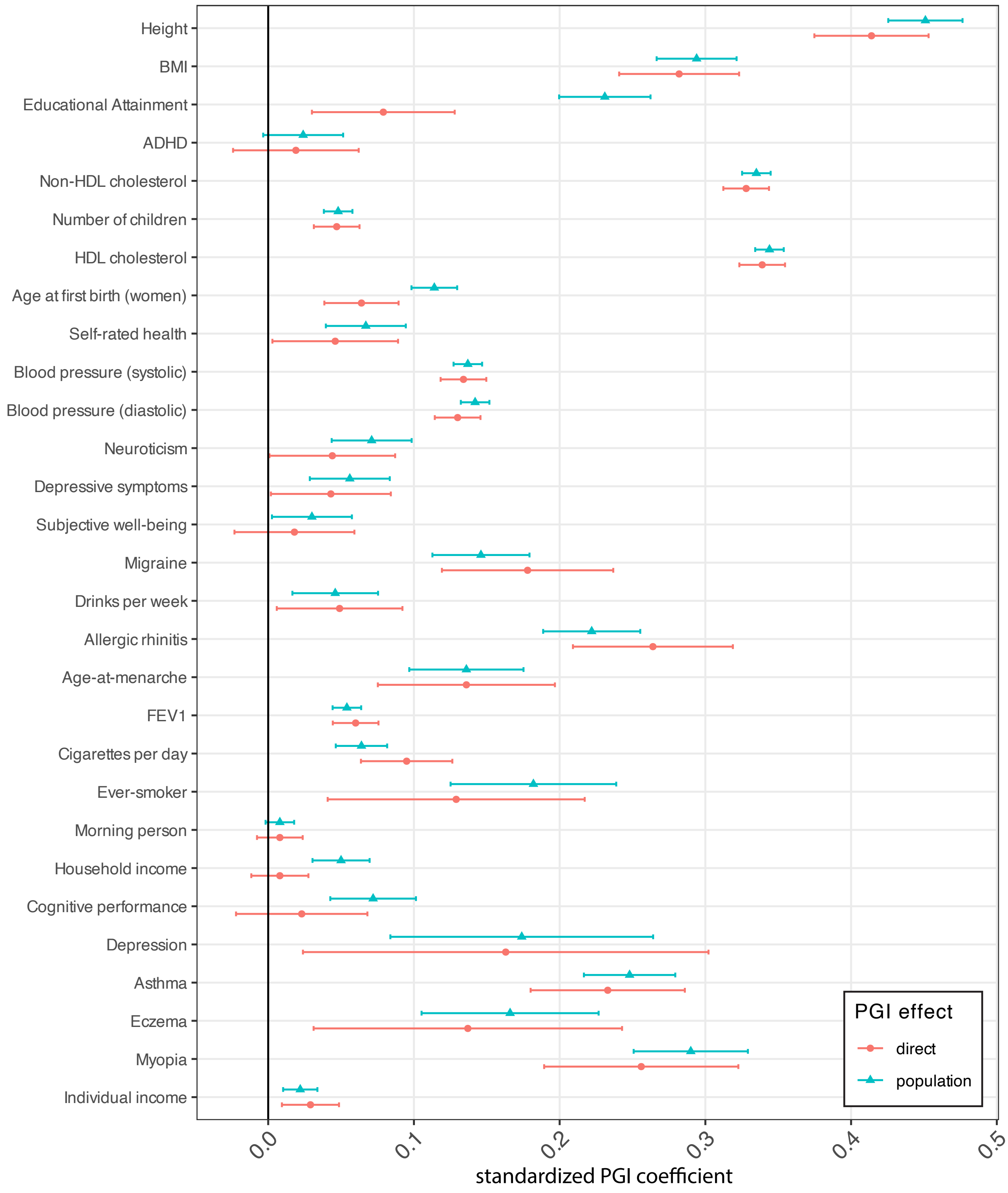


**Supplementary Figure 14. Out-of-sample polygenic prediction analyses using population genetic effect summary statistics.** We give standardized effect estimates (for phenotype and PGI normalized to 1), corresponding to partial correlation coefficients. Error bars give 95% confidence intervals. Abbreviations: EA, educational attainment (years); BMI, body mass index; HDL, high density lipoprotein; FEV1, forced expiratory volume in 1 second; Ever-smoker, whether an individual has ever smoked; Non-HDL, total cholesterol minus HDL cholesterol.

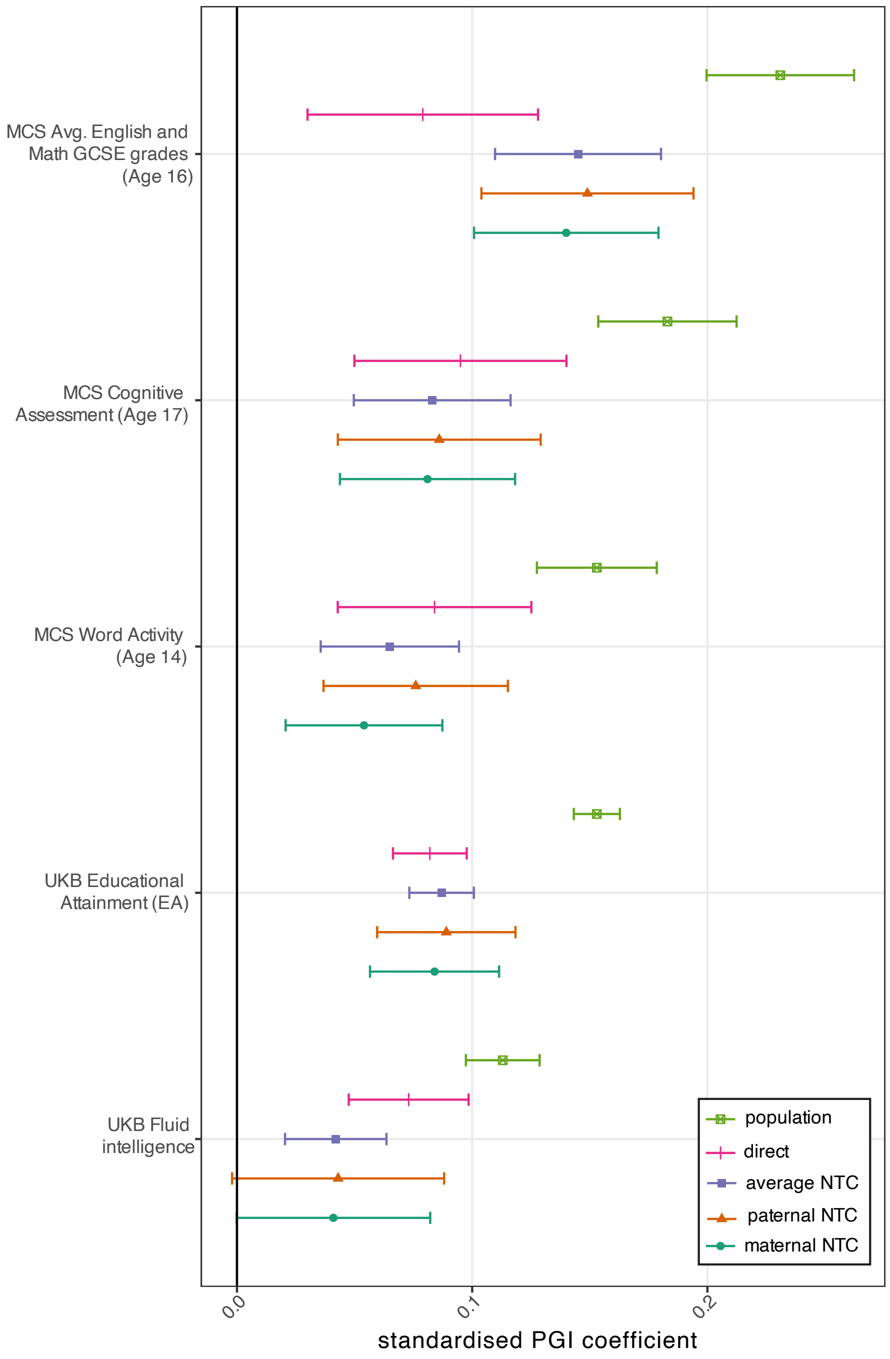


**Supplementary Figure 15. Out-of-sample polygenic prediction analysis using population effect PGIs for educational attainment (EA) to predict education and cognitive performance related outcomes.** Error bars give 95% confidence intervals. Outcome phenotypes: Avg. Eng. & Math GCSE Score, mean of English and Math GCSE grade Z-scores; educational attainment outcome as defined in Okbay et al. (2022); MCS S6 Word Activity, Word Activity score from MCS Sweep 6; MCS S7 Cognitive Assessment, cognitive assessment outcome from MCS Sweep 7; UKB Fluid Intelligence, fluid intelligence score from UK Biobank. Full descriptions of outcome phenotypes can be found in Supplementary Table 8 and Supplementary Note Section 4. Numerical values are contained in Supplementary Table 9.
