## Supplementary File 1 Analysis Plan for "Family-GWAS reveals effects of environment and mating on genetic associations"

### Analysis plan for 'Meta-analysis of Mendelian Imputation based Genome-Wide Association Studies'

#### Overview

We are conducting a meta-analysis of family-based genome wide association studies aided by Mendelian Imputation of missing parental genotypes. The underlying model for the effect of a SNP on a phenotype is:

$$E\left[Y_{ij} \middle| g_{ij}, g_{p(i)}, g_{m(i)}\right] = \delta g_{ij} + \beta_p g_{p(i)} + \beta_m g_{m(i)},$$

where $Y_{ij}$ is the phenotype of sibling $j$ in family $i$, $g_{ij}$ is the genotype of that individual at the SNP, $g_{p(i)}$ is the genotype of the father, and $g_{m(i)}$ is the genotype of the mother; $\delta$ is the direct effect of the SNP, $\beta_p$ is the 'paternal effect' of the SNP, and $\beta_m$ is the 'maternal effect' of the SNP.

When parental genotypes are unobserved, they are imputed from the observed genotypes in the family according to the inferred inheritance pattern (Mendelian Imputation), where identity-by-descent sharing information is leveraged as appropriate: see https://www.biorxiv.org/content/10.1101/2020.07.02.185199v1.abstract.

For cohorts where no parental genotypes are observed, the imputed maternal and paternal genotypes are the same, so we use the imputed sum of maternal and paternal genotypes and estimate $\delta$ and $(\beta_p + \beta_m)/2$. Despite the fact that some cohorts cannot directly estimate $\beta_p$ and $\beta_m$ separately, we will use a form of multivariate meta-analysis to produce meta-analysis estimate of $\delta$, $\beta_p$, $\beta_m$ for each SNP. From these meta-analysis estimates of the underling parameter vector, we will be able to produce meta-analysis estimate of linear transformations of this parameter vector, including average parental effects, $(\beta_p + \beta_m)/2$, and 'population effects', $\delta + (\beta_p + \beta_m)/2$, as estimated by standard GWAS of unrelated individuals.

#### Phenotypes

| Phenotype | Age restriction | Coding/notes |
|---|---|---|
| ADHD | None | Clinical diagnosis or self-report; binary coding |
| Age at first birth | NA | NA |
| age at menarche | NA | NA |
| asthma | None | ICD10, self-report (of diagnosis or symptoms) or clinical diagnosis; binary coding |
| Eczema (atopic dermatitis) | None | ICD10, self-report (of diagnosis or symptoms) or clinical diagnosis; binary coding |
| BMI | 18+ | None |
| HDL Cholesterol | 18+ | None |
| Non-HDL cholesterol | 18+ | Total cholesterol-HDL cholesterol |
| Blood pressure | 18+ | Both diastolic and systolic |
| ever cannabis | 18+ | Self-report; Binary coding |
| cigarettes per day | 18+ | both former and current smokers; those with no data set to missing |
| Drinks-per-week | 18+ | Self-report; quantitative, midpoint of range used if response is binned |
| depression | 18+ | self-report (of diagnosis or symptoms) or clinical diagnosis; binary coding; exclusions for individuals identified as suffering from other mental illnesses |
| Depressive symptoms | 18+ | Use as quantitative scale |
| educational attainment | 30+ | Map to ISCED-1995 categories and convert to US years of schooling equivalent; contact for help with this |
| Cognitive ability/IQ | None | Use most complete measure of general cognitive ability |
| ever smoker | 18+ | Binary coding |
| extraversion | 18+ | scores based on answers to personality trait tests; quantitative |
| FEV1 | 18+ | Forced expiratory volume in 1 second (lung health measure) |
| hayfever | None | ICD10, self-report (of diagnosis or symptoms) or clinical diagnosis; binary coding |
| height | 18+ | None |
| Household income | 18+ | self-report: if binned variable analyze by treating the midpoints of categories of income as a continuous variable; log scale |
| Hourly income | 18+ | Imputed from occupation using survey data; transformed to log scale; quantitative |
| migraine | 18+ | self-report or clinical diagnosis or diagnosis from questionnaire responses; binary coding |
| morning person | 18+ | Self-report; Binary coding |
| nearsightedness | None | questionnaire responses or self-report (of diagnosis or symptoms); binary coding |

| | | |
|---|---|---|
| number ever born | 45+ for men; 55+ for women | None |
| neuroticism | 18+ | scores based on answers to personality trait tests; quantitative |
| self-rated health | 18+ | Use as quantitative scale from 1 (worst self-rated health) to N (best self-rated health), where N is the number of possible ranked answers to survey question |
| subjective well-being | 18+ | Use as quantitative scale from 1 (worst subjective well-being) to N (best subjective well-being), where N is the number of possible ranked answers to survey question; combine scores from questions for 'positive affect' and 'life satisfaction' if appropriate |

#### Cohort Level Analyses

1. Filter variants for MAF>1% and INFO>0.99, or equivalent imputation information metric, and convert to .bed format. (If phased haplotypes are available, do the same and put in phased .bgen format.)
2. Run KING with –related –degree 1 options to infer first degree relatives
3. Run KING with –ibdsegs –degree 1 options to infer IBD segments between first-degree relatives
4. Download and install SNIPar: https://github.com/AlexTISYoung/SNIPar
5. Run tests (python setup.py pytest) to ensure correct compilation and installation
6. Impute each chromosome separately using the impute_runnner.py script: see the tutorial for examples https://github.com/AlexTISYoung/SNIPar/blob/master/docs/tutorial.rst. If memory is an issue, use the –chunks argument to read the SNPs in chunks to reduce maximum memory usage. The script takes the IBD segments and relations inferred by KING along with a plain text file giving the age and sex of each individual (columns FID, IID, age, sex), with sex coded as 'M' for male or 'F' for female. These are used to construct a pedigree and to perform the imputation based on the pedigree and the IBD segments. Alternatively, a user-input pedigree can be provided with columns FID, IID, FATHER_ID, MOTHER_ID
7. Run fGWAS.py script for each chromosome for each phenotype. Phenotypes should be adjusted for age at measurement (and year-of-birth if different), sex, age^2, age^3, age*sex, age^2*sex, age^3*sex and the top 20 principal components. This can be done before the phenotype is input to the fGWAS.py, or the covariates can be provided to the fGWAS.py script. The script takes the observed genotypes, the imputed genotypes, and the phenotype file as minimal inputs. This will output HDF5 and gzipped text summary statistics.
8. Send text and HDF5 summary statistics files to. Please also send any other imputation/genotyping quality metrics on the SNPs, such as imputation

INFO, call-rate, and Hardy-Weinberg Equilibrium P-value. Please also send the allsegs output from the KING IBD inference that contains the intervals where IBD is assessed.

#### Meta-analysis and downstream analyses

1. We will estimate the variance due to direct and population effects within each cohort, as well as their correlation. We will check the correlation between the population effects and publicly available summary statistics as a form of quality control.
2. We will use METAL and multivariate meta-analysis to obtain meta-analysis estimates of direct, paternal, and maternal effects at each SNP for each phenotype.
3. We will construct polygenic indexes based on direct effects only for each phenotype and test their predictive ability in the population and within-families in left-out prediction cohorts.
4. We will estimate genetic correlations between phenotypes using direct-effect summary statistics.
5. For each phenotype, we will estimate genetic correlations between direct and average parental effects and between direct and population effects.
