## Supplementary Note for "Family-GWAS reveals effects of environment and mating on genetic associations"

### 1. Quality Control for family-based GWAS

Summary statistics provided by each cohort are passed through a quality control pipeline. The pipeline serves to filter out low-quality SNPs and to carry out data operations that prepare the summary statistics for meta-analysis; the pipeline also generates diagnostic plots.

First, we give more detail on our analysis of the relationship between imputation quality and sibling genotype correlation (Supplementary Figure 1). We analysed variants with INFO scores (a measure of imputation quality that estimates the fraction of genotype variation the imputation has recovered) ranging between 0.3 and 1 and minor allele frequencies (MAF) greater than 1% in a sample of 19,290 sibling pairs from the white British subsample of the UK Biobank. For imputed genotype dosages, the mean correlation between siblings' genotypes across 1000 SNPs with INFO scores between 0.30 and 0.31 and minor allele frequency (MAF) at least 1% was 0.437 (S.E.=0.001), much lower than the expected correlation of 0.5 based on Mendelian laws; for imputed hard calls (most likely genotype, used in the Howe et al. sib-GWAS[9]), the correlation was even lower at 0.376 (S.E.=0.001). The correlation between siblings' genotypes only approaches the expected correlation of 0.5 as the INFO score gets very close to 1, with detectable deviations from the expectation for hard-call genotypes for SNPs with INFO scores as high as 0.96-0.97: mean correlation 0.4983 (S.E.=$4.7 \times 10^{-4}$, P=$1.6 \times 10^{-4}$ for correlation below 0.5 from a one-sided Z-test). These results imply all but the highest quality imputed genotypes are appropriate for family-based analyses that assume genotypes follow Mendelian Laws. We therefore set a stringent imputation quality threshold of 0.99 for our quality control procedure. One consequence of the stringent QC requirement of FGWAS is that it results in lots of variants being filtered out in many cohorts, resulting in variable coverage of genome-wide variants from different cohorts.

#### 1.1. Data Operations

##### 1.1.1. Rescaling

Effect estimates in each cohort were first divided by the phenotypic standard deviation in that cohort to put them on a consistent scale across cohorts. For binary phenotypes, we divided effect estimates by the phenotypic variance to transform them to the logistic scale[56].

##### 1.1.2. Variant filters

We apply the following variant filters using EasyQC[57]:

- Filter out variants with missing values for any of: variant ID, chromosome, reference and/or alternative alleles, allele frequency, effect estimates and their standard errors
- Filter out variants with allele frequencies outside the range
- Filter out variants with reference or alternative alleles not in the set {'A', 'C', 'G', 'T'}
- Keep only variants on chromosomes 1 to 22
- Filter out monomorphic variants
- Filter out variants with a Hardy-Weinberg Equilibrium exact test P-value less than
- Filter out variants which have a call rate of less than 0.99

- Filter out variants which have an imputation accuracy (INFO score or imputation $R^2$) less than 0.99.
- Filter out variants with minor allele frequency (MAF) < 0.01.
- Filter out variants with duplicated positions, i.e. variants with positions shared by other variants in the summary statistics
- Bin variants by allele frequency (at increments of 0.01) and calculate the mean effective sample size based on DGEs and the standard deviation of DGE effective N for SNPs in each allele frequency bin. Filter out SNPs with effective N outside the mean ± 5 S.D. Repeat for average NTC effective N.

The above filters may still miss SNPs with high genotyping error rates or in regions where IBD information is missing or unreliable. Both high genotyping error rate and missing/unreliable IBD information will lead to an atypical relationship between proband and observed/imputed parental genotypes at that SNP — e.g. a lower correlation than is possible based on Mendelian Laws. The sampling correlations between estimates of different elements of the parameter vector are themselves functions of the correlations between the offspring and observed/imputed parental genotypes in the regression design matrix. Therefore, variants that have outlying sampling correlations are indicative of issues that may have affected the results for that SNP, including low genotype quality (e.g. leading to high Mendelian error rate) and/or errors in IBD inference/imputation in the *snipar* pipeline[3,58].

This suggests a novel form of quality control for FGWAS: to filter out SNPs with outlying sampling correlations between effects (Supplementary Figure 2). There is a cluster of SNPs in the bottom right corner of Panel A of Supplementary Figure 2 with outlying sampling correlations. To remove such variants, we filter out variants whose correlations are more than 6 standard deviations away from the mean sampling correlation across variants. We do this for every effect-pair (i.e., DGE-population, DGE-average NTC, maternal NTC-paternal NTC, etc.)

##### 1.1.3. Aligning Alleles

We align the alleles, allele frequencies, and estimated effects to the public release of the Haplotype Reference Consortium (HRC)[59] reference panel. Only SNPs present in the HRC reference panel are kept. Additionally, SNPs with alleles that do not match the HRC reference data are removed – for example if the alleles in HRC show AG but the alleles in cohort data show AC.

#### 1.2. Diagnostic Plots

##### 1.2.1. Comparison of allele frequencies with reference

After filtering variants and aligning alleles, we check the sample allele frequencies against the HRC allele frequencies. We show an example from the UK Biobank summary statistics in Supplementary Note Figure 1.

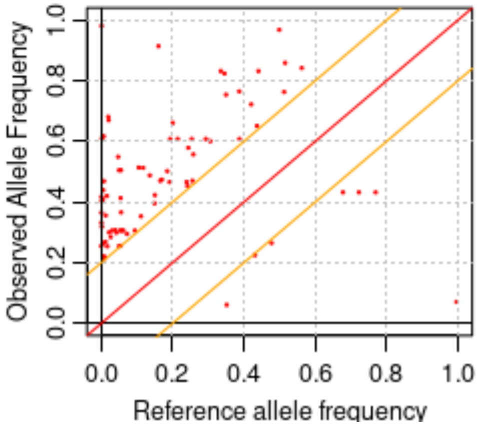


***Supplementary Note Figure 1. Allele frequency plot.*** *The x-axis plots the reference allele frequency from the HRC; the y-axis plots the allele frequencies from UK Biobank summary statistics. The red diagonal line in the center is the 45-degree line, indicating equality of sample and reference allele frequency. The yellow diagonal lines show the boundaries within which the sample allele frequency is within +/- 0.2 of the reference allele frequency. Points outside of this range are plotted as red points.*

#### 1.2.2. QQ-plots

We generate QQ-plots for each effect estimated in each cohort. We show an example from the Educational Attainment (EA) phenotype from the UK Biobank in Supplementary Note Figure 2:

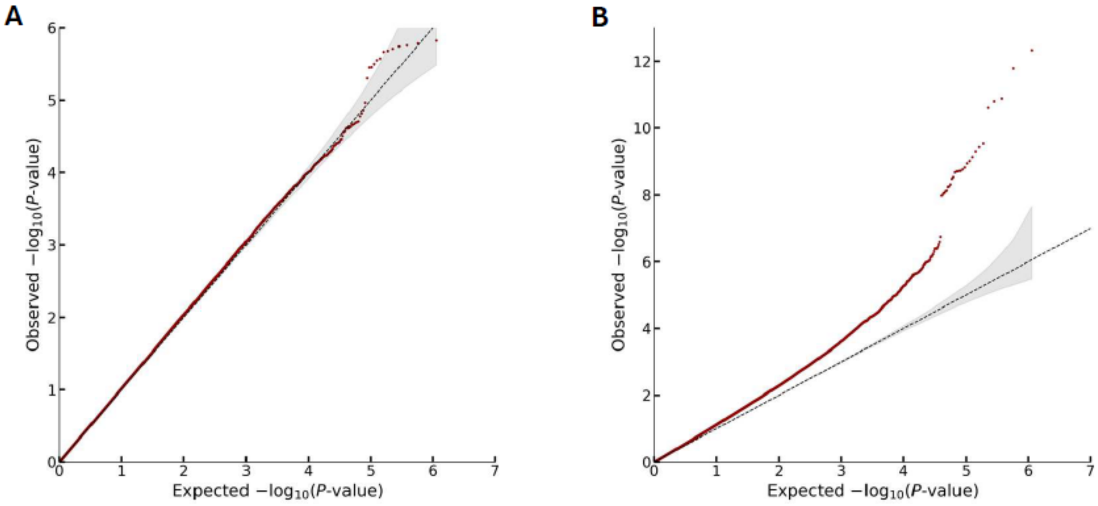


***Supplementary Note Figure 2. Quantile-Quantile (QQ) plots for educational attainment summary statistics from the UK Biobank.*** *A) QQ-plot for direct genetic effects. B) QQ-plot for population effects.*

We visually inspected the QQ-plots for unusual patterns or excessive inflation. We did this for each effect estimated in each cohort.

#### 1.2.3. SE vs Allele Frequency Plots

Based on theory, we expect that the standard errors should be approximately proportional to $1/\sqrt{f\ (1-f)}$, where f is the minor allele frequency, implying that a plot for a set of summary statistics without any issues would look like Supplementary Note Figure 3. The points should be tightly clustered and there shouldn't be many outliers from the expected relationship. We checked these plots for each estimated effect in each cohort.

#### 1.2.4. Effective sample size vs allele frequency plots

We produced plots of effective samples size against allele frequency using summary statistics on DGEs, population effects, and average NTCs. We expect these plots to indicate no clear relationship between effective sample size and allele frequency, as illustrated in Figure 5 below.

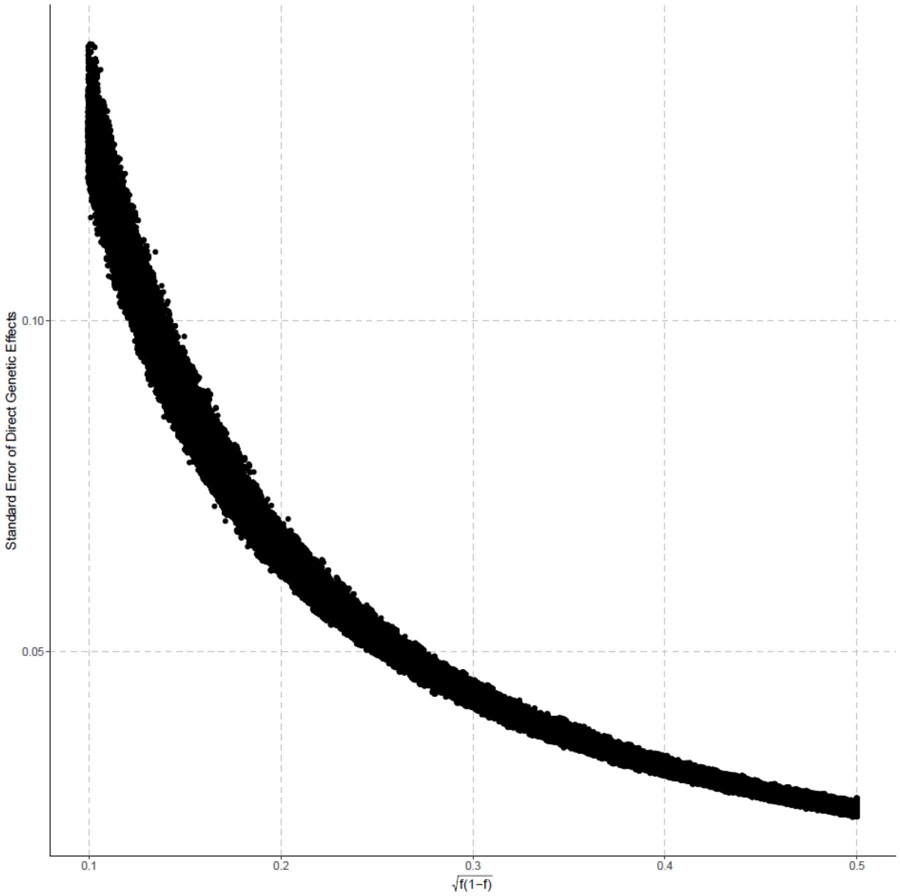


***Supplementary Note Figure 3. Standard error-MAF plot.*** *The x-axis plots* $\sqrt{f\,(1-f)}$*, where* $f$ *is the minor allele frequency; the y-axis plots the standard error of DGEs from the summary statistics on BMI from Framingham Heart Study.*

### 1.3. Genetic correlation with reference GWAS

We calculate the genetic correlation between the population effect estimates and estimates from a reference GWAS using LD-score regression (LDSC). Supplementary Table 10 shows the reference GWAS studies, and Supplementary Table 3 shows the cohort level results. We expect this to be close to 1, and we used this to discover and fix any issues such as reverse-coding of phenotypes at the cohort level. We decided not to include summary statistics on EA from Botnia due to a low genetic correlation with the reference GWAS.

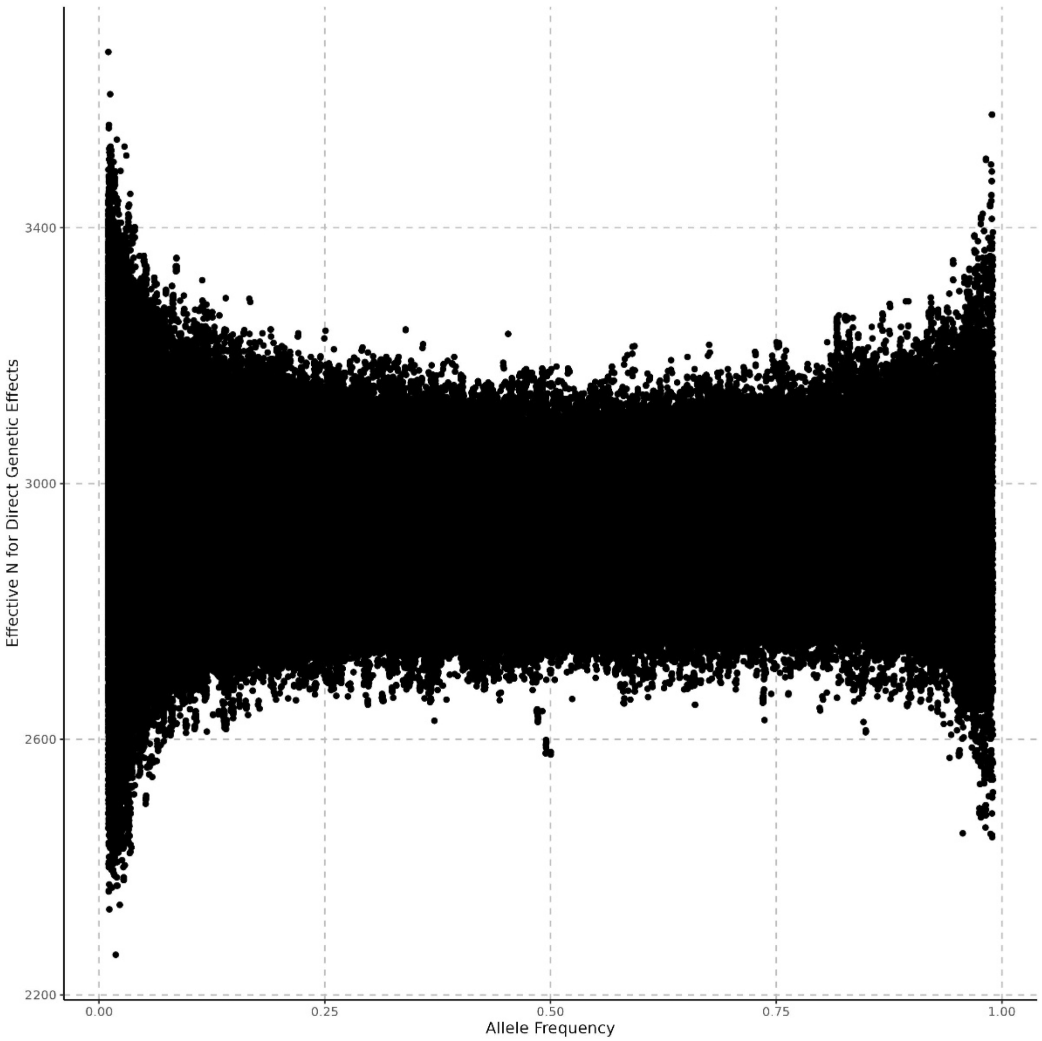


***Supplementary Note Figure 5. Effective sample size against allele frequency plots.*** *The x-axis plots allele frequency for each SNP, the y-axis plots the effective sample size for direct genetic effects for each SNP from the summary statistics on BMI from Framingham Heart Study.*

### 2. Estimating variances and covariances of effects

#### 2.1 Estimating variances

To estimate variances, we consider weighted estimators and optimize the weights for minimum sampling variance. Let $v_\delta = \text{Var}_l(\delta_l)$, then we consider an estimator of the form:

$$\hat{v}_\delta = \sum_{l=1}^{L} w_l\left(\hat{\delta}_l^2 - \sigma_{\delta l}^2\right)$$

For this estimator to be unbiased, we require that $\sum_{l=1}^{L} w_l = 1$. Using the constraint as a Lagrange multiplier, we seek $w_l$ that minimizes

$$L(\hat{v}_\delta) = \sum_{l=1}^{L} w_l^2 \text{Var}\left(\hat{\delta}_l^2\right) + \lambda\left(\sum_{l=1}^{L} w_l - 1\right)$$

To make this tractable, we assume that $\delta_l \sim \mathcal{N}(0, v_\delta)$, so that marginally,

$$\hat{\delta}_l \sim \mathcal{N}\left(0, v_\delta + \sigma_{\delta l}^2\right).$$

Although we use this assumption to derive an optimal weighting, this assumption is not required for the estimator to be unbiased. Given this assumption, we can apply the formula for the variance of a scaled, central $\chi^2$ to obtain $\text{Var}\left(\hat{\delta}_l^2\right) = 2\left(v_\delta + \sigma_{\delta l}^2\right)^2$. Therefore,

$$L(\hat{v}_\delta) = 2\sum_{l=1}^{L} w_l^2 \left(v_\delta + \sigma_{\delta l}^2\right)^2 + \lambda\left(\sum_{l=1}^{L} w_l - 1\right)$$

Differentiating with respect to $w_l$ and $\lambda$ and solving for the optimum, we obtain

$$w_l = \frac{\left(v_\delta + \sigma_{\delta_l}^2\right)^{-2}}{\sum_{l=1}^{L} \left(v_\delta + \sigma_{\delta_l}^2\right)^{-2}}.$$

Since the weights depend upon the parameter being estimated, we propose an iteratively reweighted estimator: we initialize $v_\delta = 0$ to obtain $w_l$, then estimate $v_\delta$ with those weights, then re-estimate the weights, and re-estimate $v_\delta$ given the new weights, until the $v_\delta$ estimates converge. We stopped iterations when the change in the estimate was less than $10^{-8}$.

2.2 Estimating covariances

Consider estimating $\text{Cov}_l(\delta_l, \beta_l)$ using the following weighted estimator:

$$\hat{c}_{\delta\beta} = \sum_{l=1}^{L} w_{cl}\left(\hat{\delta}_l \hat{\beta}_l - r_l \sigma_{\delta l} \sigma_{\beta l}\right).$$

Assuming independence between variants, the variance of this estimator is:

$$\text{Var}\left(\hat{c}_{\delta\beta}\right) = \sum_{l=1}^{L} w_{cl}^2 \text{Var}\left(\hat{\delta}_l \hat{\beta}_l\right).$$

Let $\text{Var}_l\,(\beta_l) = v_\beta$. We now show that, assuming that $\beta_l$ is normally distributed,

$$\text{Var}\left(\hat{\delta}_l \hat{\beta}_l\right) = \left(v_\delta + \sigma_{\delta l}^2\right)\left(v_\beta + \sigma_{\beta l}^2\right) + \left(c_{\delta\beta} + r_l \sigma_{\delta l} \sigma_{\beta l}\right)^2$$

We first derive the variance of the product of two correlated but standardized normal random variables, $U_1$ and $U_2$, with correlation $r$. We do this by expressing each of these in terms of two independent normal random variables, $Z_1$ and $Z_2$ :

$$U_1 = Z_1; U_2 = rZ_1 + \sqrt{1-r^2}Z_2$$

It can be confirmed that $U_1$ and $U_2$ have $\mathcal{N}(0,1)$ marginal distributions and that $\text{Corr}\,(U_1, U_2) = r$. We now compute

$$\text{Var}\,(U_1U_2) = r^2\text{Var}\,(Z_1^2) + (1-r^2)\text{Var}\,(Z_1Z_2) + r\sqrt{1-r^2}\text{Cov}\,(Z_1^2, Z_1Z_2);$$
$$\text{Var}\,(U_1U_2) = 2r^2 + (1-r^2) = 1 + r^2;$$

where we have used the fact that $\text{Cov}\,(Z_1^2, Z_1Z_2) = 0$ due to independence of $Z_1$ and $Z_2$ and that $\text{Var}\,(Z_1^2) = 2$ since $Z_1^2 \sim \chi_1^2$. We can then generalize this to variances different from 1. Let $\text{Var}\,(U_1) = v_1$ and $\text{Var}\,(U_2) = v_2$, then

$$\text{Var}\,(U_1U_2) = v_1v_2(1+r^2)$$

Marginally,

$$\begin{bmatrix}\hat{\delta}_l \\ \hat{\beta}_l\end{bmatrix} \sim \mathcal{N}\left(\begin{bmatrix}0 \\ 0\end{bmatrix}, \begin{bmatrix}(v_\delta + \sigma_{\delta l}^2) & (c_{\delta\beta} + r_l\sigma_{\delta l}\sigma_{\beta l}) \\ (c_{\delta\beta} + r_l\sigma_{\delta l}\sigma_{\beta l}) & (v_\beta + \sigma_{\beta l}^2)\end{bmatrix}\right).$$

Therefore, applying the above formula, we obtain

$$\text{Var}\,(\hat{\delta}_l\hat{\beta}_l) = (v_\delta + \sigma_{\delta l}^2)(v_\beta + \sigma_{\beta l}^2) + (c_{\delta\beta} + r_l\sigma_{\delta l}\sigma_{\beta l})^2.$$

Following the same optimization procedure as above for the variance parameters, we obtain optimal weights:

$$w_{cl} = \frac{\left[(v_\delta + \sigma_{\delta l}^2)(v_\beta + \sigma_{\beta l}^2) + (c_{\delta\beta} + r_l\sigma_{\delta l}\sigma_{\beta l})^2\right]^{-1}}{\sum_{l=1}^{L}\left[(v_\delta + \sigma_{\delta l}^2)(v_\beta + \sigma_{\beta l}^2) + (c_{\delta\beta} + r_l\sigma_{\delta l}\sigma_{\beta l})^2\right]^{-1}}.$$

Given converged estimates of $v_\delta$ and $v_\beta$ from the above procedure, we can than perform an iterative reweighting procedure to obtain optimal weights for $c_{\delta\beta}$.

#### 2.3 Accounting for correlations between nearby variants

Following previous work[3,60], we accounted for correlations between nearby variants by multiplying the weights by the inverse of the LD-score for each SNP and renormalizing the weights such that they sum to 1. This is done at each iteration.

### 3. Random effects meta-analysis of SNP heritability and correlations between effects

In addition to estimating heritability and correlations between effects from meta-analysis summary statistics, we also calculated them at the cohort level to conduct random effects meta-analysis of the estimates. We were interested in whether heterogeneity at the cohort level may result in the random effects meta-analysis estimates of heritability and effect correlations differing estimates from meta-analysis summary statistics.

For each European genetic ancestry cohort, we used LDSC to calculate SNP heritability from DGE and population effect estimates. We used *snipar* to calculate the genome-wide correlations between DGEs and population effects.

After obtaining the estimates, we fit a random effects meta-analysis model to the estimates for each combination of effect and phenotype using the *metafor* package in R (Supplementary Table 3). We excluded cohort-level estimates with a DGE SNP heritability standard error of greater than 0.25 from the analysis.

### 4. Validation phenotypes

#### 4.1. Millennium Cohort Study (MCS)

More information about how we produced and processed each phenotype is provided below and in Supplementary Table 8.

*ADHD:*

We use hyperactivity/inattention scores from the Strengths and Difficulties Questionnaire (SDQ), assessed at age 17 (MCS Sweep 7). This is given as a score from 0-10, which we standardize within sex to have mean 0 and variance 1.

*Age-at-Menarche:*

Data on individual age at menarche was taken from MCS Sweep 7. Outliers were removed (top and bottom 0.5%) and the phenotype was standardized to have mean 0 and variance 1.

*Cannabis:*

Whether or not an individual has ever used cannabis was coded as a binary variable with 0 = never used cannabis and 1 = has used cannabis. This outcome was assessed at age 17 (MCS Sweep 7).

*Cognitive Performance:*

We use two measures of cognitive performance from MCS. The first measure is a word activity score at age 14 (MCS Sweep 6). Individuals were asked to complete a word activity, which was graded out of 20. These scores were standardized within sex to have mean 0 and variance 1.

The second measure is a cognitive assessment score at age 17 (MCS Sweep 7). The assessment comprised 10 questions, with 1 point awarded for the correct answer and 0 otherwise. Individuals who had 6 or more missing answers were excluded; missing values were otherwise imputed using conditional means. The total number of correct answers (including imputed values) was then summed to get a total score for each individual. Scores were then standardized within sex to have mean 0 and variance 1.

*Depression:*

Whether or not an individual has depression was coded as a binary variable with 0 = no depression and 1 = has depression. This outcome was measured at age 17 (MCS Sweep 7).

*Depressive Symptoms:*

Depressive symptoms are measured using the Kessler 6 (K6) scale. The outcome variable ranges from 0 to 24, based off individuals' responses to six questions about depressive and anxiety symptoms experienced in the last 30 days, assessed at age 17 (MCS Sweep 7). We standardize the phenotype to have mean 0 and variance 1 within each sex.

*Drinks-per-week:*

For this phenotype, we use a measure of how many drinks an individual has consumed in the last 4 weeks, taken from MCS Sweep 7. The raw data is a binned variable with 7 possible responses: never, 1-2 times, 3-5 times, 6-9 times, 10-19 times, 20-39 times, or 40 or more times. We use the midpoint of each bin, with 0 for "never" and 40 for "40 or more times." We then standardize the variable within each sex to have mean 0 and variance 1.

*Educational Achievement:*

We produced a measure of educational achievement at age 16 from MCS data across individuals from England, Wales and Northern Ireland using the following procedure:

1. Calculated the expected exam taking year for each student based on birth year and month.
2. For each of GCSE and iGCSE collect the grades achieved for English Language and Mathematics for each student on one row, as well as binary variables denoting whether they studied for each qualification.
3. If a student has an iGCSE but not a GCSE in a subject, use the iGCSE grade to fill in the GCSE grade for that subject. From now on, iGCSEs will be treated like GCSEs.
4. Use the population grade distributions for each subject, year and qualification to convert the grades to expected z-scores, where z is a latent variable representing some unobserved, normally distributed notion of educational attainment. To go from the population grade distribution to expected z-score for each grade, we simulate z from a standard normal distribution, and calculate the mean z for those who would have attained a certain grade. Then, we convert from grade to expected z-score for each subject.
5. If it is not possible to match a student to an appropriate grade distribution, then use the most appropriate other distribution. For example, in the case of iGCSEs, we use the most recent (2016) GCSE grade distribution to avoid bias due to the differing quality of students taking iGCSEs. If a student reports legacy grades for GCSEs when they should report reformed grades, take the most recent (2016) legacy GCSE grade distribution.
6. If a student has taken a qualification but has no grade, this will be taken as an indication that the (compulsory) course was not completed, and the student will be excluded from the analysis.
7. Let EA equal the mean of the two z-scores.

We then standardize the phenotype within each sex to have mean 0 and variance 1.

*Ever-smoker:*

Whether or not an individual has ever smoked was assessed at age 17 (MCS Sweep 7) and coded as a 0/1 binary variable, with 0 = never smoked cigarettes and 1 = has smoked cigarettes. In the questionnaire, cohort members were asked to choose 1 of 6 options: "I have never smoked cigarettes," "I have only ever tried smoking cigarettes once," "I used to smoke cigarettes sometimes but I never smoke a cigarette now," "I sometimes smoke cigarettes now but I don't smoke as many as one a week," "I usually smoke between one and six cigarettes a week," or "I usually smoke more than six cigarettes a week." If the respondent chose "I have never smoked cigarettes," this was coded as 0; all other responses were coded as 1.

*Extraversion:*

Extraversion was measured using the OCEAN extraversion subscale, which gives a score from 3 to 21 based off answers to three questions assessing extraversion as part of the OCEAN ("Big 5") personality trait test in MCS Sweep 7. For each question, cohort members were asked to rate how much the provided statement applied to them using a scale of 1 to 7, with 1 representing "does not apply to me at all" and 7 being "applies to me perfectly." We removed individuals with scores below 3 and standardized the variable to have mean 0 and variance 1 within each sex.

*Height and BMI:*

We use height and BMI at age 17 (MCS Sweep 7). Outliers are removed (top and bottom 0.5%), and each phenotype is standardized within sex to have mean 0 and variance 1.

*Household Income:*

We used OECD equivalized weekly family income from MCS Sweep 6 for the household income phenotype. The variable was standardized within each sex to have mean 0 and variance 1.

*Neuroticism:*

Neuroticism was measured using the OCEAN extraversion subscale, which gives a score from 3 to 21 based off answers to three questions assessing neuroticism as part of the OCEAN ("Big 5") personality trait test in MCS Sweep 7. For each question, cohort members were asked to rate how much the provided statement applied to them using a scale of 1 to 7, with 1 representing "does not apply to me at all" and 7 being "applies to me perfectly." We removed individuals with scores below 3 and standardized the variable to have mean 0 and variance 1 within each sex.

*Self-rated Health:*

Self-rated health was assessed in MCS Sweep 7, whereby respondents were asked to rank their general health on a scale of 1 to 5, with 1 meaning "excellent" and 5 representing "poor." We reversed the order of the scale so that higher values corresponded to higher self-rated health, and standardized the scores to have mean 0 and variance 1 within each sex.

*Subjective Wellbeing:*

To measure subjective wellbeing, we used the sum of raw mental wellbeing scores transformed to a metric scale, assessed at age 17 (MCS Sweep 7). Scores ranged from a minimum of 7 to a maximum of 35. We standardized scores within each sex to have a mean of 0 and variance of 1.

#### 4.2. UK Biobank (UKB)

The validation sample used in the UKB is the same as used in Young et al[3]: all genotyped individuals identified by UKB as part of the 'White British' subsample[41] who had at least one genotyped sibling or parent and who passed quality control filters (no excess heterozygosity, no aneuploidy, no excess relatives).

##### 4.2.1. UKB validation phenotype descriptions

*Age at First Birth (Women):*

See Okbay et al.[17] (2022), Supplementary Table 9.

*Allergic Rhinitis:*

See Becker et al.[61] (2021), Supplementary Table 5.

*Asthma:*

See Becker et al. (2021), Supplementary Table 5.

*Educational Attainment (EA):*

See Okbay et al. (2022) Supplementary Table 9 and Supplementary Note Section 1.

*Cognitive Performance:*

See Okbay et al. (2022), Supplementary Table 9.

*HDL Cholesterol:*

For each of the two measurements, logarithm of HDL cholesterol (field 30760) is residualized on an age indicator (age winsorized at 50 and 78)[62]; sex; 5-year age bin – sex interactions; 20-quantiles of sampling time (field 3166); fasting time (field 74), winsorized at 1 and 18; 20-quantiles of estimated sample dilution factor (field 30897); assessment center (field 54); assessment month; and HDL aliquot (field 30762). The phenotype was set to the mean of the two residuals. Outliers were removed (top and bottom 0.05%), and the phenotype was standardized within each sex to have a mean of 0 and variance of 1.

*Eczema:*

See Becker et al. (2021), Supplementary Table 5.

*Non-HDL Cholesterol:*

Computed as total cholesterol minus HDL cholesterol, where HDL cholesterol is computed as above and total cholesterol is computed by adjusting total cholesterol (field 30690) for statin usage by multiplying it by 1.33588 if field 20003 is coded as one of the following: 1140861958, 1140888594, 1140888648, 1141146234, 1141192410, 1140861922, 1141146138. This was done for each of the two measurements. Then logarithm of adjusted total cholesterol was then residualized on and age indicator (age winsorized at 50 and 78); sex; 5-year age bin – sex interactions; 20-quantiles of sampling time (field 3166); fasting time (field 74), winsorized at 1 and 18; 20-quantiles of estimated sample dilution factor (field 30897); assessment center (field 54); assessment month; and total cholesterol aliquot (field 30692). The phenotype was set to the mean of the two residuals. Outliers were removed (top and bottom 0.05%), and the phenotype was standardized within each sex to have a mean of 0 and variance of 1.

*Diastolic Blood Pressure:*

Diastolic blood pressure was calculated by first taking the average of four readings (two automatic: field 4079, two manual: field 94. The obtained value was then adjusted for medication by adding 10 if any of field 6153 or field 6177 is coded 2. The value was set to missing if information about medication is missing (both of fields 6153 and 6177 are missing, -1 or -3). Finally, the value was residualized on sex, third-degree polynomial in age, and all age-sex interactions. Outliers were removed (top and bottom 0.05%), and the phenotype was standardized within each sex to have a mean of 0 and variance of 1.

*Systolic Blood Pressure:*

The systolic blood pressure phenotype was generated by first taking the average of four readings (two automatic: field 4080, two manual: field 93). The obtained value was then adjusted for medication by adding 15 if any of field 6153 or field 6177 is coded 2. The value was set to missing if information about medication is missing (both of fields 6153 and 6177 are missing, -1 or -3). Finally, the value was residualized on sex, third-degree polynomial in age, and all age-sex interactions. Outliers were removed (top and bottom 0.05%), and the phenotype was standardized within each sex to have a mean of 0 and variance of 1.

*Cigarettes-per-day:*

See Okbay et al. (2022), Supplementary Table 9.

*Forced Expiratory Volume in One Second (FEV1):*

See Okbay et al. (2022), Supplementary Table 9.

*Individual Income:*

Individual income was imputed from occupation using survey data; and transformed to log scale. See Supplementary Materials Section 2.1.4 of Kweon et al. (2024). It was then standardized within each sex to have mean 0 and variance 1.

*Migraine:*

See Becker et al. (2021), Supplementary Table 5.

*Morning Person:*

The morning person phenotype was coded by setting the variable equal to 2 if the respondent reported being “Definitely a morning person”; set to -2 if they responded “Definitely an evening person”; set to 0 if “Do not know”; set to 1 if “More a ‘morning’ than an ‘evening’ person”; set to -1 if “More an ‘evening’ than a ‘morning’ person”; and set to NA if “Prefer not to answer.” The phenotype was then standardized within each sex to have mean 0 and variance 1.

*Myopia:*

See Becker et al. (2021), Supplementary Table 5.

*Number of Children:*

See Okbay et al. (2022), Supplementary Table 9.

### 5. Additional Acknowledgements

**Botnia Family Study** — The Botnia Family Study has been financially supported by grants from Folkhälsan Research Foundation, the Sigrid Juselius Foundation, The Academy of Finland (grants no. 263401, 267882, 312063, 336822, 312072 and 336826), University of Helsinki, Nordic Center of Excellence in Disease Genetics, EU (EXGENESIS, MOSAIC FP7-600914), Ollqvist Foundation, Swedish Cultural Foundation in Finland, Finnish Diabetes Research Foundation, Foundation for Life and Health in Finland, Finnish Medical Society, State Research Funding via the Helsinki University Hospital, Perklén Foundation, Närpes Health Care Foundation and Ahokas Foundation. The study has also been supported by the Municipal Heath Care Center and Hospital in Jakobstad and Health Care Centers in Vasa, Närpes and Korsholm. The research leading to these results has received funding from the European Research Council under the European Union's Seventh Framework Programme (FP7/2007-2013) / ERC grant agreement n° 269045. The role of the founding PI, Professor Leif Groop, in designing the study and the skillful assistance of the Botnia Study Group is gratefully acknowledged. The genotyping was performed by Regeneron Pharmaceuticals. The study sponsors had no role in the analysis and interpretation of the data, writing of the manuscript or decision to submit the manuscript for publication.

Dataset Profile:

Lyssenko V, Almgren P, Anevski D, Perfekt R, Lahti K, Isomaa B, Forsen B, Nissén M, Homström N, Saloranta C, Taskinen M-R, Groop L and Tuomi T. Predictors and longitudinal changes in insulin sensitivity and secretion preceding onset of type 2 diabetes. Diabetes 2005; 54:166-174.

**China Kadoorie Biobank —** We gratefully acknowledge the contributions of the participants, the project staff, and the China National Centre for Disease Control and Prevention and its regional offices for assisting with the fieldwork. The CKB baseline survey and the first re-survey were supported by the Kadoorie Charitable Foundation in Hong Kong. Long-term follow-up of the CKB study was supported by grants from the Wellcome Trust (212946/Z/18/Z,

202922/Z/16/Z, 104085/Z/14/Z, 088158/Z/09/Z) and from the National Key Research and Development Program of China (2016YFC0900500, 2016YFC0900501, 2016YFC0900504, 2016YFC1303904) and the National Natural Science Foundation of China (91843302). DNA extraction and genotyping was supported by grants from GlaxoSmithKline and the UK Medical Research Council (MC-PC- 13049, MC-PC-14135). The project was supported by core funding from the UK Medical Research Council (MC_UU_00017/1,MC_UU_12026/2 MC_U137686851), Cancer Research UK (C16077/A29186; C500/A16896) and the British Heart Foundation (CH/1996001/9454) to the Clinical Trial Service Unit and Epidemiological Studies Unit at Oxford University. Computation used the Oxford Biomedical Research Computing (BMRC) facility, a joint development between the Wellcome Centre for Human Genetics and the Big Data Institute supported by Health Data Research UK and the NIHR Oxford Biomedical Research Centre; the views expressed are those of the author(s) and not necessarily those of the NHS, the NIHR, or the Department of Health.

Dataset Profile:

Chen, Z. et al. China Kadoorie Biobank of 0.5 million people: survey methods, baseline characteristics and long-term follow-up. Int. J. Epidemiol. 40, 1652–1666 (2011).

Walters, R. G. et al. Genotyping and population characteristics of the China Kadoorie Biobank. Cell Genom. 3, 100361 (2023).

**Finnish Twin Cohort** — Phenotype and genotype data collection in the Finnish Twin Cohort has been supported by  the Wellcome Trust Sanger Institute, the Broad Institute, ENGAGE – European Network for Genetic and Genomic Epidemiology, FP7-HEALTH-F4-2007, grant agreement number 201413, National Institute of Alcohol Abuse and Alcoholism (grants AA-12502, AA-00145, and AA-09203 to R J Rose; AA15416 and AA018755 to D M Dick; R01AA015416 to J Salvatore) and the Academy of Finland (grants 100499, 205585, 118555, 141054, 264146, 308248 to JKaprio) and Academy of Finland Center of Excellence in Complex Disease Genetics (grant # 352792 to Kaprio).

Dataset Profile:

Kaidesoja M, Aaltonen S, Bogl LH, Heikkilä K, Kaartinen S, Kujala UM, Kärkkäinen U, Masip G, Mustelin L, Palviainen T, Pietiläinen KH, Rottensteiner M, Sipilä PN, Rose RJ, Keski-Rahkonen A, Kaprio J. FinnTwin16: A Longitudinal Study from Age 16 of a Population-Based Finnish Twin Cohort. Twin Res Hum Genet. 2019 ;22(6):530-539. doi: 10.1017/thg.2019.106. . PMID: 31796134.

Rose RJ, Salvatore JE, Aaltonen S, Barr PB, Bogl LH, Byers HA, Heikkilä K, Korhonen T, Latvala A, Palviainen T, Ranjit A, Whipp AM, Pulkkinen L, Dick DM, Kaprio J. FinnTwin12 Cohort: An Updated Review. Twin Res Hum Genet. 2019;22(5):302-311. doi: 10.1017/thg.2019.83. . PMID: 31640839;

Kaprio J, Bollepalli S, Buchwald J, Iso-Markku P, Korhonen T, Kovanen V, Kujala U, Laakkonen EK, Latvala A, Leskinen T, Lindgren N, Ollikainen M, Piirtola M, Rantanen T, Rinne J, Rose RJ, Sillanpää E, Silventoinen K, Sipilä S, Viljanen A, Vuoksimaa E, Waller K. The Older

Finnish Twin Cohort - 45 Years of Follow-up. Twin Res Hum Genet. 2019;22(4):240-254. doi: 10.1017/thg.2019.54.. PMID: 31462340.

**Estonian Biobank.** We thankfully acknowledge the contributions of the participants, recruitment project staff and investigators of the Estonian biobank (EstBB). The activities of the EstBB are regulated by the Human Genes Research Act, adopted in 2000 specifically for the operations of the EstBB. All participants have provided broad written consent that covers the provision of samples for future research use along with the acquisition of electronic health records from national registries and databases. As a general population biobank, the EstBB is managed by the Institute of Genomics at the University of Tartu. The EstBB cohort currently contains genotype data, health information and metabolic profiles for more than 212,000 participants, representing about 20% of Estonia's adult population.

**Finngen** — We want to acknowledge the participants and investigators of the FinnGen study. The FinnGen project is funded by two grants from Business Finland (HUS 4685/31/2016 and UH 4386/31/2016) and the following industry partners: AbbVie Inc., AstraZeneca UK Ltd, Biogen MA Inc., Bristol Myers Squibb (and Celgene Corporation & Celgene International II Sàrl), Genentech Inc., Merck Sharp & Dohme LCC, Pfizer Inc., GlaxoSmithKline Intellectual Property Development Ltd., Sanofi US Services Inc., Maze Therapeutics Inc., Janssen Biotech Inc, Novartis AG, and Boehringer Ingelheim International GmbH. Following biobanks are acknowledged for delivering biobank samples to FinnGen: Auria Biobank (www.auria.fi/biopankki), THL Biobank (www.thl.fi/biobank), Helsinki Biobank (www.helsinginbiopankki.fi), Biobank Borealis of Northern Finland (https://www.ppshp.fi/Tutkimus-ja-opetus/Biopankki/Pages/Biobank-Borealis-briefly-in-English.aspx), Finnish Clinical Biobank Tampere (www.tays.fi/en-US/Research_and_development/Finnish_Clinical_Biobank_Tampere), Biobank of Eastern Finland (www.ita-suomenbiopankki.fi/en), Central Finland Biobank (www.ksshp.fi/fi-FI/Potilaalle/Biopankki), Finnish Red Cross Blood Service Biobank (www.veripalvelu.fi/verenluovutus/biopankkitoiminta), Terveystalo Biobank (www.terveystalo.com/fi/Yritystietoa/Terveystalo-Biopankki/Biopankki/) and Arctic Biobank (https://www.oulu.fi/en/university/faculties-and-units/faculty-medicine/northern-finland-birth-cohorts-and-arctic-biobank). All Finnish Biobanks are members of BBMRI.fi infrastructure (https://www.bbmri-eric.eu/national-nodes/finland/). Finnish Biobank Cooperative -FINBB (https://finbb.fi/) is the coordinator of BBMRI-ERIC operations in Finland. The Finnish biobank data can be accessed through the Fingenious® services (https://site.fingenious.fi/en/) managed by FINBB.

**Framingham Heart Study (FramHS)** – The Framingham Heart Study is conducted and supported by the National Heart, Lung, and Blood Institute (NHLBI) in collaboration with Boston University (Contract No. N01-HC-25195 and HHSN268201500001I). Funding for SHARe Affymetrix genotyping was provided by NHLBI Contract N02-HL64278. SHARe Illumina genotyping was provided under an agreement between Illumina and Boston University. Funding for Affymetrix genotyping of the FHS Omni cohorts was provided by Intramural NHLBI funds from Andrew D. Johnson and Christopher J. O'Donnell. Funding support for the Framingham Food Frequency Questionnaire dataset was provided by ARS Contract #53- 3k06-5-10, ARS Agreement #'s 58-1950-9-001, 58-1950-4-401 and 58-1950-7-707. This manuscript was not prepared in collaboration with investigators of the Framingham Heart Study and does not necessarily reflect the opinions or views of the Framingham Heart Study, Boston University, or NHLBI. The datasets used for the analyses described in this manuscript were obtained from dbGaP at http://www.ncbi.nlm.nih.gov/sites/entrez?db=gap through dbGaP accession phs000007.v29.p10.

**Geisinger** — The authors would like to acknowledge the participants of the MyCode Community Initiative for the use of their health and genomic information, without whom this study would not be possible. The patient enrollment and exome sequencing were funded by the Regeneron Genetics Center. Data for this project was made possible by the Geisinger-Regeneron DiscovEHR Collaboration.

**GSII (Generation Scotland)** — We would like to acknowledge the contributions of the families who took part in the Generation Scotland: Scottish Family Health Study, the general practitioners and Scottish School of Primary Care for their help in recruiting them, and the whole Generation Scotland team, which includes academic researchers, IT staff, laboratory technicians, statisticians and research managers. Generation Scotland received core support from the Chief Scientist Office of the Scottish Government Health Directorates [CZD/16/6] and the Scottish Funding Council [HR03006] and is currently supported by the Wellcome Trust [216767/Z/19/Z]. Genotyping of the GS:SFHS samples was carried out by the Genetics Core Laboratory at the Edinburgh Clinical Research Facility, University of Edinburgh, Scotland and was funded by the Medical Research Council UK and the Wellcome Trust (Wellcome Trust Strategic Award "STratifying Resilience and Depression Longitudinally" (STRADL) Reference 104036/Z/14/Z). We are grateful to all the families who took part, the general practitioners and the Scottish School of Primary Care for their help in recruiting them, and the whole Generation Scotland team, which includes interviewers, computer and laboratory technicians, clerical workers, research scientists, volunteers, managers, receptionists, healthcare assistants and nurses. Information on applications for access to Generation Scotland data can be found at http://www.generationscotland.org/

**HUNT -** The Trøndelag Health Study (The HUNT Study) is a collaboration between HUNT Research Centre (Faculty of Medicine and Health Sciences, NTNU, Norwegian University of Science and Technology), Trøndelag County Council, Central Norway Regional Health Authority, and the Norwegian Institute of Public Health. The genotyping in HUNT was financed by the National Institutes of Health; University of Michigan; the Research Council of Norway; the Liaison Committee for Education, Research and Innovation in Central Norway; and the Joint

Research Committee between St Olavs hospital and the Faculty of Medicine and Health Sciences, NTNU. The genetic investigations of the HUNT Study are a collaboration between researchers from the HUNT Center for Molecular and Clinical Epidemiology (formerly known as the K.G. Jebsen Center for Genetic Epidemiology as of August 1st 2023), NTNU, and the University of Michigan Medical School and the University of Michigan School of Public Health. We thank HUNT participants for donating their time, samples, and information to help others; clinicians and other employees at Nord-Trøndelag Hospital Trust for their support and for contributing to data collection.

The genotyping in HUNT and work presented in this study was approved by the Regional Committee for Ethics in Medical Research, Central Norway (application numbers 2014/144, 2015/1197, 2015/575, 2015/586, 2015/616, 2017/2479, 2018/2488). All participants signed informed consent for participation and the use of data in research.

Thanks to Bjørn Olav Åsvold, Kristian Hveem, Arnulf Langhammer, Laxmi Bhatta, Ailin Falkmo Hansen, Bendik S. Winsvold, and Ben Brumpton for their contributions to this manuscript before their decision to withdraw.

Cohort profile:

Brumpton BM, Graham S, Surakka I, Skogholt AH, Løset M, Fritsche LG, Wolford B, Zhou W, Nielsen JB, Holmen OL, Gabrielsen ME, Thomas L, Bhatta L, Rasheed H, Zhang H, Kang HM, Hornsby W, Moksnes MR, Coward E, Melbye M, Giskeødegård GF, Fenstad J, Krokstad S, Næss M, Langhammer A, Boehnke M, Abecasis GR, Åsvold BO, Hveem K, Willer CJ. The HUNT study: A population-based cohort for genetic research. Cell Genom. 2022 Oct 12;2(10):100193. doi: 10.1016/j.xgen.2022.100193. PMID: 36777998; PMCID: PMC9903730.

Åsvold BO, Langhammer A, Rehn TA, Kjelvik G, Grøntvedt TV, Sørgjerd EP, Fenstad JS, Heggland J, Holmen O, Stuifbergen MC, Vikjord SAA, Brumpton BM, Skjellegrind HK, Thingstad P, Sund ER, Selbæk G, Mork PJ, Rangul V, Hveem K, Næss M, Krokstad S. Cohort Profile Update: The HUNT Study, Norway. Int J Epidemiol. 2023 Feb 8;52(1):e80-e91. doi: 10.1093/ije/dyac095. PMID: 35578897; PMCID: PMC9908054.

Krokstad S, Langhammer A, Hveem K, Holmen TL, Midthjell K, Stene TR, Bratberg G, Heggland J, Holmen J. Cohort Profile: the HUNT Study, Norway. Int J Epidemiol. 2013 Aug;42(4):968-77. doi: 10.1093/ije/dys095. Epub 2012 Aug 9. PMID: 22879362.

**Lifelines Cohort Study** — The Lifelines Biobank initiative has been made possible by funding from the Dutch Ministry of Health, Welfare and Sport, the Dutch Ministry of Economic Affairs, the University Medical Center Groningen (UMCG the Netherlands), University of Groningen and the Northern Provinces of the Netherlands. The generation and management of GWAS genotype data for the Lifelines Cohort Study is supported by the UMCG Genetics Lifelines Initiative (UGLI). UGLI is partly supported by a Spinoza Grant from NWO, awarded to Cisca Wijmenga. The authors wish to acknowledge the services of the Lifelines Cohort Study, the contributing research centers delivering data to Lifelines, and all the study participants.

**The Norwegian Mother, Father, and Child Cohort Study (MoBa)** — The Norwegian Mother, Father, and Child Cohort Study (MoBa) is a prospective population-based pregnancy cohort study conducted by the Norwegian Institute of Public Health. The Norwegian Mother, Father and Child Cohort Study is supported by the Norwegian Ministry of Health and Care Services and the Ministry of Education and Research. We are grateful to all the participating families in Norway who take part in this on-going cohort study.

Thanks to Rosa Catherine Gillespie Cheesman, Eivind Ystrøm, and Alexandra Havdahl for their contributions to this manuscript before their decision to withdraw.

Dataset Profile:

Magnus, P. et al. Cohort profile update: the norwegian mother and child cohort study (moba). Int. J. Epidemiol. 45, 382–388 (2016).

**Minnesota Twins** — MCTFR recruitment, assessment and genotyping was supported in part by USPHS Grants from the National Institute on Alcohol Abuse and Alcoholism (AA09367 and AA11886), the National Institute on Drug Abuse (DA05147, DA13240, and DA024417), and the National Institute on Mental Health (MH066140).

Dataset Profile:

1. Miller, M. B., Basu, S., Cunningham, J., Eskin, E., Malone, S. M., Oetting, W. S., Schork, N., Sul, J. H., Iacono, W. G., & McGue, M. (2012). The Minnesota Center for Twin and Family Research genome-wide association study. Twin Research and Human Genetics, 15(6), 767–774.

2. Wilson, S., Haroian, K., Iacono, W. G., Krueger, R. F., Lee, J. J., Luciana, M., Malone, S. M., McGue, M., Roisman, G. I., & Vrieze, S. (2019). Minnesota Center for Twin and Family Research. Twin Research and Human Genetics, 22(6), 747–752.

**Netherland Twin Register (NTR)**: Funding was obtained from the Netherlands Organization for Scientific Research (NWO) and The Netherlands Organisation for Health Research and Development (ZonMW) grants 904-61-090, 985-10-002, 912-10-020, 904-61-193,480-04-004, 463-06-001, 451-04-034, 400-05-717, Addiction-31160008, 016-115-035, 481-08-011, 400-07-080, 056-32-010, Middelgroot-911-09-032, OCW_NWO Gravity program –024.001.003, NWO-Groot 480-15-001/674, Center for Medical Systems Biology (CSMB, NWO Genomics), NBIC/BioAssist/RK(2008.024), Biobanking and Biomolecular Resources Research Infrastructure (BBMRI –NL, 184.021.007 and 184.033.111), X-Omics 184-034-019; Spinozapremie (NWO- 56-464-14192), KNAW Academy Professor Award (PAH/6635) and University Research Fellow grant (URF) to DIB; Amsterdam Public Health research institute (former EMGO+) , Neuroscience Amsterdam research institute (former NCA) ; the European Community's Fifth and Seventh Framework Program (FP5- LIFE QUALITY-CT-2002-2006, FP7- HEALTH-F4-2007-2013, grant 01254: GenomEUtwin, grant 01413: ENGAGE and grant 602768: ACTION); the European Research Council (ERC Starting 284167, ERC Consolidator 771057, ERC Advanced 230374), Rutgers University Cell and DNA Repository (NIMH U24 MH068457-06), the National Institutes of Health (NIH, R01D0042157-01A1, R01MH58799-03, MH081802, DA018673, R01 DK092127-04, Grand Opportunity grants 1RC2 MH089951, and

1RC2 MH089995); the Avera Institute for Human Genetics, Sioux Falls, South Dakota (USA). Part of the genotyping and analyses were funded by the Genetic Association Information Network (GAIN) of the Foundation for the National Institutes of Health. Computing was supported by NWO through grant 2018/EW/00408559, BiG Grid, the Dutch e-Science Grid and SURFSARA.

Informed consent was obtained from all NTR participants. The study was approved by the Central Ethics Committee on Research Involving Human Subjects of the VU University Medical Centre, Amsterdam, an Institutional Review Board certified by the U.S. Office of Human Research Protections (IRB number IRB00002991 under Federal-wide Assurance-FWA00017598; IRB/institute codes, NTR 03-180).

**QIMR Berghofer Medical Research Institute (QIMR)** — We greatly thank the twins and their families for their participation. Thanks also to Grant Montgomery and his team for DNA collection and processing and to Scott Gordon for quality control and imputation of the genomic data. Data collection in the Australian sample was made possible by multiple grants from National Health and Medical Research Council and the Australian Research Council.

Dataset Profile:

1. Gillespie N, Kirk KM, Heath AC, Martin NG, Hickie I. Somatic distress as a distinct psychological dimension. Social Psychiatry and Psychiatric Epidemiology. 1999;34(9):451–458.

2. Kirk KM, Birley AJ, Statham DJ, Haddon B, Lake RI, Andrews JG, et al. Anxiety and depression in twin and sib pairs extremely discordant and concordant for neuroticism: prodromus to a linkage study. Twin research : the official journal of the International Society for Twin Studies. 2000;3(4):299.

3. Treloar SA, Martin NG, Bucholz KK, Madden PAF, Heath AC. Genetic influences on post-natal depressive symptoms: findings from an Australian twin sample. Psychological Medicine. 1999;29(3):645–654.

4. Wright MJ, Martin NG. The Brisbane Adolescent Twin Study: outline of study methods and research projects. Australian Journal of Psychology. 2004;56:65–78.

**STR (Swedish Twin Registry) —** The Swedish Twin Registry is managed by Karolinska Institutet and receives funding through the Swedish Research Council under the grant no 2017-00641. Genotyping was performed by the SNP&SEQ Technology Platform in Uppsala (www.genotyping.se). The facility is part of the National Genomics Infrastructure supported by the Swedish Research Council for Infrastructures and Science for Life Laboratory, Sweden. The SNP&SEQ Technology Platform is also supported by the Knut and Alice Wallenberg Foundation

**UK Biobank** — This research has also been conducted using the UK Biobank

Resource under Application Numbers 11425 and 12505. Informed consent was

obtained from UK Biobank subjects.

**E.M.T-D.** was supported by NIH grants R01MH120219 and R01AG073593. E.M.T-D. is a member of the Population Research Center and the Center on Aging and Population Sciences at the University of Texas at Austin, which are supported by NIH grants P2CHD042849 and P30AG066614, respectively.
